# Exploring the effect of maternal glycemic traits on offspring cardiometabolic risk factors in adulthood: an intergenerational Mendelian randomization study

**DOI:** 10.1101/2025.05.07.25326949

**Authors:** Laxmi Bhatta, Tom A Bond, Nicole M Warrington, Geng Wang, Marion Denos, David M Evans, Deborah A Lawlor, Bjørn Olav Åsvold, Gunn-Helen Moen, Ben M Brumpton

## Abstract

**Introduction:** Maternal higher circulating glucose levels in pregnancy have been proposed to influence the offspring’s cardiometabolic health in adulthood through developmental mechanisms. Previous observational studies may have been subject to confounding, including by the inheritance of maternal alleles that predispose to hyperglycemia in the offspring.

**Objectives:** We aimed to test the causal effect of maternal glycemic traits on offspring cardiometabolic risk factors in adulthood.

**Methods:** Two-sample intergenerational Mendelian randomization (MR) analyses were performed. Exposures in these MR analyses were maternal fasting glucose, type 2 diabetes, and gestational diabetes. Similar paternal exposures were used as negative controls. Single Nucleotide Polymorphism (SNP)-exposure associations were obtained from recent genome-wide association studies (GWAS) of fasting glucose (61 SNPs), type 2 diabetes (210 SNPs) and gestational diabetes (8 SNPs). SNP-outcome associations were obtained from adjusted GWAS (parental genotype adjusted for offspring genotype) of adult cardiovascular outcomes which we undertook in three European cohorts (n=up to 566,132). Outcomes in these MR analyses were offspring body mass index, waist-to-hip ratio, systolic and diastolic blood pressure, blood glucose, HbA1c, total cholesterol, low- and high-density lipoprotein cholesterol, triglycerides, and C-reactive protein, measured at 24-62 years of age. Birthweight was used as a positive control.

**Results:** The crude observed positive association between maternal glycemic traits and offspring blood glucose and HbA1c completely attenuated to the null after adjusting for the offsprinǵs genotype. We found no strong evidence that higher maternal fasting glucose, type 2 diabetes, or gestational diabetes were causal for adult offspring cardiometabolic risk factors. For example, the MR analyses suggested that 1SD greater maternal fasting glucose resulted in a 0.088 (95% CI: −0.090, 0.267; *P*=0.331) SD difference in mean offspring BMI. Results were similar for type 2 diabetes (−0.007 SD change in BMI per unit increase in log odds; 95% CI: - 0.043, 0.030; *P*=0.726) and gestational diabetes (−0.016 SD change in BMI per unit increase in log odds; 95% CI: −0.105, 0.072; *P*=0.716). In contrast, we found strong evidence that higher maternal (but not paternal) fasting glucose, type 2 diabetes, and gestational diabetes were causal for increased offspring birthweight.

**Conclusions:** Our data suggest that maternal glucose levels do not have a major influence on offspring cardiometabolic risk factors in adulthood.

## Introduction

The Developmental Origins of Health and Disease (DOHaD) hypothesis suggests that maternal health during pregnancy and the *in utero* environment in which the fetus develops, may be critical for offspring later life health^1^. The fetus’ major energy source is maternal circulating glucose, and transplacental passage of maternal glucose occurs by facilitated diffusion with high capacity^2^. Even small increases in maternal glucose levels will, therefore, be transmitted to the fetus and can stimulate growth of both lean and adipose tissues^2^ ^3^. This is seen, for example in gestational diabetes, where mothers are unable to compensate with insulin for the increase in circulating glucose naturally occurring in pregnancy, resulting in maternal hyperglycemia and excess fetal growth^2^ ^3^. Under the DOHaD hypothesis, this fetal overgrowth or the impact of high glucose levels on the fetal pancreas and liver could lead to permanent metabolic changes that predispose the offspring to adiposity, diabetes and adverse cardiovascular health in later life^4^ ^5^. If true, this could result in an intergenerational vicious cycle, whereby the prevalence of cardiometabolic disease inexorably increases over subsequent generations^6^.

Consistent with this hypothesis, higher maternal glucose levels are associated with increased cardiometabolic risk for the offspring in childhood and adolescence^4^ ^7^. However, the observational associations may be influenced by confounding, in particular by genetic and environmental factors shared by parents and their children^8–11^. Randomized controlled trials investigating effects of glucose-lowering treatment in pregnancy on offspring cardiometabolic health in late-life are unfeasible. Consequently, important evidence comes from observational study designs that are less influenced by confounding, such as sibling studies. In the Pima Indians of Arizona, who have extremely high rates of type 2 diabetes, mean BMI and risk of type 2 diabetes at age 2-24 years were greater for children born after their mother had been diagnosed with type 2 diabetes, compared to their siblings born before maternal diagnosis^12^. Exposure to paternal type 2 diabetes (a negative control) was not associated with offspring diabetes risk and BMI within siblings, suggesting a specific effect of intrauterine exposure^12^. Similarly, mean BMI in 18 year old Swedish men was greater for offspring exposed to maternal diabetes during pregnancy than for their unexposed siblings^13^. Because siblings share familial confounders, including maternal genotype, these studies provide suggestive evidence for an intrauterine effect of maternal diabetes on offspring BMI and type 2 diabetes risk. However, sibling studies may be affected by biases, due to carryover^14^ ^15^, where an exposure such as gestational diabetes, or outcome (e.g. large for gestational age infant) influences management and exposures or outcomes in a subsequent pregnancy, and selection bias^16^. Furthermore, to our knowledge no sibling studies have investigated offspring BMI beyond age 24 years, or other offspring cardiometabolic risk factors. Mendelian randomization (MR), which uses observational data and genetic variants as instrumental variables, could be used to improve causal inference^8–10^. MR has previously been used to demonstrate a causal effect of higher maternal glucose levels on higher offspring birthweight^17^ ^18^, but to our knowledge there are no MR studies of the effect of maternal glucose levels and related traits on adult cardiometabolic outcomes.

The aim of this study is to use the novel intergenerational MR framework^19^ developed by us and others to quantify the causal effect of maternal fasting glucose, type 2 diabetes, and gestational diabetes on offspring cardiometabolic risk factors in adulthood. As a positive control (for which we would expect to find a causal effect), we aimed to investigate the effect of maternal glycemic traits on offspring birthweight^17^ ^18^. As a negative control (for which we would not expect to find a causal effect), we aimed to investigate the effect of paternal glycemic traits on offspring cardiometabolic factors.

## Methods

We followed the MR-STROBE reporting guidelines^20^.

### Study design

To perform intergenerational MR in a two-sample MR framework, we used summary statistics from genome wide association studies (GWAS) of the exposures (publicly available) and a GWAS of the outcomes (our analyses)^19^. The outcome GWAS summary statistics included estimates of indirect maternal/paternal genetic effects (adjusted for offspring genotype) and direct offspring genetic effects (adjusted for parental genotype). In intergenerational MR it is important to control for the inheritance of parental alleles by the offspring, which could violate the exclusion restriction assumption^19^ ^21^. Details of the study design are explained below, and a flowchart is presented in **Figure 1**. A two-sample MR design for the intergenerational MR approach and the assumptions made are presented in **Figure 2**. In short, the assumptions on which this intergenerational MR study are based are: i) relevance: the independent genetic instruments (Z) that proxy glycemic trait (X) in the general population are also robustly associated with the relevant maternal glycemic trait (X) during pregnancy; ii) independence: there is no confounding between the genetic instruments (Z) and offspring outcome (Y). This assumption could be violated by for example population stratification or assortative mating; iii) exclusion restriction: the maternal genetic instruments (Z) only potentially influence the offspring outcome (Y) through the maternal glycemic trait (X) during pregnancy. This assumption would be violated for example if alleles inherited by the offspring at the same loci (Z) directly influenced the offspring outcome (Y). Therefore, we used estimates of the maternal indirect genetic effect adjusted for offspring genotype at the locus in question. In addition, if paternal genotype at the same loci affects the offspring outcome (Y) independently of the inherited alleles by offspring (Z), then estimates of maternal and offspring genetic effect should also ideally be adjusted for paternal genotype, because the offspring genotype would be a collider, which once adjusted for will bias maternal genetic effects. This assumption could also be violated by horizontal pleiotropy in the maternal genome; and iv) linearity for continuous trait: both the genetic instruments (Z) – maternal fasting glucose (X) effects and the maternal fasting glucose (X) - offspring outcome (Y) effect are linear, and for each individual the maternal fasting glucose (X) is a monotonic (increasing or decreasing) function of the genetic instruments (Z).

**Figure 1.**
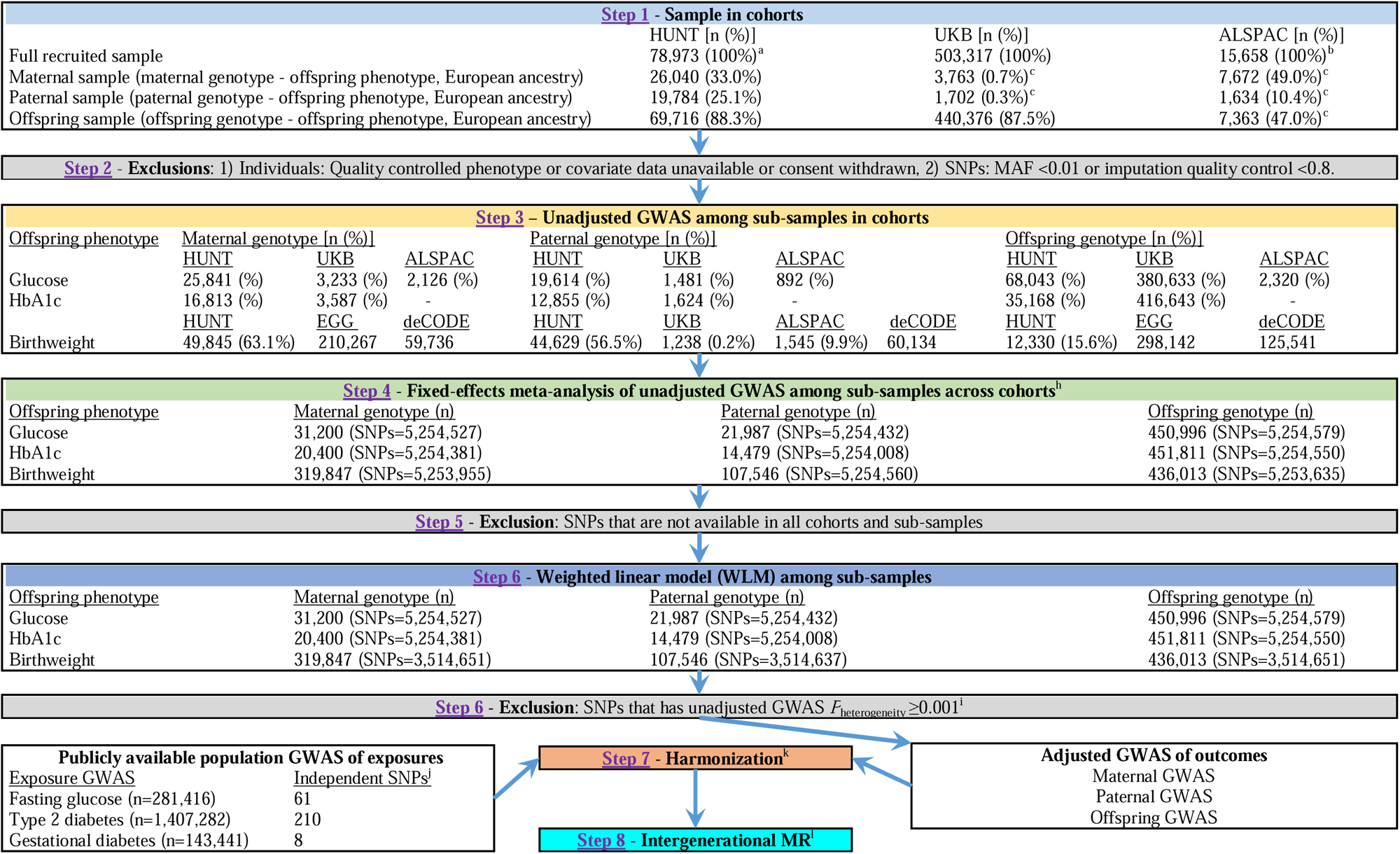
Flow chart of study design including selection of study participants for the adjusted GWAS of offspring outcomes. HUNT (Trøndelag Health Study), UKB (UK Biobank), ALSPAC (Avon Longitudinal Study of Parents and Children), SNP (single nucleotide polymorphism), MAF (minor allele frequency), GWAS (genome-wide association study), HbA1c (glycated haemoglobin), EGG (early growth genetics), deCODE (deCODE genetics), MR (mendelian randomization), ^a^ total number of participant in HUNT2 and HUNT3 study waves, ^b^ total numbers of fetuses included in ALSPAC, ^c^ total number of unrelated maternal, paternal, and offspring sample, ^d^ For offspring birthweight, parent-offspring pairs were identified via medical birth registry (MFR), but for offspring adulthood outcomes, the parent-offspring pairs were identified via kinship analysis^42^ using genotype information, therefore, the sample size of birthweight for maternal and paternal sample were much higher than other offspring adulthood outcomes, ^e^ EGG consortium analyses of birthweight for maternal and offspring sample includes UKB and ALSPAC, ^f^ we retrieved publicly available GWAS summary statistics for EGG^39^ and deCODE^40^, ^g^ sample sizes were similar for other offspring adulthood outcomes, ^h^ Prior to meta-analysis, A/T and C/G SNPs were removed from the birthweight deCODE GWAS^40^, and A/T and C/G SNPs also were removed from our GWAS when comparison of their allele frequency to the HRC or 1000 Genome Project reference panel suggested harmonization errors, ^i^ excluded those SNPs having *p* value ≥0.001 in test of heterogeneity during meta-analysis of unadjusted GWAS, ^j^ independent SNPs were selected that passed threshold *P*<5.0x10^-8^, r^2^<0.001, kb=10000, and MAF≥0.01 and imputation quality score (INFO)≥0.80 in HUNT, UKB, or ALSPAC, ^k^ GWAS summary statistics of exposure and outcome were matched on the genetic instruments of exposure and then the risk alleles were harmonized, ^l^ a two-sample MR approach was used for intergenerational MR.

**Figure 2.**
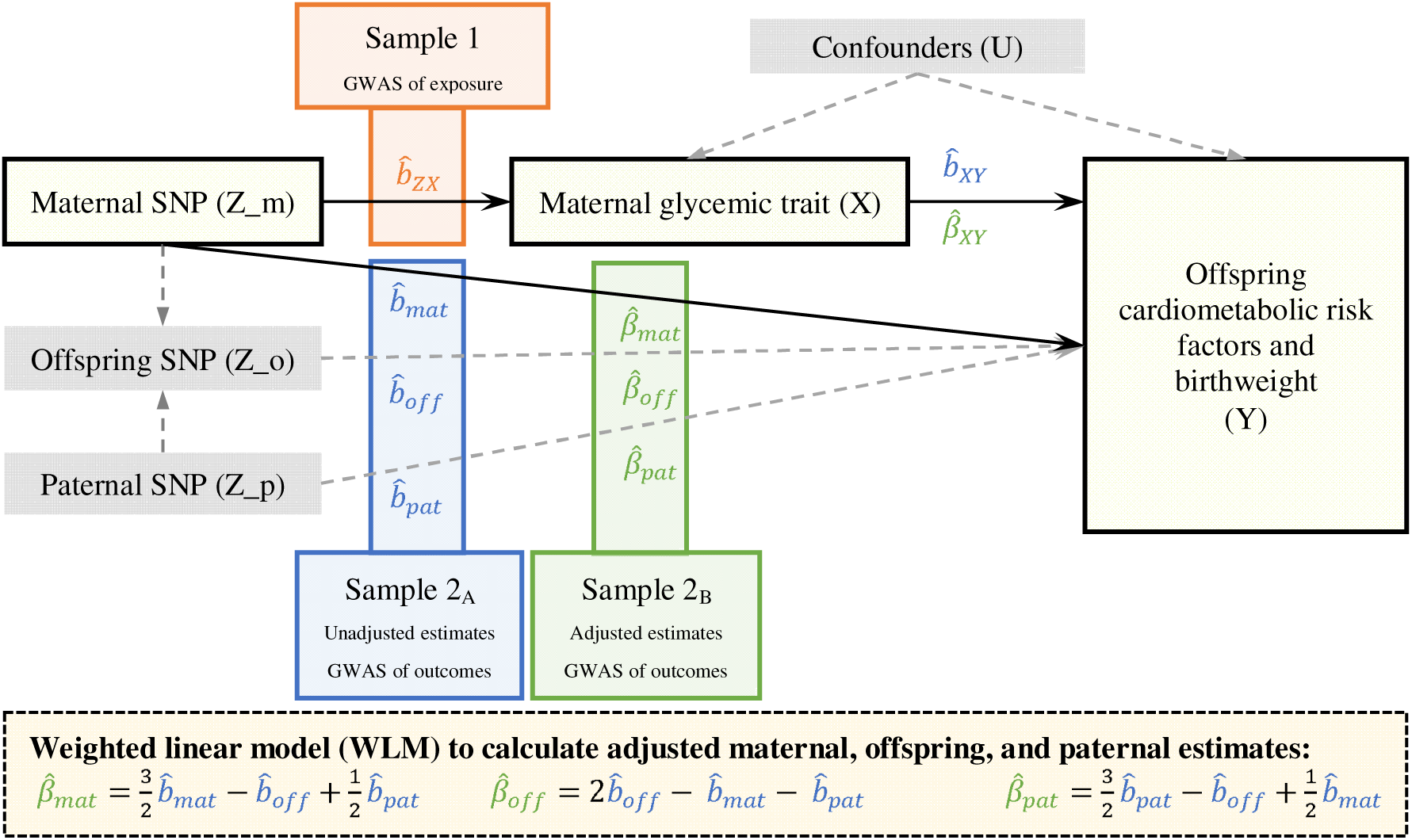
Two-sample MR design for estimating the causal effect of maternal glycemic trait on offspring outcome. The effect of the maternal SNP (Z_m) on maternal glucose (X) 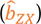 is obtained in Sample 1. The unadjusted effect of the maternal SNP (Z_m) on the offspring outcome (Y) 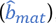, the unadjusted effect of the offspring SNP (Z_o) on the offspring outcome (Y) 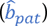, the unadjusted effect of the paternal SNP (Z_p) on the offspring outcome (Y) 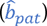 are all obtained in Sample 2_A_. The unadjusted effect of maternal glucose (X) on offspring outcome (Y) 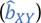 can be estimated by the ratio 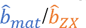, however this estimate is likely to be contaminated by pleiotropy through the offspring genome due to the correlation between maternal and offspring genotypes. The effect of the maternal SNP (Z_m) on offspring outcome (Y) after controlling for offspring and paternal influences 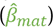, the effect of offspring SNP (Z_o) on offspring outcome (Y) after controlling for maternal and paternal influences 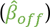 and the effect of the paternal SNP (Z_p) on offspring outcome (Y) after controlling for maternal and offspring influences 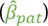 are estimated in Sample 2_B_. The ratio 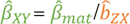 is the estimated adjusted effect of maternal glucose (X) on offspring outcome (Y). Finally, unadjusted 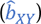 and adjusted estimates 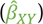 at each SNP can be combined using inverse-variance weighted meta-analysis to estimate the overall unadjusted and adjusted estimate of maternal glucose (X) on offspring outcome (Y), respectively. This intergenerational MR analysis makes several assumptions, including i) the maternal SNPs (Z_m) are robustly associated with the maternal glycemic trait (X) during pregnancy. We used SNPs that were strongly associated with the relevant glycemic trait (*P*<5×10^-8^) in population-based GWAS, and these SNPs explained from 1.0% to 4.9% of the variance in fasting glucose during pregnancy in the Born in Bradford (BiB) cohort (n=3,044), ii) 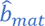 or 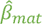 is not confounded i.e. there is no arrow from confounders (U) to maternal SNP (Z_m). This assumption could be violated by population stratification or assortative mating, iii) maternal SNP (Z_m) only influences the offspring outcome (Y) through maternal glucose (X) during pregnancy. This assumption would be violated if the offspring SNP (Z_o) associated with glucose directly influences the offspring outcome (Y). Therefore, we adjusted 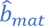 for offspring influences at the locus in question, and also ideally for paternal influences because offspring genotype is a collider, which once adjusted for will bias 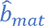, if 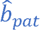 affects the offspring outcome (Y) independently of the offspring SNP (Z_o). This assumption could also be violated by horizontal pleiotropy through the maternal genome, and iv) both maternal SNP (Z_m) - maternal glucose (X) effect 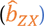 and the maternal glucose (X) - offspring outcome (Y) effect (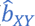 and 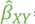 are linear, and for each individual the maternal glucose (X) is a monotonic (increasing or decreasing) function of the maternal SNP (Z_m).

### GWAS of exposures (genetic instruments for glycemic traits)

European ancestry-specific GWAS summary statistics for fasting glucose (mmol/L) were obtained from Chen et. al, 2021 (n=281,416)^22^. European ancestry-specific GWAS summary statistics for type 2 diabetes were obtained from Vujkovic et. al., 2020^23^ (n_case_=228,499; n_control_=1,178,783). We defined independent genetic instruments for fasting glucose (n=68) and type 2 diabetes (n=232) as those that passed clumping with the thresholds kb=10000, r^2^=0.001, and *P*=5.0x10^-8^. We retrieved independent SNPs for gestational diabetes (n=12) reported by Elliott et. al., 2024^24^ (n_case_=12,332; n_control_=131,109) in a European ancestry GWAS. **Supplementary information S1** provides details of the publicly available GWAS data used for the exposures in this study. After exclusion of SNPs that were not available with MAF≥0.01 and imputation quality score (INFO)≥0.80 in each of our three main analysis cohorts (see below), there were 61 independent SNPs for fasting glucose (in the general population), 210 for type 2 diabetes (in the general population), and 8 for gestational diabetes (in pregnant women)^22–24^ (**Figure 1**)).

#### Relevance in pregnancy

We explored the genetic instruments’ relevance for glycemic traits during pregnancy by examining their associations with fasting blood glucose among pregnant women in the Born in Bradford cohort (BiB)^25^. We constructed weighted polygenic risk scores using genetic effect estimates from the original GWAS studies and calculated the variance of maternal fasting and 2-hour post-load glucose (2hGlu) in mid pregnancy (mean 26.2 weeks gestation) explained by each PRS [fasting glucose PRS *R*^2^=4.9% (fasting glucose), 0.6% (2hGlu); type 2 diabetes PRS *R*^2^=1.0% (fasting glucose), 1.1% (2hGlu); and gestational diabetes PRS *R*^2^=3.9% (fasting glucose), 0.6% (2hGlu)] in BiB (n=3,044). Further details of the BiB cohort and analyses are presented in **Supplementary information S2**.

### GWAS of outcomes (offspring cardiometabolic risk factors and birthweight)

#### Study cohorts and genotyping

Our GWAS analyses of outcomes used data from three population-based cohorts: the Trøndelag Health Study (HUNT), UK Biobank (UKB), and the Avon Longitudinal Study of Parents and Children (ALSPAC). **HUNT** invited the entire adult population (≥20 years, ∼230,000 participants) of Trøndelag, Norway to attend clinical examinations and answer questionnaires in four waves from 1984-2019^26–29^. We included HUNT2 (1995 to 1997, n=65,402 participants, 69.7% of those invited) and HUNT3 (2006 to 2008, n=50,663 participants, 54.0% of those invited) in the analyses. Genotyping, imputation, and quality control of HUNT genetic data have been described in detail elsewhere^27^ ^29^. About 88% (n=69,716 unique individuals) of participants from HUNT2 and HUNT3 have genotype data. Furthermore, HUNT was linked to the Medical Birth Registry of Norway (MBRN)^30^, which allowed all the HUNT participants born after 1967 to have a record of their own birthweight and their offsprinǵs birthweight (even if offspring are not participants of HUNT). The validity of this information has been reported elsewhere^31^ (**Figure 1**). **UKB** invited 9.2 million eligible adults (40-69 years, n=503,317 participants, 5.4% of those invited) from the UK between 2006 and 2010 to attend clinical examinations and answer questionnaires^32^. Genotyping, imputation, and quality control of UKB genetic data have been described in detail elsewhere^33^. After quality control and selection of European ancestry there were 440,376 participants from UKB (88% of those participated in UKB) with genotype data (**Figure 1**). **ALSPAC** invited pregnant women resident in Avon, UK with expected dates of delivery between 1^st^ April 1991 and 31st December 1992 (n=14,541 initial enrolled pregnancies, and 80% of women invited participated) to attend clinical examinations and answer questionnaires^34–37^. Further enrolments after 1998 resulted in a baseline sample size of 14,901 children alive at one year of age. We used data collected during pregnancy/birth and the 24-year follow up. Genotyping, imputation, and quality control of ALSPAC genetic data have been described in detail elsewhere^38^ (**Figure 1**). In all cohorts, only individuals of European ancestry, as defined through ancestrally informative principal components, were included in the study. Further details of study cohorts and genotyping is presented in **Supplementary information S3**. In addition to our main analyses cohorts, we used the publicly available summary level data (unadjusted GWAS summary statistics) from the Early Growth Genetics (EGG) Consortium^39^ and deCODE genetics^40^ (further detail in **Supplementary information S3**).

#### Identifying genotyped parent–offspring pairs

**HUNT.** The procedure used to identify parent-offspring pairs has been described elsewhere^11^^41^. In short, a kinship analysis was performed to identify parent-offspring pairs, where we used the recommended thresholds for relatedness implemented in the KING software package v2.2.4^42^. Additionally, parent-offspring pairs with birthweight information were identified via linkage within the MBRN^30^ (**Figure 1**). In **UKB**, a similar procedure (kinship analysis) as in HUNT was used to identify parent-offspring pairs. In **ALSPAC**, parent-offspring pairs were identified by matching study IDs, including a QC check for familial relatedness across the overall father-offspring sample via the genetic data using KING^42^ “--related” command. For UKB and ALSPAC, only unrelated parent-offspring pairs were included in the analyses (**Figure 1**). Further detail on parent-offspring pairs is presented in **Supplementary information S4**.

#### Cardiometabolic risk factors in adulthood and birthweight

Information on body mass index (BMI, kg/m^2^), waist-to-hip ratio (WHR, cm/cm), systolic (SBP, mmHg) and diastolic blood pressure (DBP, mmHg), blood glucose (mmol/L), glycated hemoglobin (HbA1c, mmol/mol), total cholesterol (TC, mmol/L), high-density (HDL-C, mmol/L) and low-density lipoprotein cholesterol (LDL-C, mmol/L), triglycerides (TG, mmol/L), and C-reactive protein (CRP, mg/L) were retrieved from each cohort and collected following similar procedures. Blood samples in HUNT and UKB were non-fasting while in ALSPAC individuals fasted overnight or at least 6-hour prior to blood sampling. CRP was strongly right skewed; hence the values were natural log transformed. As a sensitivity analyses, we log transformed BMI, WHR, glucose, HbA1c, TG, and HDL-C. Additionally, we used offspring birthweight as a positive control outcome, and tested whether we found the expected positive effect of maternal higher glucose levels on higher birthweight. Offspring birthweight was reported by mothers in UKB, obtained from clinical records or measured by trained research assistants in ALSPAC and obtained from the MBRN for HUNT participants. Sex-specific z-score of birthweights (kg) were calculated for analyses. Further details of these outcomes, correction for medication use (blood pressure and lipid lowering medication), and covariates are provided in **Supplementary information S5**.

#### Statistical analysis

##### Maternal, paternal, and offspring GWAS (unadjusted estimates)

In each cohort i.e. HUNT, UKB, and ALSPAC, we included three sub-samples to perform GWASs (**Figure 1**), which were i) maternal (maternal genotype and offspring phenotype), ii) paternal (paternal genotype and offspring phenotype), and iii) offspring (offspring genotype and offspring phenotype). To estimate genome-wide genetic variant-outcome associations, we performed GWAS for offspring outcomes separately in each sample. To account for relatedness in the samples (HUNT: all three samples, UKB: offspring sample), we fit a linear mixed model (LMM) using the fastGWA method^43^ implemented in the GCTA^44^ software package version 1.93.3beta2. When the sample size was insufficient to fit the LMM, we analyzed unrelated individuals only (UKB: maternal and paternal sample, ALSPAC: all three samples) and fit a linear regression implemented in fastGWA. To fit the fastGWA LMM, we constructed a genetic relationship matrix (GRM) for each sample using GCTA with genotyped autosomal variants which passed QC (**Supplementary information S3 and S4**) and had MAF ≥0.01. For UKB, we also filtered SNPs with missingness >0.1 and HWE *P* >1.0x10^−06^ in PLINK version 1.9^45^ prior to GRM calculation. The GRM was calculated using the default sparsity threshold of 0.05. In HUNT, maternal-offspring and paternal-offspring pairs included up to six siblings in the offspring generation. To account for this relatedness whilst fitting the maternal and paternal GWAS models, we included mothers and fathers in the GRM multiple times (once for each of their children) and coded the coefficient between them as one. We performed GWAS for autosomal variants with imputation quality score ≥0.3 and MAF ≥0.01 and applied a more stringent imputation quality score filter (≥0.8) prior to MR analyses. The sample sizes and covariates included in the GWAS models for all the offspring outcomes are presented in **Supplementary information S6 and S7**.

##### Meta-analysis of maternal, paternal, and offspring GWAS (unadjusted estimates)

We performed inverse variance weighted fixed-effect meta-analysis across the three cohorts (HUNT, UKB, and ALSPAC), separately for the maternal, paternal, and offspring GWAS, using METAL software version 2011-03-25^46^ (**Figure 1**). On the pooled estimates of maternal, paternal, and offspring GWAS, we performed standard quality control (QC) procedures^47^ including tests for inflation due to population structure including the genomic control inflation factor (λ) and linkage disequilibrium score regression (LDSC) intercept, and attenuation ratio^48^ ^49^ (**Supplementary information S8**).

For the meta-analysis of birthweight, we additionally included publicly available unadjusted GWAS for the maternal sample from the EGG consortium^39^ (which already included UKB and ALSPAC, therefore we did not rerun this analysis in UKB and ALSPAC) and deCODE genetics cohort^40^ (combined n=270,002), for the paternal sample from deCODE^40^ (n=60,134), and for the offspring sample from EGG^39^ and deCODE^40^ (combined n=423,683) (**Figure 1**). Prior to meta-analysis, pallindromic (A/T and C/G) SNPs were removed from the birthweight deCODE GWAS^40^, and from our GWAS of offspring cardiometabolic risk factors and birthweight when comparison of their allele frequency to the HRC or 1000 Genome Project reference panel suggested strand discrepancies.

##### Indirect maternal/paternal and direct offspring GWAS (adjusted estimates)

Offspring share alleles with their parents, therefore to calculate the indirect maternal, indirect paternal, and direct offspring GWAS summary statistics we used a weighted linear model (WLM)^39^ ^50^ ^51^ implemented in the DONUTS R package^52^. Prior to applying the WLM, we removed genetic variants missing in any of the cohorts meta-analyzed. Using the WLM, we calculated adjusted genetic effects (i.e. the mutually adjusted coefficients for maternal, paternal and offspring genotype, fitted jointly in the same model), as a linear combination of the unadjusted (maternal, paternal, and offspring) genetic effects (**Figure 2, Supplementary information S9**). Additionally, we fit a WLM to the parent-offspring duos, which only adjusted for one parent’s genotype and assumed that the other parent’s genotype had no independent effect on the offspring outcome, but yielded smaller standard errors than the full (trios) WLM (**Supplementary information S9**). The trios WLM results were found to be similar to multivariable linear regression in genotyped parent-offspring trios (unrelated trios sample, n=28,614) from the Norwegian Mother, Father and Child Cohort Study (MoBa, a population based pregnancy cohort)^53–55^. We also confirmed via simulation that adjusted genetic effect estimates from the WLM are unbiased, even when the sample sizes for maternal, paternal and offspring GWAS are drastically different^55^.

### Intergenerational MR analysis

We filtered GWAS summary statistics based on imputation quality score ≥0.8 and unadjusted GWAS meta-analysis between-cohort *P*_heterogeneity_≥0.001 prior to MR analyses.

The intergenerational MR design and its assumptions are illustrated in **Figure 2**. We applied a two-sample MR approach implemented in the TwoSampleMR R package^56^ to estimate the causal effect (maternal-offspring effect) of exposures (using SNP-exposure associations from publicly available GWAS of glycemic traits) on outcomes [using maternal SNP-outcome associations adjusted for offspring genotype from our GWAS of offspring cardiometabolic risk factors and birthweight (a positive control analysis)]. Similarly, for the paternal-offspring MR analyses we used paternal SNP-outcome associations adjusted for offspring genotype from our GWAS analyses. We performed MR analyses using the adjusted GWAS summary statistics of outcomes from both the trios and duos WLM. We do not present MR estimates from the duos WLM for paternal glycemic traits - offspring birthweight association because this would be inappropriate, given the strong effect of maternal glycemic traits on offspring birthweight^17^ ^18^ and the possible risk of collider bias. We reported inverse variance weighted (IVW) MR estimates [beta (95% confidence interval (CI))] as the main results. Additionally, sensitivity analyses including MR Egger^57^, weighted median^58^, and weighted mode^59^ estimates were performed, which made less stringent assumptions regarding horizontal pleiotropy than the IVW analysis. We calculated F-statistics^60^ to test the strength of the SNPs as instruments of maternal glycemic traits. MR Egger intercept^57^, MR-PRESSO global test^61^, and between-SNPs MR heterogeneity test (Cohrańs Q test)^62^ were performed to test for horizontal pleiotropy. We interpreted *P*<4.2x10^-3^ (0.05/12 offspring outcomes) as strong evidence against the null hypothesis. The MR estimates were re-scaled to the standard deviation (SD) scale to facilitate comparison between outcomes, as described in **Supplementary information S10**. The estimates for binary exposures (type 2 diabetes, gestational diabetes) were presented on the log odds scale, which can be multiplied by log_e_(2) to enable interpretation of estimates as SD change in the outcome per doubling of the prevalence of the exposure^60^. The genetic instruments for glycemic traits and the harmonized datafile for exposure and outcomes used in the MR analyses are presented in **Supplementary table S1**.

#### Software

The statistical software used for analyses were PLINK version 1.9^45^, QCTOOL v2.0.7^63^, KING software package v2.2.4^42^, GCTA software package version 1.93.3beta2^44^, and R version 4.0.3^64^.

#### Ethics

Ethical approval was received from the Regional Committee for Ethics in Medical Research, Central Norway (REK Central application number 2018/2488) (HUNT), the North West Multi-centre Research Ethics Committee (MREC) (ref 11/NW/0382) (UKB), the ALSPAC Ethics and Law Committee and the Local Research Ethics Committees (ALSPAC), the Bradford National Health Service Ethics Committee (ref 06/Q1202/48) (Born in Bradford). Informed consent was obtained from all participants. We excluded all participants who had withdrawn consent to participate in the study.

## Results

In our GWAS meta-analysis of outcomes, the analyses included up to 319,847 mothers, 107,546 fathers, and 511,253 offspring from the HUNT, UK Biobank, and ALSPAC cohorts and publicly available GWAS including EGG consortium and deCODE cohort (**Figure 1, Table 1, Supplementary information S6**). The descriptive characteristics of the participants included in our GWASs are presented in **Table 1**. Mean age of the offspring ranged from 39.2-62.0 years in HUNT, 43.1-56.8 years in UKB, and was 24.5 years in ALSPAC. The proportion of females ranged from 48.8-55.7% in HUNT, 53.7-64.7% in UKB, and 46.0-61.3% in ALSPAC. GWAS quality control statistics including the genomic control inflation factor (λ), LDSC intercept, and LDSC attenuation ratio for each GWAS meta-analysis are reported in **Supplementary information S8**.

**Table 1.** Descriptive characteristics of participants in HUNT, UKB, and ALSPAC cohorts.

| Offspring outcome (units) | Maternal genotype GWAS |  |  |  | Paternal genotype GWAS |  |  |  | Offspring genotype GWAS |  |  |  |
| --- | --- | --- | --- | --- | --- | --- | --- | --- | --- | --- | --- | --- |
|  | *n* | Mean (SD) or median (IQR) offspring outcome<sup>a</sup> | Mean (SD) offspring age (years) | Offspring % female | *n* | Mean (SD) or median (IQR) offspring outcome<sup>a</sup> | Mean (SD) offspring age (years) | Offspring % female | *n* | Mean (SD) or median (IQR) offspring outcome<sup>a</sup> | Mean (SD) offspring age (years) | Offspring % female |
| **HUNT** |  |  |  |  |  |  |  |  |  |  |  |  |
| BMI (kg/m<sup>2</sup>) | 25998 | 26.6 (23.9, 29.7) | 49.1 (14.7) | 51.6 | 19757 | 26.5 (23.8, 29.6) | 47.1 (14.2) | 51.6 | 68316 | 26.7 (24.1, 29.8) | 59.5 (17.0) | 52.7 |
| WHR (cm/cm) | 25953 | 0.93 (0.86, 0.99) | 49.0 (14.7) | 51.5 | 19709 | 0.92 (0.86, 0.99) | 47.0 (14.1) | 51.4 | 68294 | 0.92 (0.86, 0.98) | 59.3 (16.7) | 52.7 |
| SBP (mmHg) | 26013 | 131.2 (19.2) | 49.1 (14.7) | 51.6 | 19761 | 129.4 (18.3) | 47.1 (14.2) | 51.6 | 68583 | 139.9 (23.9) | 59.6 (17.0) | 52.8 |
| DBP (mmHg) | 26011 | 76.1 (11.7) | 49.1 (14.7) | 51.6 | 19760 | 75.34 (11.5) | 47.1 (14.2) | 51.6 | 68555 | 78.96 (13.1) | 59.6 (17.0) | 52.8 |
| Glucose (mmol/L) | 25841 | 5.10 (4.70, 5.60) | 41.3 (12.7) | 51.6 | 19614 | 5.10 (4.70, 5.60) | 39.2 (12.0) | 51.6 | 68043 | 5.3 (4.80, 5.90) | 53.4 (17.4) | 52.9 |
| HbA1c (mmol/mol) | 16813 | 34.0 (32.0, 36.0) | 55.2 (11.3) | 53.9 | 12855 | 33.0 (31.0, 36.0) | 53.3 (10.8) | 53.8 | 35168 | 34.0 (32.0, 37.0) | 62.0 (13.6) | 55.3 |
| TC (mmol/L) | 25960 | 5.35 (1.10) | 49.1 (14.7) | 51.6 | 19715 | 5.31 (1.08) | 47.1 (14.2) | 51.6 | 68501 | 5.58 (1.23) | 59.6 (17.0) | 52.8 |
| HDL-C (mmol/L) | 25958 | 1.31 (1.10, 1.59) | 49.1 (14.7) | 51.6 | 19714 | 1.31 (1.10, 1.59) | 47.1 (14.2) | 51.6 | 68502 | 1.3 (1.10, 1.60) | 59.6 (17.0) | 52.8 |
| LDL-C (mmol/L) | 25753 | 3.24 (0.96) | 48.9 (14.8) | 51.9 | 19572 | 3.21 (0.94) | 46.9 (14.2) | 51.8 | 67635 | 3.43 (1.08) | 59.5 (17.1) | 53.1 |
| TG (mmol/L) | 26028 | 1.40 (0.98, 2.03) | 49.0 (14.7) | 51.6 | 19773 | 1.38 (0.96, 2.00) | 47.0 (14.2) | 51.6 | 68627 | 1.49 (1.05, 2.12) | 59.6 (17.0) | 52.8 |
| CRP (mg/L)<sup>b</sup> | 22375 | 1.19 (0.58, 2.54) | 51.6 (13.6) | 52.6 | 17110 | 1.13 (0.55, 2.44) | 49.6 (13.1) | 52.6 | 51581 | 1.33 (0.65, 2.90) | 60.2 (15.8) | 53.9 |
| Birthweight (kg)<sup>c</sup> | 49845 | 3.519 (0.635) | NA | 48.8 | 44629 | 3.530 (0.624) | NA | 48.8 | 12330 | 3.519 (0.546) | NA | 55.7 |
| **UKB** |  |  |  |  |  |  |  |  |  |  |  |  |
| BMI (kg/m<sup>2</sup>) | 3742 | 26.0 (23.3, 29.0) | 43.1 (2.3) | 61.1 | 1692 | 25.9 (23.2, 28.9) | 42.6 (2.0) | 58.3 | 438258 | 26.7 (24.1, 29.9) | 56.8 (8.0) | 54.1 |
| WHR (cm/cm) | 3756 | 0.84 (0.77, 0.90) | 43.1 (2.3) | 61.1 | 1698 | 0.84 (0.77, 0.90) | 42.6 (2.0) | 58.4 | 439481 | 0.87 (0.80, 0.94) | 56.8 (8.0) | 54.1 |
| SBP (mmHg) | 3758 | 128.4 (16.0) | 43.1 (2.3) | 61.1 | 1700 | 128.2 (15.7) | 42.6 (2.0) | 58.4 | 439871 | 141.3 (20.6) | 56.8 (8.0) | 54.1 |
| DBP (mmHg) | 3758 | 80.7 (10.7) | 43.1 (2.3) | 61.1 | 1700 | 80.6 (10.7) | 42.6 (2.0) | 58.4 | 439914 | 84.3 (11.2) | 56.8 (8.0) | 54.1 |
| Glucose (mmol/L) | 3233 | 4.77 (4.47, 5.10) | 43.1 (2.3) | 60.6 | 1481 | 4.81 (4.50, 5.13) | 42.6 (2.0) | 57.7 | 380633 | 4.93 (4.60, 5.30) | 56.8 (8.0) | 53.9 |
| HbA1c (mmol/mol) | 3587 | 33.1 (30.8, 35.4) | 43.1 (2.3) | 61.3 | 1624 | 33.2 (31.0, 35.2) | 42.6 (2.0) | 59.1 | 416643 | 35.1 (32.7, 37.7) | 56.8 (8.0) | 54.2 |
| TC (mmol/L) | 3583 | 5.47 (1.00) | 43.1 (2.3) | 61.0 | 1625 | 5.43 (1.01) | 42.6 (2.0) | 58.5 | 419577 | 5.92 (1.09) | 56.8 (8.0) | 54.1 |
| HDL-C (mmol/L) | 3257 | 1.36 (1.15, 1.62) | 43.1 (2.3) | 60.5 | 1482 | 1.35 (1.14, 1.59) | 42.6 (2.0) | 57.6 | 384017 | 1.40 (1.17, 1.68) | 56.8 (8.0) | 53.7 |
| LDL-C (mmol/L) | 3576 | 3.43 (0.79) | 43.1 (2.3) | 61.0 | 1625 | 3.40 (0.81) | 42.6 (2.0) | 58.5 | 418612 | 3.78 (0.85) | 56.8 (8.0) | 54.1 |
| TG (mmol/L) | 3568 | 1.26 (0.87, 1.93) | 43.1 (2.3) | 61.3 | 1619 | 1.28 (0.86, 1.96) | 42.6 (2.0) | 58.7 | 417559 | 1.55 (1.08, 2.26) | 56.8 (8.0) | 54.2 |
| CRP (mg/L)<sup>b</sup> | 3563 | 1.07 (0.52, 2.28) | 43.1 (2.3) | 61.0 | 1613 | 1.01 (0.52, 2.18) | 42.5 (2.0) | 58.6 | 415269 | 1.32 (0.65, 2.70) | 56.8 (8.0) | 54.1 |
| Birthweight (kg)<sup>c</sup> | 3061 | 3.320 (0.542) | NA | 64.7 | 1238 | 3.318 (0.529) | NA | 62.4 | 250042 | 3.324 (0.665) | NA | 60.8 |
| **ALSPAC** |  |  |  |  |  |  |  |  |  |  |  |  |
| BMI (kg/m<sup>2</sup>) | 2587 | 23.7 (21.5, 27.0) | 24.5 (0.8) | 61.2 | 1043 | 23.5 (21.4, 26.5) | 24.5 (0.8) | 54.7 | 2767 | 23.8 (21.5, 27.0) | 24.5 (0.8) | 60.9 |
| WHR (cm/cm) | 2583 | 0.79 (0.75, 0.84) | 24.5 (0.8) | 61.3 | 1041 | 0.80 (0.75, 0.84) | 24.5 (0.8) | 54.7 | 2763 | 0.79 (0.75, 0.85) | 24.5 (0.8) | 60.9 |
| SBP (mmHg) | 2601 | 116.0 (11.3) | 24.5 (0.8) | 61.3 | 1049 | 116.6 (11.8) | 24.5 (0.8) | 54.9 | 2785 | 116.1 (11.4) | 24.5 (0.8) | 60.9 |
| DBP (mmHg) | 2602 | 66.8 (7.8) | 24.5 (0.8) | 61.3 | 1048 | 66.4 (7.9) | 24.5 (0.8) | 54.9 | 2784 | 66.7 (7.9) | 24.5 (0.8) | 60.9 |
| Glucose (mmol/L) | 2126 | 5.25 (4.95, 5.59) | 24.5 (0.8) | 59.4 | 892 | 5.25 (4.95, 5.60) | 24.5 (0.8) | 52.5 | 2320 | 5.26 (4.95, 5.59) | 24.5 (0.8) | 58.6 |
| TC (mmol/L) | 2135 | 4.44 (0.84) | 24.5 (0.8) | 59.3 | 894 | 4.44 (0.85) | 24.5 (0.8) | 52.5 | 2326 | 4.44 (0.83) | 24.5 (0.8) | 58.6 |
| HDL-C (mmol/L) | 2135 | 1.52 (1.26, 1.80) | 24.5 (0.8) | 59.3 | 894 | 1.52 (1.26, 1.81) | 24.5 (0.8) | 52.5 | 2326 | 1.53 (1.26, 1.80) | 24.5 (0.8) | 58.6 |
| LDL-C (mmol/L) | 2134 | 2.44 (0.76) | 24.5 (0.8) | 59.4 | 892 | 2.42 (0.78) | 24.5 (0.8) | 52.6 | 2324 | 2.44 (0.75) | 24.5 (0.8) | 58.6 |
| TG (mmol/L) | 2121 | 0.84 (0.66, 1.15) | 24.5 (0.8) | 59.6 | 887 | 0.84 (0.66, 1.15) | 24.5 (0.8) | 52.9 | 2312 | 0.84 (0.66, 1.14) | 24.5 (0.8) | 58.9 |
| CRP (mg/L)<sup>b</sup> | 1973 | 0.80 (0.38, 2.08) | 24.5 (0.8) | 60.1 | 829 | 0.75 (0.36, 1.96) | 24.5 (0.8) | 53.4 | 2152 | 0.80 (0.39, 2.10) | 24.5 (0.8) | 59.1 |
| Birthweight (kg)<sup>c</sup> | 7361 | 3.416 (0.547) | NA | 49.5 | 1545 | 3.443 (0.537) | NA | 46.0 | 7363 | 3.437 (0.533) | NA | 48.5 |

HUNT (Trøndelag Health Study), UKB (UK Biobank), ALSPAC (Avon Longitudinal Study of Parents and Children), BMI (body mass index), WHR (waist-hip ratio), SBP (systolic blood pressure), DBP (diastolic blood pressure), HbA1c (glycated hemoglobin), TC (total cholesterol), HDL-C (high density lipoprotein cholesterol), LDL-C (low density lipoprotein cholesterol), TG (triglycerides), CRP (C-reactive protein).
<sup>a</sup>-mean (standard deviation) or median (interquartile range) is presented for normal and non-normal variables, respectively, <sup>b</sup>-represents offspring outcomes which were natural log transformed for the main analyses, therefore, *n* may differ slightly in Table 1 than GWAS analyses, <sup>c</sup>-publicly available maternal and offspring birthweight GWAS data from EGG (early growth genetics) already included UKB and ALSPAC participants, therefore analyses were not rerun for these samples

### Intergenerational MR of maternal glycemic traits on offspring cardiometabolic risk factors and birthweight

We observed that MR estimates from the duos WLM (maternal and offspring samples or paternal and offspring samples) had higher statistical power (reduced standard errors) than MR estimates from the trios WLM (**Supplementary information S11**). Therefore, we present the MR estimates from the duos WLM as the main result (**Figure 3**, **Figure 4**). This is appropriate as i) we observed that the MR point estimates from the duos WLM did not substantially differ from the trios WLM, and ii) the paternal MR estimates from the trios WLM were null (**Supplementary information S11**).

**Figure 3.**
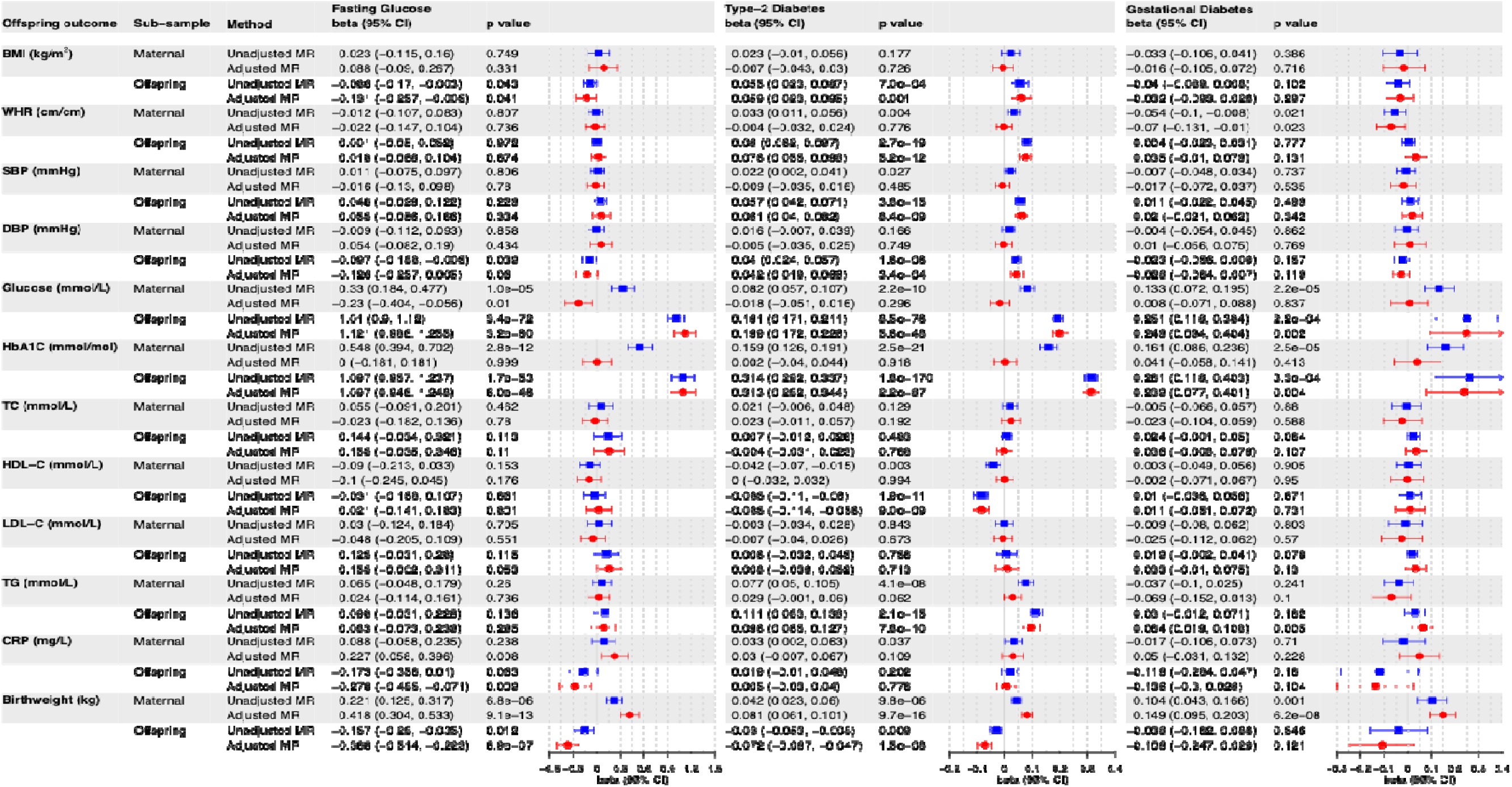
Unadjusted and adjusted intergenerational MR of maternal glycemic traits and offspring cardiometabolic risk factors in adulthood and birthweight.

**Figure 4.**
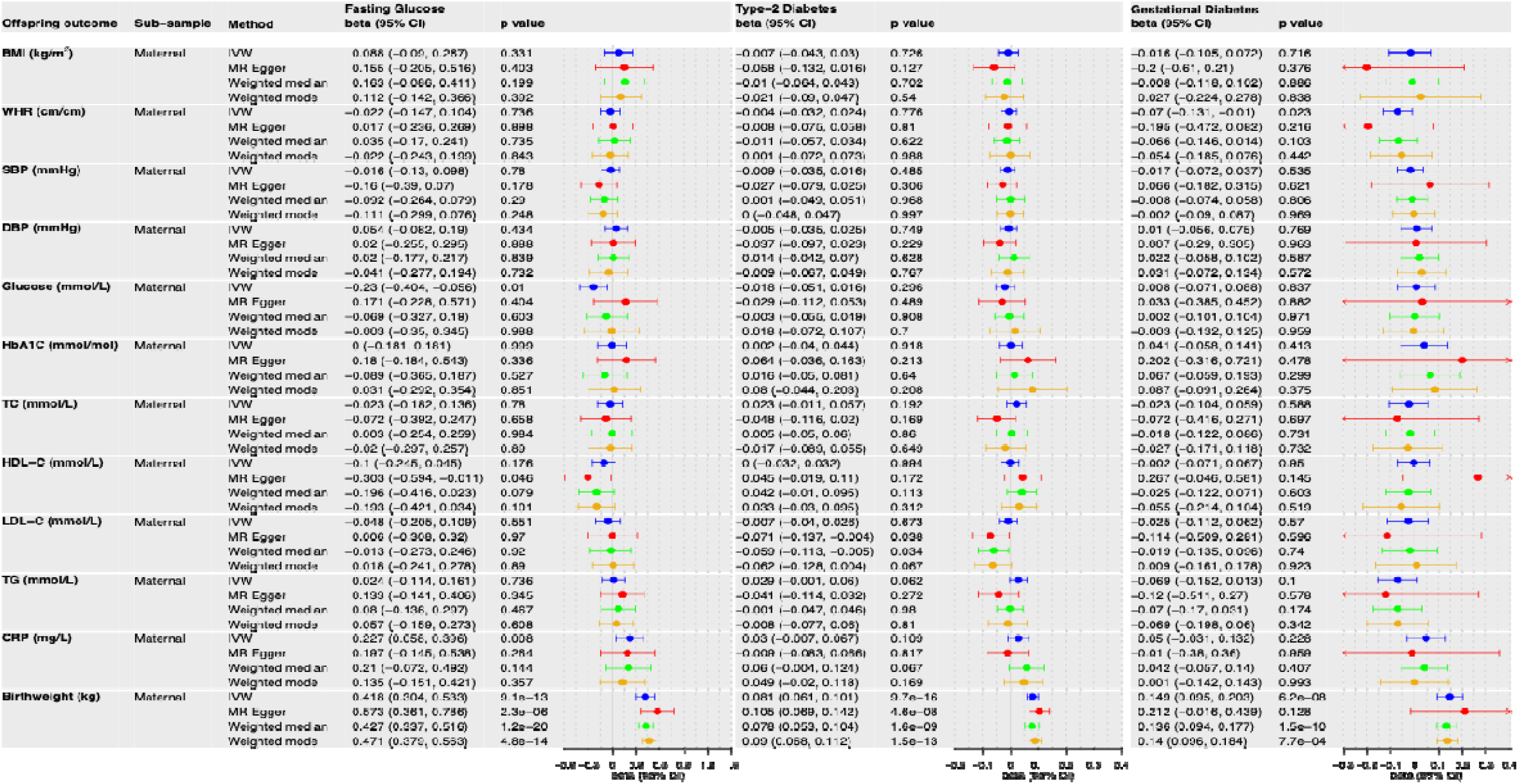
Sensitivity analysis for adjusted intergenerational MR of maternal glycemic traits and offspring cardiometabolic risk factors in adulthood and birthweight.

We observed no strong evidence for a causal effect of maternal glycemic traits on offspring cardiometabolic risk factors in adulthood, after adjusting for offspring genotype (**Figure 3**). When offspring genotype was not adjusted for, maternal glycemic traits appeared to causally increase offspring blood glucose (unadjusted estimates: maternal fasting glucose IVW_blood glucose_ estimate=0.330, 95% CI 0.184 to 0.477, *P*=1.0x10^-5^) and HbA1c (unadjusted estimates: maternal fasting glucose IVW_HbA1c_ estimate=0.548, 95% CI 0.394 to 0.702, *P*=2.8x10^-12^) in adulthood, but the increase was attenuated to the null after adjusting for offspring genotype (adjusted estimates: maternal fasting glucose IVW_blood glucose_ estimate=−0.230, 95% CI −0.404 to −0.056, *P*=0.010; maternal fasting glucose IVW_HbA1c_ estimate=−0.0003, 95% CI −0.181 to 0.181, *P*=0.999). Similar results were observed for maternal type 2 diabetes and gestational diabetes. This attenuation may reflect violation of the exclusion restriction MR assumption due to genetic inheritance.

In the positive control analysis, we observed that maternal glycemic traits increased offspring birthweight (adjusted estimates: maternal fasting glucose IVW_birthweight_ estimate=0.418, 95% CI 0.304 to 0.533, *P*=9.1x10^-13^) but the inherited risk alleles of glycemic traits in offspring tends to decrease the offspring birthweight (adjusted estimates: offspring fasting glucose IVW_birthweight_ estimate=−0.368, 95% CI −0.514 to −0.223, *P*=6.9x10^-7^). Similar results were observed for maternal type 2 diabetes and gestational diabetes (**Figure 3**).

In the negative control analysis, we observed that paternal glycemic traits were not causally associated with any offspring cardiometabolic risk factors in adulthood (**Supplementary information S11**).

The F-statistics used to test the strength of SNPs of glycemic traits used for MR analyses, were generally high (mean F-statistic >41) (**Supplementary information S12**).

In sensitivity analyses, results from other MR methods such as MR Egger, Weighted median, and Weighted mode were not materially different from the IVW estimates (**Figure 4**, **Supplementary information S13**). No strong evidence of bias due to horizontal pleiotropy was indicated by the MR Egger intercept, Cochran’s Q test, and MR-PRESSO global test (**Supplementary information S14**).

## Discussion

In this intergenerational MR analyses of up to 566,132 individuals from the HUNT, UKB, and ALSPAC cohorts, higher maternal glucose, type 2 diabetes, and gestational diabetes had no strong causal effect on offspring cardiometabolic risk factors in adulthood. We observed an apparent effect on offspring glycemic traits that attenuated to the null with adjustment for offspring and paternal genotype. Furthermore, our finding that higher maternal levels of fasting glucose and type 2/gestational diabetes resulted in higher offspring birth weight (our positive control) supports the validity of our main findings.

Observational studies^12^ ^13^ ^65–68^ have suggested that maternal glucose levels during pregnancy may cause a poor offspring cardiometabolic risk factor profile in childhood and early adulthood. However, in our MR analyses we observed no strong evidence of causal effect of maternal glycemic traits on offspring BMI, blood glucose, HbA1c, and other cardiometabolic risk factors, suggesting that these observed relationships might be due to confounding. Importantly, we observed a putative causal effect of maternal glycemic traits on offspring blood glucose and HbA1c in our unadjusted MR analyses which did not control for genetic inheritance. However, when we adjusted for offspring genotype in our adjusted MR analyses, the causal effects were nullified, strongly suggesting that the unadjusted MR estimates are biased by genetic inheritance. Prior work has supported the importance of genetic confounding as a driver of parent-offspring cardiometabolic risk factor association^11^^69^. Hence, our results suggest that previous observational findings might have meaningful confounding issues illustrating the need for intergenerational MR approaches that appropriately account for genetic confounding. In a companion paper, we found no strong evidence for causal effects of parental BMI on the same offspring cardiovascular factors^55^. Together with the present analyses, this suggests that important exposures relating to fetal overnutrition may not have large effects on offspring cardiovascular risk factors in adulthood. However, we acknowledge large studies with participant follow-up into older ages are important to rule out effects on cardiovascular diseases such as coronary heart disease and stroke.

Similar to previous studies^17^ ^18^, when we accounted for confounding (including via genetic inheritance) using an intergenerational MR approach, we observed that maternal glycemic traits increased offspring birthweight, indicating intrauterine environmental effects of maternal glucose on fetal overgrowth. We also observed that the direct (fetal) effect of the inherited glycemic alleles on birthweight was negative. This is in line with the fetal insulin hypothesis (FIH)^70^, which postulates that alleles which cause reduced insulin production or sensitivity predispose the offspring to lower intrauterine growth (since insulin is a fetal growth factor), and subsequently also to type 2 diabetes in adulthood.

In contrast to our study, two studies^12^ ^13^ found a within-sibling association between maternal diabetes with higher BMI and type 2 diabetes in the offspring at ages 2-24 years. Such sibling designs should avoid bias due to genetic confounding, but might be affected by selection bias or bias due to carryover – i.e. where the exposure or outcome in one pregnancy/sibling influences the exposure or outcome in the subsequent sibling^14^ ^15^. Such carryover is plausible and could result in the observed results. For example, a pregnancy resulting in an infant born large for gestational age will lead to women in subsequent pregnancies having a diagnostic test for gestational diabetes. That said, our MR results reflect the potential effect of genetic liability to glycemic traits, which is not the same as a diagnosis. Such liability would be directly carried over to subsequent pregnancies but not always the diagnosis. Thus, a direct comparison between the two designs is challenging. Additionally, although not directly comparable to our examined glycemic traits, a study^71^ leveraging United Kingdom’s sugar rationing until 1953 in a quasi-experimental design, suggested that sugar rationing *in utero* and in infancy reduced the risk of type 2 diabetes and hypertension in adulthood.

### Strengths and limitations

Our study involves the largest intergenerational MR of offspring cardiometabolic risk factors in adulthood to date. The combination of large-scale family-based datasets, combined with recently established statistical methods^39^ ^50–52^ allowed us to expand our total sample size beyond the subset of individuals with complete maternal, paternal, and offspring genotype data, dramatically increasing statistical efficiency. Further to this, offspring in HUNT and UKB are of an age (>40 years) where one would expect cardiometabolic risk factors to deteriorate if lifelong effects had accumulated since conception. Our intergenerational MR method facilitates the application of a two-sample MR approach, enabling sensitivity analyses to test for bias due to horizontal pleiotropy. We have made the summary statistics from our adjusted GWAS publicly available, enabling researchers to investigate causal effects of numerous other parental exposures on offspring cardiometabolic risk factors in adulthood.

Our study is limited by the genetic instruments available for each exposure (glycemic traits). The genetic instruments for fasting glucose and type 2 diabetes have been identified over repeated rounds of meta-analysis, while GWAS of gestational diabetes have far smaller sample sizes, may identify variants which lack distinction from type 2 diabetes variants. The GWAS summary statistics for fasting glucose were adjusted for BMI, but the authors confirmed in sensitivity analysis that collider bias did not influence locus discovery for more than 98% of discovered SNPs^72^. We also showed that GWAS summary statistics for unadjusted and BMI-adjusted random glucose are extremely similar (regression *R*^2^=0.998) in 370,554 UKB participants (**Supplementary information S15**). Furthermore, we demonstrated that genetic effect estimates for fasting glucose from general population GWAS were similar in pregnant women from the BiB study. Our high-income country, European ancestry specific analyses could limit the generalizability of our findings and follow up studies involving participants from other ancestries and low/middle income countries would be highly beneficial. Our available sample size was insufficient to perform the age-stratified analyses to test life-course effects. Our GWAS analysis assumes that the cross-locus genetic correlations due to assortative mating were null but any bias due to such cross-locus assortative mating effects may be small especially for biological traits (such as glucose levels) rather than social traits. The WLM method for our GWAS analysis could i) produce biased adjusted estimates in the presence of genotyping error or poor imputation as these could lead to attenuation bias (however, our GWAS summary statistics for MR analysis were filtered on imputation quality score ≥0.80), ii) assumes no sex heterogeneity in direct genetic effects, and iii) works best when maternal, paternal, and offspring GWAS are of similar size/precision of estimates (although we confirmed via simulation that adjusted WLM estimates are unbiased, even when the sample sizes for maternal, paternal, and offspring GWAS are drastically different). Furthermore, our method was not able to directly disentangle the intrauterine effect from potential postnatal environmental (early life) effects^1^ ^65^ ^67^ ^73^; however, in our analysis, the small maternal MR estimates, and positive (birthweight) and negative control (paternal effect) estimates suggest neither of these mechanisms had strong effects on offspring cardiometabolic risk factors in adulthood. Our paternal analysis however provides some insight on the possible effect of the intrauterine environment because if there is postnatal effect, we might expect to observe it in both parents.

## Conclusions

Our intergenerational MR analysis suggests that intrauterine exposure to higher maternal glucose levels does not have a major influence on offspring cardiometabolic risk factors in adulthood up to mid-life. If replicated, particularly in larger samples with disease outcomes, these findings argue against the previously hypothesized intergenerational vicious cycle of cardiometabolic disease.

## Author Contributions

Conceptualization: LB, TAB, DME, BOA, GHM, BMB. Study design: LB, TAB, NMW, DME, GHM, BMB. Software/coding: LB, TAB, GW. Formal analysis: LB, TAB. Data curation: LB, TAB, DAL, BOA, BMB. Writing-original draft preparation: LB, TAB, BOA, GHM, BMB. Writing-review and editing: LB, TAB, NMW, GW, MD, DME, DAL, BOA, GHM, BMB. LB and TAB combined had full access to all the data in the study and take responsibility for the integrity of the data and the accuracy of the data analysis. This publication is the work of the authors, all authors read and approved the final manuscript, and LB, TAB, BOA, GHM, BMB will serve as guarantors for the contents of this paper.

## Data Sharing

The analyses used individual level data from HUNT, UKB, ALSPAC, and BiB. These data are available on application to the corresponding cohorts and requirements for accessing data are described at www.ntnu.edu/hunt/, https://www.ukbiobank.ac.uk/, http://www.bristol.ac.uk/alspac/researchers/our-data, and https://borninbradford.nhs.uk/, respectively. The birthweight data used in HUNT are available via linking to Medical Birth Registry of Norway managed by National institute of Public Health (https://www.fhi.no/en/ch/medical-birth-registry-of-norway/). Publicly available population GWAS summary statistics of fasting glucose, type 2 diabetes, and gestational diabetes are available via https://www.nature.com/articles/s41588-021-00852-9, https://www.nature.com/articles/s41588-020-0637-y, and https://www.nature.com/articles/s41588-023-01607-4, respectively. The publicly available GWAS summary statistics of birthweight are available via EGG consortium website www.egg-consortium.org and deCODE genetics website https://www.decode.com/summarydata/. The GWAS summary statistics of our analyses are freely available on publication via HUNT MCE – “Within Family” website https://www.ntnu.edu/web/hunt/mce/family. The summary data to reproduce the MR analyses reported in this paper are available in **Supplementary table S1** and required R code are publicly available at https://github.com/laxmibhatta/Intergenerational_MR_analysis/.

## Data Availability

The analyses used individual level data from HUNT, UKB, ALSPAC, and BiB. These data are available on application to the corresponding cohorts and requirements for accessing data are described at www.ntnu.edu/hunt/, https://www.ukbiobank.ac.uk/, http://www.bristol.ac.uk/alspac/researchers/our-data, and https://borninbradford.nhs.uk/, respectively. The birthweight data used in HUNT are available via linking to Medical Birth Registry of Norway managed by National institute of Public Health (https://www.fhi.no/en/ch/medical-birth-registry-of-norway/). Publicly available population GWAS summary statistics of fasting glucose, type 2 diabetes, and gestational diabetes are available via https://www.nature.com/articles/s41588-021-00852-9, https://www.nature.com/articles/s41588-020-0637-y, and https://www.nature.com/articles/s41588-023-01607-4, respectively. The publicly available GWAS summary statistics of birthweight are available via EGG consortium website www.egg-consortium.org and deCODE genetics website https://www.decode.com/summarydata/. The GWAS summary statistics of our analyses are freely available on publication via HUNT MCE Within Family website https://www.ntnu.edu/web/hunt/mce/family. The summary data needed to reproduce the MR analyses reported in this paper are available in Supplementary table S1.

## Acknowledgements

The authors would like to thank the research participants of the HUNT study which is a collaboration between HUNT Research Centre (Faculty of Medicine and Health Sciences, NTNU Norwegian University of Science and Technology), Trøndelag County Council, Central Norway Regional Health Authority, and the Norwegian Institute of Public Health. The HUNT genotype quality control and imputation has been conducted by the K.G. Jebsen Center for Genetic Epidemiology, Department of Public Health and Nursing, Faculty of Medicine and Health Sciences, NTNU.

This research has been conducted using the UK Biobank resource (Reference 53641).

We would like to thank all the families who took part in ALSPAC, the midwives for their help in recruiting them, and the whole ALSPAC team.

This study used data from Born in Bradford (BiB) cohort and would like to thank participating families and the whole BiB team.

This study used data from Medical Birth Registry of Norway (MBRN).

Furthermore, this study used publicly available GWAS summary statistics contributed by MAGIC Consortium (https://www.magicinvestigators.org/), FinnGen (www.finbb.fi), EGG consortium (http://egg-consortium.org/), and deCODE genetics (https://www.decode.com/summarydata/).

This work used the University of Bristol’s ACRC (Advanced Computing Research Centre) supercomputing facilities.

## References

1. Barker DJ. The fetal and infant origins of adult disease. British Medical Journal 1990;301(6761):1111. doi: 10.1136/bmj.301.6761.1111

2. Castillo-Castrejon M, Powell TL. Placental Nutrient Transport in Gestational Diabetic Pregnancies. Front Endocrinol (Lausanne) 2017;8:306. doi: 10.3389/fendo.2017.00306 [published Online First: 20171107]

3. Plows JF, Stanley JL, Baker PN, et al. The Pathophysiology of Gestational Diabetes Mellitus. Int J Mol Sci 2018;19(11) doi: 10.3390/ijms19113342 [published Online First: 20181026]

4. Tam WH, Ma RCW, Ozaki R, et al. In Utero Exposure to Maternal Hyperglycemia Increases Childhood Cardiometabolic Risk in Offspring. Diabetes Care 2017;40(5):679–86. doi: 10.2337/dc16-2397 [published Online First: 20170309]

5. Lawlor DA. The Society for Social Medicine John Pemberton Lecture 2011. Developmental overnutrition--an old hypothesis with new importance? International journal of epidemiology 2013;42(1):7–29. doi: 10.1093/ije/dys209

6. Pettitt DJ, Knowler WC. Diabetes and obesity in the Pima Indians: a cross-generational vicious cycle. Journal of obesity and weight regulation 1988

7. Gillman MW, Rifas-Shiman S, Berkey CS, et al. Maternal Gestational Diabetes, Birth Weight, and Adolescent Obesity. Pediatrics 2003;111(3):e221–e26. doi: 10.1542/peds.111.3.e221

8. Brumpton B, Sanderson E, Heilbron K, et al. Avoiding dynastic, assortative mating, and population stratification biases in Mendelian randomization through within-family analyses. Nature Communications 2020;11(1):3519. doi: 10.1038/s41467-020-17117-4

9. Davies NM, Howe LJ, Brumpton B, et al. Within family Mendelian randomization studies. Hum Mol Genet 2019;28(R2):R170–r79. doi: 10.1093/hmg/ddz204 [published Online First: 2019/10/28]

10. Smith GD, Ebrahim S. Mendelian randomization: prospects, potentials, and limitations. International journal of epidemiology 2004;33(1):30–42. doi: 10.1093/ije/dyh132

11. Wang G, Bhatta L, Moen GH, et al. Investigating a Potential Causal Relationship Between Maternal Blood Pressure During Pregnancy and Future Offspring Cardiometabolic Health. Hypertension 2022;79(1):170–77. doi: 10.1161/hypertensionaha.121.17701 [published Online First: 20211117]

12. Dabelea D, Hanson RL, Lindsay RS, et al. Intrauterine exposure to diabetes conveys risks for type 2 diabetes and obesity: a study of discordant sibships. Diabetes 2000;49(12):2208–11. doi: 10.2337/diabetes.49.12.2208

13. Lawlor DA, Lichtenstein P, Långström N. Association of Maternal Diabetes Mellitus in Pregnancy With Offspring Adiposity Into Early Adulthood. Circulation 2011;123(3):258–65. doi: 10.1161/CIRCULATIONAHA.110.980169

14. Westvik-Johari K, Håberg SE, Lawlor DA, et al. The challenges of selective fertility and carryover effects in within-sibship analyses: the effect of assisted reproductive technology on perinatal mortality as an example. International journal of epidemiology 2023;52(2):403–13. doi: 10.1093/ije/dyad003

15. Sjölander A, Frisell T, Kuja-Halkola R, et al. Carryover Effects in Sibling Comparison Designs. Epidemiology 2016;27(6):852–8. doi: 10.1097/ede.0000000000000541

16. Frisell T. Invited Commentary: Sibling-Comparison Designs, Are They Worth the Effort? American Journal of Epidemiology 2021;190(5):738–41. doi: 10.1093/aje/kwaa183

17. Chen J, Bacelis J, Sole-Navais P, et al. Dissecting maternal and fetal genetic effects underlying the associations between maternal phenotypes, birth outcomes, and adult phenotypes: A mendelian-randomization and haplotype-based genetic score analysis in 10,734 mother–infant pairs. PLOS Medicine 2020;17(8):e1003305. doi: 10.1371/journal.pmed.1003305

18. Tyrrell J, Richmond RC, Palmer TM, et al. Genetic Evidence for Causal Relationships Between Maternal Obesity-Related Traits and Birth Weight. Jama 2016;315(11):1129–40. doi: 10.1001/jama.2016.1975

19. Evans DM, Moen G-H, Hwang L-D, et al. Elucidating the role of maternal environmental exposures on offspring health and disease using two-sample Mendelian randomization. International journal of epidemiology 2019;48(3):861–75. doi: 10.1093/ije/dyz019

20. Skrivankova VW, Richmond RC, Woolf BAR, et al. Strengthening the Reporting of Observational Studies in Epidemiology Using Mendelian Randomization: The STROBE-MR Statement. JAMA 2021;326(16):1614–21. doi: 10.1001/jama.2021.18236

21. Lawlor D, Richmond R, Warrington N, et al. Using Mendelian randomization to determine causal effects of maternal pregnancy (intrauterine) exposures on offspring outcomes: Sources of bias and methods for assessing them. Wellcome Open Res 2017;2:11. doi: 10.12688/wellcomeopenres.10567.1 [published Online First: 20170214]

22. Chen J, Spracklen CN, Marenne G, et al. The trans-ancestral genomic architecture of glycemic traits. Nature Genetics 2021;53(6):840–60. doi: 10.1038/s41588-021-00852-9

23. Vujkovic M, Keaton JM, Lynch JA, et al. Discovery of 318 new risk loci for type 2 diabetes and related vascular outcomes among 1.4 million participants in a multi-ancestry meta-analysis. Nature Genetics 2020;52(7):680–91. doi: 10.1038/s41588-020-0637-y

24. Elliott A, Walters RK, Pirinen M, et al. Distinct and shared genetic architectures of gestational diabetes mellitus and type 2 diabetes. Nature Genetics 2024;56(3):377–82. doi: 10.1038/s41588-023-01607-4

25. Wright J, Small N, Raynor P, et al. Cohort Profile: The Born in Bradford multi-ethnic family cohort study. International journal of epidemiology 2013;42(4):978–91. doi: 10.1093/ije/dys112

26. Åsvold BO, Langhammer A, Rehn TA, et al. Cohort Profile Update: The HUNT Study, Norway. International journal of epidemiology 2022 doi: 10.1093/ije/dyac095 [published Online First: 20220517]

27. Brumpton BM, Graham S, Surakka I, et al. The HUNT study: A population-based cohort for genetic research. Cell Genomics 2022;2(10):100193. doi: 10.1016/j.xgen.2022.100193

28. Krokstad S, Langhammer A, Hveem K, et al. Cohort Profile: The HUNT Study, Norway. International journal of epidemiology 2012 9 August 2012. http://www.ncbi.nlm.nih.gov/pubmed/22879362 http://ije.oxfordjournals.org/content/early/2012/08/09/ije.dys095.full.pdf (accessed Aug 9).

29. Næss M, Kvaløy K, Sørgjerd EP, et al. Data Resource Profile: The HUNT Biobank. International journal of epidemiology 2024;53(3):dyae073. doi: 10.1093/ije/dyae073

30. Norwegian Institute of Public Health. Medical Birth Registry of Norway Available at: www.fhi.no/en/hn/health-registries/medical-birth-registry-of-norway/medical-birth-registry-of-norway.

31. Moth FN, Sebastian TR, Horn J, et al. Validity of a selection of pregnancy complications in the Medical Birth Registry of Norway. Acta Obstet Gynecol Scand 2016;95(5):519–27. doi: 10.1111/aogs.12868 [published Online First: 20160309]

32. UK Biobank. 2006-2010, https://www.ukbiobank.ac.uk/, Accessed 01 February 2019.

33. Bycroft C, Freeman C, Petkova D, et al. The UK Biobank resource with deep phenotyping and genomic data. Nature 2018;562(7726):203–09. doi: 10.1038/s41586-018-0579-z [published Online First: 20181010]

34. Fraser A, Macdonald-Wallis C, Tilling K, et al. Cohort Profile: the Avon Longitudinal Study of Parents and Children: ALSPAC mothers cohort. International journal of epidemiology 2013;42(1):97–110. doi: 10.1093/ije/dys066 [published Online First: 2012/04/16]

35. Boyd A, Golding J, Macleod J, et al. Cohort Profile: the ‘children of the 90s’--the index offspring of the Avon Longitudinal Study of Parents and Children. International journal of epidemiology 2013;42(1):111–27. doi: 10.1093/ije/dys064 [published Online First: 20120416]

36. Northstone K, Lewcock M, Groom A, et al. The Avon Longitudinal Study of Parents and Children (ALSPAC): an update on the enrolled sample of index children in 2019. Wellcome Open Res 2019;4:51. doi: 10.12688/wellcomeopenres.15132.1 [published Online First: 20190314]

37. Northstone K, Ben Shlomo Y, Teyhan A, et al. The Avon Longitudinal Study of Parents and children ALSPAC G0 Partners: A cohort profile [version 2; peer review: 1 approved]. Wellcome Open Research 2023;8(37) doi: 10.12688/wellcomeopenres.18782.2

38. Taylor AE, Jones HJ, Sallis H, et al. Exploring the association of genetic factors with participation in the Avon Longitudinal Study of Parents and Children. International journal of epidemiology 2018;47(4):1207–16. doi: 10.1093/ije/dyy060

39. Warrington NM, Beaumont RN, Horikoshi M, et al. Maternal and fetal genetic effects on birth weight and their relevance to cardio-metabolic risk factors. Nature Genetics 2019;51(5):804–14. doi: 10.1038/s41588-019-0403-1

40. Juliusdottir T, Steinthorsdottir V, Stefansdottir L, et al. Distinction between the effects of parental and fetal genomes on fetal growth. Nat Genet 2021;53(8):1135–42. doi: 10.1038/s41588-021-00896-x [published Online First: 20210719]

41. Moen GH, Brumpton B, Willer C, et al. Mendelian randomization study of maternal influences on birthweight and future cardiometabolic risk in the HUNT cohort. Nat Commun 2020;11(1):5404. doi: 10.1038/s41467-020-19257-z [published Online First: 20201026]

42. Manichaikul A, Mychaleckyj JC, Rich SS, et al. Robust relationship inference in genome-wide association studies. Bioinformatics 2010;26(22):2867–73. doi: 10.1093/bioinformatics/btq559 [published Online First: 20101005]

43. Jiang L, Zheng Z, Qi T, et al. A resource-efficient tool for mixed model association analysis of large-scale data. Nature Genetics 2019;51(12):1749–55. doi: 10.1038/s41588-019-0530-8

44. Yang J, Lee SH, Goddard ME, Visscher PM. GCTA: a tool for genome-wide complex trait analysis. American journal of human genetics 2011;88(1):76–82. doi: 10.1016/j.ajhg.2010.11.011 [published Online First: 2010/12/17]

45. Purcell S, Neale B, Todd-Brown K, et al. PLINK: a tool set for whole-genome association and population-based linkage analyses. American journal of human genetics 2007;81(3):559–75. doi: 10.1086/519795 [published Online First: 20070725]

46. Willer CJ, Li Y, Abecasis GR. METAL: fast and efficient meta-analysis of genomewide association scans. Bioinformatics 2010;26(17):2190–91. doi: 10.1093/bioinformatics/btq340

47. Ani A, van der Most PJ, Snieder H, et al. GWASinspector: comprehensive quality control of genome-wide association study results. Bioinformatics 2021;37(1):129–30. doi: 10.1093/bioinformatics/btaa1084

48. Bulik-Sullivan BK, Loh PR, Finucane HK, et al. LD Score regression distinguishes confounding from polygenicity in genome-wide association studies. Nat Genet 2015;47(3):291–5. doi: 10.1038/ng.3211 [published Online First: 20150202]

49. Loh P-R, Kichaev G, Gazal S, et al. Mixed-model association for biobank-scale datasets. Nature Genetics 2018;50(7):906–08. doi: 10.1038/s41588-018-0144-6

50. Warrington NM, Hwang LD, Nivard MG, Evans DM. Estimating direct and indirect genetic effects on offspring phenotypes using genome-wide summary results data. Nat Commun 2021;12(1):5420. doi: 10.1038/s41467-021-25723-z [published Online First: 20210914]

51. Beaumont RN, Flatley C, Vaudel M, et al. Genome-wide association study of placental weight identifies distinct and shared genetic influences between placental and fetal growth. Nat Genet 2023;55(11):1807–19. doi: 10.1038/s41588-023-01520-w [published Online First: 20231005]

52. Wu Y, Zhong X, Lin Y, et al. Estimating genetic nurture with summary statistics of multigenerational genome-wide association studies. Proc Natl Acad Sci U S A 2021;118(25) doi: 10.1073/pnas.2023184118

53. Magnus P, Birke C, Vejrup K, et al. Cohort Profile Update: The Norwegian Mother and Child Cohort Study (MoBa). International journal of epidemiology 2016;45(2):382–8. doi: 10.1093/ije/dyw029 [published Online First: 20160410]

54. Elizabeth CC, Oleksandr F, Alexey AS, et al. The Norwegian Mother, Father, and Child cohort study (MoBa) genotyping data resource: MoBaPsychGen pipeline v.1. bioRxiv 2022:2022.06.23.496289. doi: 10.1101/2022.06.23.496289

55. Bond TA, Bhatta L, Yang Q, Moen GH, Wang G, Beamont RN, Morris TT, Hwang LD, Wootton RE, Corfield EC, Warrington NM, Magnus MC, Havdahl A, Borges MC, Lawlor DA, Åsvold BO, Brumpton BM, Evans DM. Parental body mass index and offspring cardiovascular risk factors in adulthood: an intergenerational Mendelian randomization study. medRxiv 2025.

56. Hemani G, Zheng J, Elsworth B, et al. The MR-Base platform supports systematic causal inference across the human phenome. Elife 2018;7 doi: 10.7554/eLife.34408 [published Online First: 2018/05/31]

57. Bowden J, Davey Smith G, Burgess S. Mendelian randomization with invalid instruments: effect estimation and bias detection through Egger regression. International journal of epidemiology 2015;44(2):512–25. doi: 10.1093/ije/dyv080 [published Online First: 2015/06/06]

58. Bowden J, Davey Smith G, Haycock PC, Burgess S. Consistent Estimation in Mendelian Randomization with Some Invalid Instruments Using a Weighted Median Estimator. Genetic epidemiology 2016;40(4):304–14. doi: 10.1002/gepi.21965 [published Online First: 2016/04/07]

59. Hartwig FP, Davey Smith G, Bowden J. Robust inference in summary data Mendelian randomization via the zero modal pleiotropy assumption. International journal of epidemiology 2017;46(6):1985–98. doi: 10.1093/ije/dyx102

60. Burgess S, Thompson SG, Collaboration CCG. Avoiding bias from weak instruments in Mendelian randomization studies. International journal of epidemiology 2011;40(3):755–64. doi: 10.1093/ije/dyr036

61. Verbanck M, Chen C-Y, Neale B, Do R. Detection of widespread horizontal pleiotropy in causal relationships inferred from Mendelian randomization between complex traits and diseases. Nature Genetics 2018;50(5):693–98. doi: 10.1038/s41588-018-0099-7

62. Greco MF, Minelli C, Sheehan NA, Thompson JR. Detecting pleiotropy in Mendelian randomisation studies with summary data and a continuous outcome. Statistics in medicine 2015;34(21):2926–40. doi: 10.1002/sim.6522 [published Online First: 20150507]

63. qctool v2. Available at: https://www.chg.ox.ac.uk/~gav/qctool_v2/.

64. R Core Team. R: A language and environment for statistical computing. Vienna, Austria: R Foundation for Statistical Computing; 2020.

65. Perng W, Ringham BM, Smith HA, et al. A prospective study of associations between in utero exposure to gestational diabetes mellitus and metabolomic profiles during late childhood and adolescence. Diabetologia 2020;63(2):296–312. doi: 10.1007/s00125-019-05036-z

66. Hillier TA, Pedula KL, Schmidt MM, et al. Childhood Obesity and Metabolic Imprinting: The ongoing effects of maternal hyperglycemia. Diabetes Care 2007;30(9):2287–92. doi: 10.2337/dc06-2361

67. Kelstrup L, Damm P, Mathiesen ER, et al. Insulin Resistance and Impaired Pancreatic β-Cell Function in Adult Offspring of Women With Diabetes in Pregnancy. The Journal of Clinical Endocrinology & Metabolism 2013;98(9):3793–801. doi: 10.1210/jc.2013-1536

68. Josefson JL, Catalano PM, Lowe WL, et al. The Joint Associations of Maternal BMI and Glycemia with Childhood Adiposity. J Clin Endocrinol Metab 2020;105(7):2177–88. doi: 10.1210/clinem/dgaa180

69. Bond TA, McAdams TA, Warrington NM, et al. Parental body mass index and offspring childhood body size and eating behaviour: causal inference via parental comparisons and extended children of twins structural equation modelling. medRxiv 2023:2023.02.06.23284912. doi: 10.1101/2023.02.06.23284912

70. Hattersley AT, Tooke JE. The fetal insulin hypothesis: an alternative explanation of the association of low birthweight with diabetes and vascular disease. *Lancet (London*, England) 1999;353(9166):1789–92. doi: 10.1016/s0140-6736(98)07546-1

71. Gracner T, Boone C, Gertler PJ. Exposure to sugar rationing in the first 1000 days of life protected against chronic disease. Science 2024;386(6725):1043–48. doi: 10.1126/science.adn5421

72. Aschard H, Vilhjálmsson BJ, Joshi AD, et al. Adjusting for heritable covariates can bias effect estimates in genome-wide association studies. American journal of human genetics 2015;96(2):329–39. doi: 10.1016/j.ajhg.2014.12.021 [published Online First: 20150129]

73. Bailbe D, Liu J, Gong P, Portha B. Effect of Postnatal Nutritional Environment Due to Maternal Diabetes on Beta Cell Mass Programming and Glucose Intolerance Risk in Male and Female Offspring. Biomolecules 2021;11(2) doi: 10.3390/biom11020179 [published Online First: 20210128]

