## Supplementary information S1-S15 for "Exploring the effect of maternal glycemic traits on offspring cardiometabolic risk factors in adulthood: an intergenerational Mendelian randomization study"

**Supplementary information S1. Publicly available GWAS data used for the exposures**

| Study | Phenotype | Percentage sample overlap for the exposure GWAS | Reference |
|---|---|---|---|
| Chen et. al, 2021[1] | Fasting glucose | 2.2% (ALSPAC) | doi: 10.1038/s41588-021-00852-9 |
| Vujkovic et. al., 2020 [2] | Type 2 diabetes | 65.7% for birthweight as an outcome (DeCODE and UKB) | doi: 10.1038/s41588-020-0637-y |
| Vujkovic et. al., 2020 [2] | Type 2 diabetes | 39.7% for all other outcomes (UKB) | doi: 10.1038/s41588-020-0637-y |
| Elliott et. al., 2024 [3] | Gestational diabetes | No sample overlap | doi: 10.1038/s41588-023-01607-4 |

**Supplementary information S2. Validation of genetic instruments for glucose related phenotypes.**

**Born in Bradford (BiB).** BiB is a multiethnic cohort that included women (n=12,453, >80% of those invited) who were booked for delivery at Bradford Royal Infirmary between 2007 and 2011, who participated in detailed data collection from pregnancy onwards[4]. The cohort includes 13,776 pregnancies and 3,448 of the partners of participating mothers were also recruited. Ethics approval was obtained from Bradford National Health Service Ethics Committee (ref 06/Q1202/48), and all participants provided informed written consent. All women were offered a standard 75g 2 hour oral glucose tolerance test (OGTT) at 26-28 weeks' gestation, which 11,442 women attended. Plasma glucose levels were assayed immediately after sampling at the biochemistry department of Bradford Royal Infirmary using the glucose oxidase method on Siemens Advia 2400 chemistry autoanalysers and Siemens Advia Centaur assay (Camberley, Surrey, UK).

BiB mothers and children were genotyped at Bristol Bioresource Laboratories, Bristol, UK on the Illumina HumanCoreExome array (12v1.0, 12v1.1, or 24v1.0) or Illumina Global Screening Array-24 v1.0 (GSA). Individuals or SNPs with >3% missing genotype calls were dropped for each array separately. 707 individuals were then dropped due to i) mismatch between genetically- and phenotypically-defined sex (110 individuals), ii) suspected sample duplication (127 individuals), and iii) discrepancy between expected and genetically inferred relationship for first degree relatives (470 individuals). A combination of principal component analysis (PCA) and self-reported/primary care record-derived ethnicity data was used to identify participants who were genetically similar to the European or South Asian HapMap samples[5]. Genotype imputation was performed for 7606 European and 8692 South Asian participants, with each ancestry and array (CoreExome/GSA) imputed separately to the HRC r1.1 reference panel[6]. Prior to imputation, variants with call rate <95%, Hardy Weinberg equilibrium (HWE) P-value $<1x10^{-6}$ or MAF <1% were removed for each of the four ancestry/array groups separately. A/T and C/G SNPs were dropped, as were those that failed checks for strand, reference/alternative alleles, position, and SNP duplication, indels, non-autosomal SNPs, SNPs with no match in the HRC panel (position or SNP ID), and SNPs with non-matching alleles.

In this analysis we included European ancestry mothers with both OGTT and quality controlled imputed genotype data available. When mothers participated in BiB for more than one pregnancy we included only their first enrolled pregnancy, giving a final sample size of 3,044 unique mothers.

We extracted independent genetic instruments associated at genome-wide significance with fasting glucose[1], 2-hour glucose[1], fasting insulin[1], glycated hemoglobin (HbA1c)[1], type 2 diabetes (Xue et. al., 2018[7] and Vujkovic et. al., 2020[2]), and gestational diabetes[3] from publicly available GWAS of mixed sex non-pregnant populations. To confirm that these genetic instruments were associated with maternal plasma glucose during pregnancy, we constructed weighted polygenic risk scores (PRS) in BiB, using effect estimates from the corresponding GWAS as weights and the

PLINK2 software package[8]. We then regressed natural log-transformed fasting and 2 hour post-load plasma glucose on each PRS, using a linear mixed model (LMM) implemented in the GCTA software package version 1.94.0beta[9] to control for population stratification and cryptic relatedness. To fit the LMM we used a genetic relatedness matrix (GRM) calculated from common (minor allele frequency ≥1%) called autosomal SNPs, having first dropped the PRS variants, and any variants in linkage disequilibrium with them (r2 = 0.001, kb = 10000), to avoid artificial weakening of PRS-glucose associations by proximal contamination. We estimated the proportion of variance ($R^2$) in fasting and 2 hour post-load plasma glucose explained by each PRS.

Finally, we compared the size of the genetic effect estimates for fasting glucose in the Chen et. al., 2021[1] publicly available GWAS (i.e. a mixed sex non-pregnant population) versus pregnant BiB participants. We conducted GWAS in the same 3,044 BiB participants, using an LMM implemented via the GCTA --mlma command and a GRM as described above. We selected independent variants which were associated with fasting glucose at $P<5x10^{-8}$ in the Chen et. al., 2021[1] GWAS and were available in the quality-controlled BiB genotype data, and we dropped the variant rs12888855 (which had a low minor allele frequency [0.00089] in BiB), giving 66 variants for comparison. We compared effect estimates for individual variants via linear regression, and compared the overall pooled effect size given by conducting a random effects meta-analysis of the 66 variants.

The table below shows the distribution of fasting plasma glucose, 2-hour plasma glucose, gestational age at OGTT, and maternal age at OGTT among 3,044 pregnant BiB participants.

| **Characteristics** | **Min.** | **Q1** | **Mean** | **Median** | **Q3** | **Max.** |
|---|---|---|---|---|---|---|
| Fasting plasma glucose (mmol/L) | 3.0 | 4.1 | 4.4 | 4.3 | 4.6 | 9.4 |
| 2-hour plasma glucose (mmol/L) | 1.6 | 4.6 | 5.5 | 5.3 | 6.2 | 15.5 |
| Gestational age at OGTT (weeks) | 6.0 | 26.0 | 26.2 | 26.0 | 26.0 | 38.0 |
| Maternal age at OGTT (years) | 15.0 | 22.5 | 27.2 | 26.7 | 31.3 | 45.8 |

OGTT (oral glucose tolerance test), Min. (minimum), Q1 (first quartile), Q3 (third quartile), Max. (maximum)

The table below shows the variance of fasting and 2 hour plasma glucose explained by each PRS among 3,044 pregnant BiB participants ($R^2$).

| **PRS (z score)** | **Outcome (mmol/L, natural log, z score)** | **$R^2$ point estimate** | **$R^2$ Lower 95% CL** | **$R^2$ Upper 95% CL** | **Beta** | **SE** | ***P* value** | **No. SNPs** |
|---|---|---|---|---|---|---|---|---|
| Fasting glucose[a] | Fasting plasma glucose | 4.9% | 3.4% | 6.4% | 0.22 | 0.02 | $5.33x10^{-36}$ | 52 |
| Fasting glucose[a] | 2-hour plasma glucose | 0.6% | 0.0% | 1.1% | 0.08 | 0.02 | $2.43x10^{-05}$ | 52 |
| 2hr glucose[a] | Fasting plasma glucose | 0.0% | 0.0% | 0.0% | 0.01 | 0.02 | 0.748287 | 12 |
| 2hr glucose[a] | 2-hour plasma glucose | 0.3% | 0.0% | 0.8% | 0.06 | 0.02 | 0.001244 | 12 |
| Fasting insulin[a] | Fasting plasma glucose | 0.2% | 0.0% | 0.5% | 0.04 | 0.02 | 0.024684 | 28 |
| Fasting insulin[a] | 2-hour plasma glucose | 0.1% | 0.0% | 0.3% | 0.03 | 0.02 | 0.144095 | 28 |
| HbA1c[a] | Fasting plasma glucose | 0.5% | 0.0% | 1.1% | 0.07 | 0.02 | $4.20x10^{-05}$ | 57 |
| HbA1c[a] | 2-hour plasma glucose | 0.1% | 0.0% | 0.4% | 0.04 | 0.02 | 0.044767 | 57 |
| HbA1c (glycaemic SNPs)[a] | Fasting plasma glucose | 1.8% | 0.9% | 2.8% | 0.14 | 0.02 | $5.72x10^{-14}$ | 12 |
| HbA1c (glycaemic SNPs)[a] | 2-hour plasma glucose | 0.7% | 0.1% | 1.3% | 0.08 | 0.02 | $5.01x10^{-06}$ | 12 |

| | | | | | | | | |
|---|---|---|---|---|---|---|---|---|
| T2D (2018)[b] | Fasting plasma glucose | 0.8% | 0.2% | 1.5% | 0.09 | 0.02 | $3.48 \times 10^{-07}$ | 90 |
| T2D (2018)[b] | 2-hour plasma glucose | 0.8% | 0.2% | 1.4% | 0.09 | 0.02 | $1.22 \times 10^{-06}$ | 90 |
| T2D (2020)[c] | Fasting plasma glucose | 1.0% | 0.3% | 1.7% | 0.10 | 0.02 | $3.61 \times 10^{-08}$ | 186 |
| T2D (2020)[c] | 2-hour plasma glucose | 1.1% | 0.4% | 1.8% | 0.11 | 0.02 | $5.61 \times 10^{-09}$ | 186 |
| Gestational diabetes[d] | Fasting plasma glucose | 3.9% | 2.6% | 5.3% | 0.20 | 0.02 | $7.29 \times 10^{-29}$ | 10 |
| Gestational diabetes[d] | 2-hour plasma glucose | 0.6% | 0.0% | 1.1% | 0.08 | 0.02 | $2.18 \times 10^{-05}$ | 10 |

PRS (polygenic risk score), HbA1c (glycated haemoglobin), T2D (type-2 diabetes), $R^2$ (variance explained), CL (confidence limit), SE (standard error), No. SNP (number of single nucleotide polymorphisms). PRS of the genetic instruments was scaled to z-score. The fasting plasma glucose and 2-hour plasma glucose values during pregnancy were natural log transformed. [a]-Chen et. al. 2021[1], [b]-Xue et. al. 2018[7], [c]-Vujkovic et. al 2020[2], [d]-Elliott et. al. 2024[3].

The plot below shows results from regression of BiB genetic effect estimates on publicly available (Chen et. al., 2021[1]) genetic effect estimates, for 66 genome-wide significant fasting glucose variants. The regression intercept was -0.009 (95% CI -0.026, 0.008), the slope was 1.452 (95% CI 0.742, 2.163) and the *P* value for the null hypothesis that the slope was equal to one was 0.212.

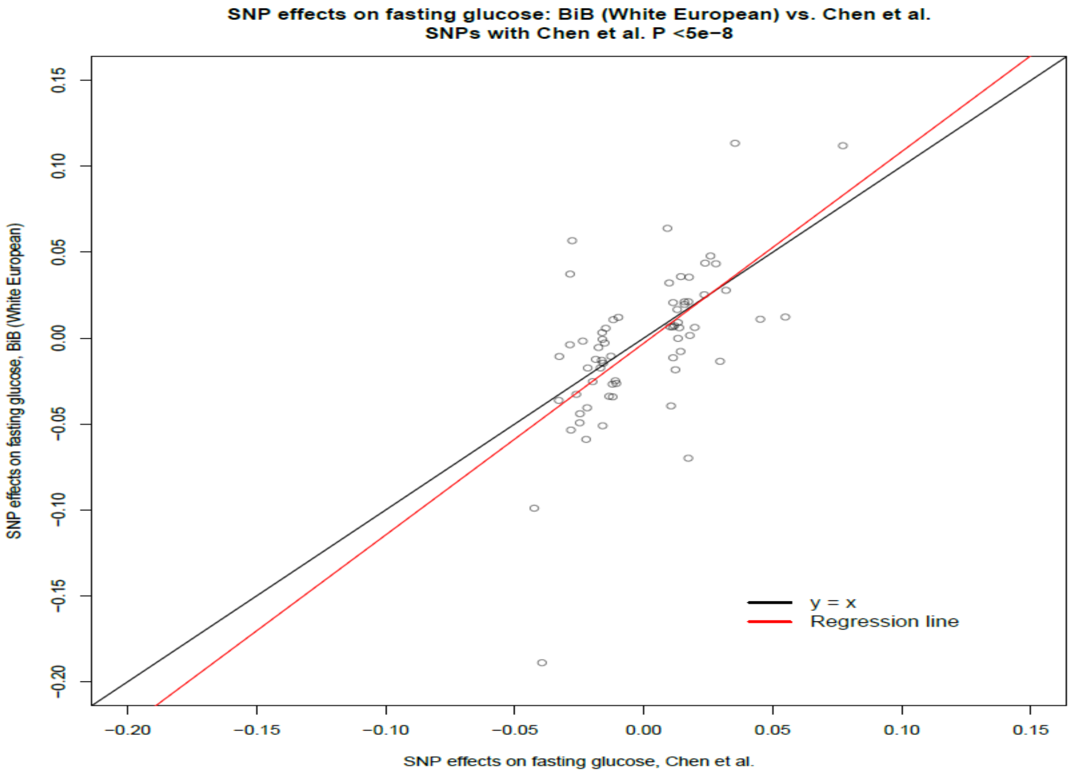

The plot below shows publicly available GWAS (Chen et. al., 2021[1]) genetic effect estimates for fasting glucose, for 66 individual genome-wide significant fasting glucose variants and the overall pooled estimate (0.0206, 95% CI: 0.0179, 0.0234)

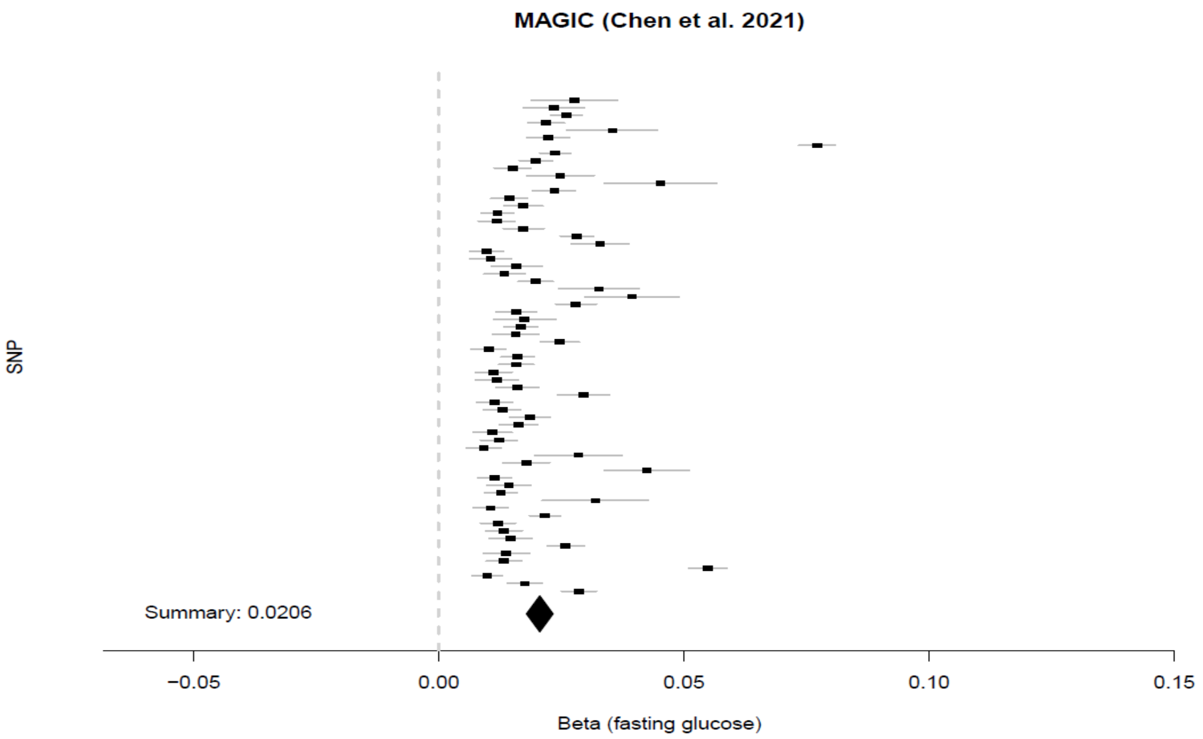

The plot below shows genetic effect estimates for fasting glucose in 3,044 pregnant BiB participants, for 66 individual genome-wide significant fasting glucose variants (Chen et. al., 2021[1]) and the overall pooled estimate (0.0246, 95% CI: 0.0191, 0.0302)

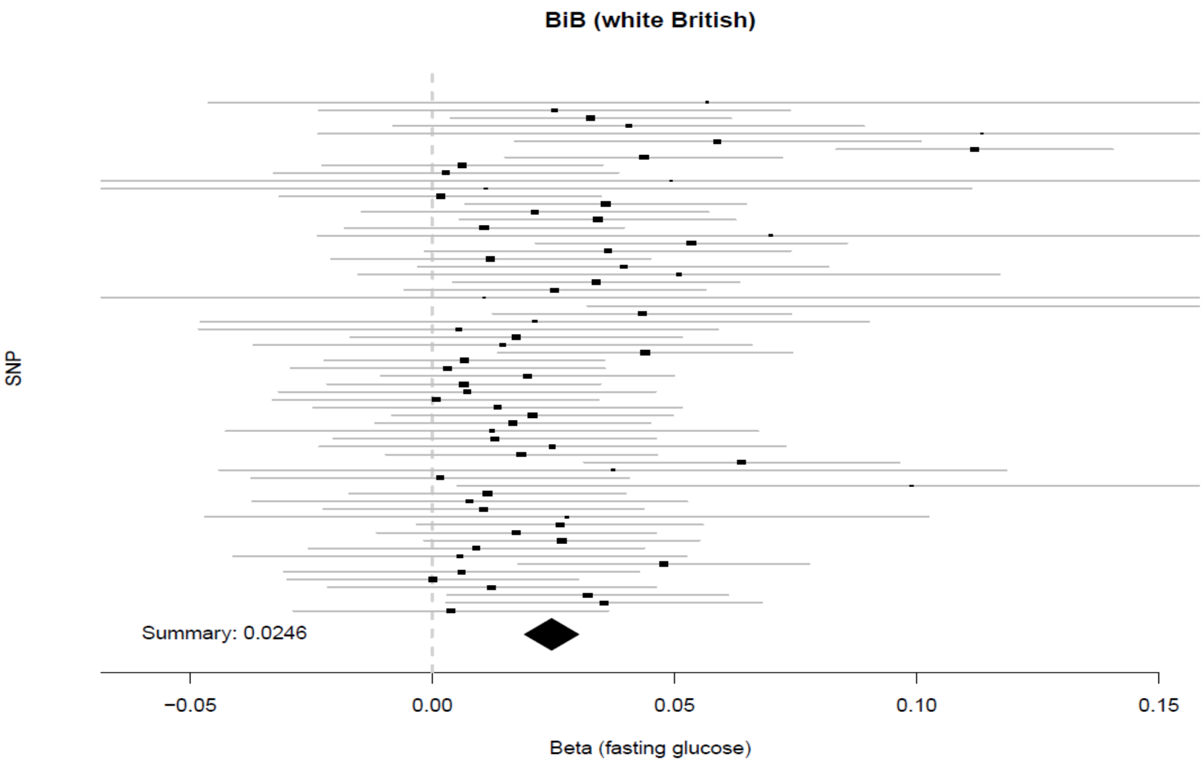

**Supplementary information S3. Study cohorts and genotyping.**

**The Trøndelag Health Study (HUNT).** HUNT is a large population-based study, which invited the entire adult population (≥20 years, ~230,000 participants) of Trøndelag to attend clinical examinations and answer questionnaires[10-13]. So far, four rounds of HUNT have been conducted, HUNT1 (1984 to 1986, n=75,027 participants, 86.8% of invited), HUNT2 (1995 to 1997, n=65,402 participants, 69.7% of invited), HUNT3 (2006 to 2008, n=50,663 participants, 54.0% of invited), and HUNT4 (2017 to 2019, n=56,042 participants, 54.0% of invited).

Genotyping, imputation, and quality control of HUNT genetic data have been described in detail elsewhere[11,13]. About 88% (n=69,716) of participants from HUNT2 and HUNT3 have genotype data. In summary, HUNT2-3 samples were genotyped using one of three different Illumina HumanCoreExome arrays (HumanCoreExome12 v1.0, HumanCoreExome12 v1.1 and UM HUNT Biobank v1.0)[11,13]. Array genotypes were imputed to a customized HRC[6] reference panel including 2,201 Norwegian (HUNT) participants with whole genome sequencing data[11,13] using the Minimac3 v2.0.1[14]. An ancestry informative principal components (PC) analysis using participants from the Human Genome Diversity Project (HGDP) reference panel was previously conducted to define participants of European ancestry[15,16]. Genetic PCs were generated using PLINK[8]. Only individuals of European ancestry were included in the study.

Furthermore, individual-level information from HUNT and the Medical Birth Registry of Norway (MBRN)[17] was linked. MBRN commenced in 1967, therefore all the HUNT participants born after 1967 have a record of their own birthweight and their offspring birthweight (even if offspring are not participants of HUNT). Around 20% (n=14,402) of HUNT genotyped participants have their own birthweight recorded in MBRN. The validity of this information has been reported elsewhere[18].

**UK Biobank (UKB).** UKB is a cohort which invited 9.2 million eligible UK adults aged 40-69 years, between 2006 and 2010, of which n=503,317 participated (5.4% of those invited). Participants attended clinical examinations and answered questionnaires at one of 22 study assessment centers located throughout England, Scotland and Wales[19].

Genotyping, imputation, and quality control of UKB genetic data have been described in detail elsewhere[20]. About 88% (n=440,376) of participants from UKB have genotype data (**Figure 1**). In summary, UKB samples were genotyped using the Applied Biosystems UK BiLEVE Axiom Array and UK Biobank Axiom Array[20]. Array genotypes were imputed to the Haplotype Reference Consortium (HRC)[6] and merged UK10K and 1000 Genomes phase 3[21] reference panels using the IMPUTE4 software package[22]. A PC analysis using participants from Phase 3 of the 1000 Genomes project[23] reference panel was used to identify participants of European ancestry. Genetic PCs were generated using FlashPCA2[24]. Only individuals of European ancestry were included in present study.

**Avon Longitudinal Study of Parents and Children (ALSPAC).** ALSPAC is a birth cohort, which invited pregnant women (expected dates of delivery 1 April 1991 to 31 December 1992, n=14,541 initial enrolled pregnancies, 80% of those invited) resident in Avon, England to attend clinical examinations and answer questionnaires[25]. Further enrolments after 1998 resulted in a baseline sample of 14,901 children alive at one year of age. We used data collected during pregnancy/birth and the at 24-year follow up.

Genotyping, imputation, and quality control of ALSPAC genetic data have been described in detail elsewhere[26]. ALSPAC children, mothers, and fathers were genotyped using the Illumina HumanHap550 quad chip, Illumina human660W-quad array, and Illumina HumanCoreExome array, respectively. Array genotypes were imputed to the Haplotype Reference Consortium (HRC) reference panel[6] using IMPUTE3[22] for ALSPAC children and mothers, and the 1000 Genomes phase 1 reference panel[23] using the Michigan Imputation Server[14] for ALSPAC fathers. Ancestry informative multidimensional scaling analysis using participants from the Hapmap II (for ALSPAC mothers and children) and 1000 Genomes project[23] Phase 3 (for ALSPAC fathers) reference panels was previously conducted to identify participants of European ancestry. Only individuals of European ancestry were included in the present study.

**Early Growth Genetics (EGG) Consortium.** The EGG consortium facilitates the joint analyses of genetic data from multiple studies for birth related phenotypes including birthweight. Warrington et. al.[27] performed GWAS meta-analysis of birthweight using European ancestry data from EGG (including ALSPAC, described above) and UK Biobank (described above). The GWAS was performed in two sub-samples, i) maternal (maternal genotype and offspring birthweight) and ii) offspring (offspring genotype and offspring birthweight). In the unadjusted GWAS, 210,267 maternal samples were included from 12 studies in EGG (including ALSPAC) and UK Biobank, and 298,142 offspring samples were included from 35 studies in EGG (including ALSPAC) and UK Biobank. The study cohorts, genotyping, and methods were described in detail by Warrington et. al.[27]. In short, the multiple births and participants with gestational age <37 weeks were excluded if sufficient data were available. Sex-specific birthweight *z*-score was regressed on genotype using an additive genetic model, adjusted for gestational age (if available), in each sample separately. Fixed-effect meta-analysis was then performed for unadjusted GWAS summary statistics in each sample across studies.

**deCODE genetics.** deCODE genetics[28] is a Icelandic nationwide research program, which included large part of the Icelandic population (n=360,000). deCODE genetics has been linked with the Icelandic birth register, enabling access to birth related outcomes including birthweight for 142,447 Icelanders born between 1982 and 2016. Juliusdottir et. al.[28] performed GWAS of birthweight in European ancestry participants. The GWAS was performed in three sub-samples, i) maternal (maternal genotype and offspring birthweight; 59,736 participants), ii) paternal (paternal genotype and offspring birthweight; 60,134 participants), and offspring (offspring genotype and offspring birthweight; 125,541 participants). The study cohort, genotyping, and methods were described in detail by Juliusdottir et. al.[28]. In short, multiple births, still births, and out-of-term births (gestational age <258 days and gestational age ≥294 days) were excluded. A linear mixed-effect model adjusted for sex, year of birth, and gestational age, implemented in BOLT-LMM[29] was applied in each sample separately. GWAS summary statistics from the maternal and offspring samples were meta-analyzed with the corresponding samples from EGG consortium[27] (described above).

**Supplementary information S4. Identifying genotyped parent–offspring pairs.**

**HUNT.** The procedure used to identify parent-offspring pairs has been described elsewhere[30,31]. In short, a second stage of genotype data cleaning was performed where, firstly, we removed 760 individuals whose inferred sex contradicted their updated reported gender (n=348) and who showed high or low heterozygosity (±5 SD from the mean) (n=412) before kinship analysis, and secondly, removed SNPs with minor allele frequency <0.005 or more than 5% missing rate. Finally, for the kinship analysis to identify the parent-offspring pairs, we used genotyped SNPs shared across the arrays on autosomal chromosomes (257,488 SNPs) and used the recommended thresholds for relatedness implemented as part of KING software v2.2.4[32]. We excluded parent-offspring pairs with 15 years or fewer difference in birth year. The number of offspring was up to 8 per parent and we included up to 6 offspring per parent (**Supplementary information S4**). We included up to 26,028 mother-offspring pairs, 19,773 father-offspring pairs, and 69,635 offspring in the study depending on completeness of phenotype information (**Figure 1**).

The following table presents an overview of total numbers of genotyped-offspring per genotyped-parent available in HUNT.

| **Parents** | **Number of unique parents** | **Total number of offspring** | **Number of offspring per parents** | | | | | | | |
|---|---|---|---|---|---|---|---|---|---|---|
| | | | 1 | 2 | 3 | 4 | 5 | 6 | 7 | 8 |
| Mothers | 15261 | 26057 | 15261 | 7165 | 2542 | 794 | 213 | 65 | 15 | 2 |
| Fathers | 11867 | 19793 | 11867 | 5382 | 1838 | 521 | 141 | 36 | 7 | 1 |

Additionally, parent-offspring pairs with birthweight information were retrieved from the MBRN[17] (described above). In MBRN, we identified 58,648 mother (genotyped)-offspring (phenotyped) pairs, 52,285 father (genotyped)-offspring (phenotyped) pairs, and 14,338 HUNT offspring with both genotype and phenotype information.

**UKB.** A similar procedure as in HUNT was used to identify parent-offspring pairs via KING[32], which has been described elsewhere[31]. We excluded parent-offspring pairs with 15 years or fewer difference in birth year and included only one randomly selected offspring from each sibling group. We removed cryptic relatedness (kinship coefficient >0.0442 by KING[32]) amongst mothers and fathers but not offspring, because the offspring sample was large enough to fit the fastGWA linear mixed model which accounts for relatedness. In total we included 3,763 unrelated mother-offspring pairs, 1,700 unrelated father-offspring pairs, and 439,914 offspring in the study.

**ALSPAC.** Parent-offspring pairs were identified by linking mother, father, and offspring study ID, including a QC check for familial relatedness across the overall father-offspring sample via the genetic data using procedures similar to HUNT. We included only one randomly selected offspring from the sibling groups. We removed cryptic relatedness amongst mothers [identity by descent (IBD) >0.125 calculated by PLINK (v1.07)[33]], fathers (using a relatedness filter of 0.05 with the --grm-singleton command in the GCTA software package[9]), and offspring (IBD >0.1, calculated by PLINK (v1.07)[33]. In total we included 7,672 unrelated mother-offspring pairs, 1,634 unrelated father-offspring pairs, and 7,363 unrelated offspring in the study.

**Supplementary information S5. Birthweight and cardiometabolic risk factors**

**HUNT.** Birthweight in kg and gestational age in days were retrieved from MBRN[17]. Multiple births (n=214) were excluded from the analyses. Additionally, for analyses of birthweight we excluded individuals if they were born before 258 or after 301 days of gestation (n=1,805). Sex-specific birthweight z-score was generated for analyses. To exclude outliers, we removed values of birthweight z-score more than 5.0 standard deviations from the mean.

Clinical measurements of cardiometabolic risk factors were used from HUNT2-4[10-13] where the most recent measurement was prioritized. Body mass index (BMI, kg/m$^2$) was calculated as weight (kg) divided by the squared value of height (meters) in HUNT2-4. Weight and height were measured in light clothes and without shoes. WHR (cm/cm) was calculated as waist circumference (cm) divided by hip circumference (cm) in HUNT2-4. Waist and hip circumference were measured in light clothing and standing with arms hanging relaxed. Waist circumference was measured horizontally at the height of the umbilicus. Hip circumference was measured horizontally at the thickest part of the hip. SBP (mmHg) and DBP (mmHg) were measured three times (each 1 minute apart) using automated measures based on oscillometry in HUNT2-4[34]. The average of the second and third measurement was used for calculation of SBP and DBP but for participants who had only two measurements, the second measurement was used. Glucose (mmol/L) was measured by an enzymatic hexokinase method using Hitachi 911 Autoanalyzer in HUNT2 and by a hexokinase/G-6-PDH method using Architect ci8200 in HUNT3. HbA1c (mmol/mol) was measured by enzymatic HbA1c assay using Architect ci8200 instrument in HUNT4. TG (mmol/L), TC (mmol/L), and HDL-C (mmol/L) were measured using Hitachi 911 Autoanalyzer in HUNT2 and Architect ci8200 in HUNT3-4. TG was measured by an enzymatic colorimetric method in HUNT2 and a glycerol phosphate oxidase method in HUNT3-4. TC was measured by an enzymatic cholesterol esterase method in HUNT2-4. HDL-C was measured by an enzymatic cholesterol oxidase method in HUNT2 and an accelerator selective detergent method in HUNT3-4. LDL-C (mmol/L) cholesterol was calculated using the Friedewald formula[35] where participants with triglycerides >4.5mmol/L were excluded. CRP (mg/L) was measured by a latex immunoassay method using Architect ci8200 instrument in HUNT3-4. The detection limit of CRP was 0.1 to 160 where the measurements below and above this range were recorded as 0.09 and 160.01, respectively.

All the above clinical measurements of cardiometabolic risk factors were taken at a non-fasting state. To exclude outliers, we removed values of cardiometabolic traits more than 4.56 standard deviations from the mean. CRP was strongly right skewed; therefore, the values were natural log transformed. In the sensitivity analyses, we log transformed BMI, WHR, glucose, HbA1c, TG, and HDL-C.

**UKB.** Birthweight in kg was reported retrospectively at the recruitment questionnaire during participation in UKB. We excluded participants who were part of a multiple birth (1.3% of participants) from analyses of birthweight. No individuals had >1kg difference between birthweight reported at different visits. Additionally, participants with birthweight <2.5kg or >4.5kg were excluded (7.2% of participants). Sex-specific z-scores of birthweights were generated for analyses.

BMI (kg/m$^2$) was calculated as weight (kg) divided by the squared value of height (meter). Weight and height were measured using the Tanita BC-418MA body composition analyser (without shoes and heavy clothing), and the Saca 202 device (barefoot in a standing position), respectively. WHR (cm/cm) was calculated as waist circumference (cm) divided by hip circumference (cm). Waist and hip circumference were measured using the Wessex non-stretchable sprung tape measure. SBP (mmHg) and DBP (mmHg) were measured up to twice using either an automated machine (Omron 705 IT electronic blood pressure monitor) or manually using a sphygmomanometer with standard procedures. The average of the two measurements was used for calculation of SBP and DBP if two measurements were available (>97% of participants). Glucose (mmol/L) was measured by the hexokinase method using Beckman Coulter AU5800 Clinical Chemistry Analyzer with Beckman Coulter reagents. HbA1c (mmol/mol) was measured using a high-performance liquid chromatography (HPLC) method by the VARIANT II TURBO Hemoglobin Testing System (Bio-Rad reagents). TG (mmol/L), TC (mmol/L), HDL-C (mmol/L), and LDL-C (mmol/L) cholesterol were measured using Beckman Coulter AU5800 Clinical Chemistry Analyzer, Beckman Coulter reagents. TG and TC were measured by an enzymatic (CHOD-POD) method. HDL-C cholesterol was measured by an enzyme immunoinhibition method. LDL-C cholesterol was measured by an enzymatic selective protection method. CRP (mg/L) was measured by an immunoturbidimetric method using Beckman Coulter AU5800 Clinical Chemistry Analyzer, Beckman Coulter reagents.

All the above clinical measurements of cardiometabolic risk factors were taken at a non-fasting state. To exclude outliers, we removed values of cardiometabolic traits more than 4.56 standard deviations from the mean. CRP was strongly right skewed; therefore, the values were natural log transformed. In the sensitivity analyses, we log transformed BMI, WHR, glucose, HbA1c, TG, and HDL-C.

**ALSPAC.** Birthweight in in kg and gestational age in weeks were measured by trained research assistants or retrieved from the birth record or from the birth notification. We excluded birthweight for participants who were part of a multiple births (3.3% of participants), Additionally, we excluded individuals if they were born before 37 completed weeks of gestation or after 42 weeks plus 6 days of gestation (8.4% of participants). Sex-specific birthweight z-score was generated for analysis. To exclude outliers, we removed values of birthweight z-score more or less than 5.0 standard deviations from the mean (1.3% of participants).

BMI (kg/m$^2$) was calculated as weight (kg) divided by the squared value of height (meter). Weight and height were measured using Tanita TBF-401A electronic body composition scales and Harpenden wall-mounted stadiometer, respectively. WHR (cm/cm) was calculated as waist circumference (cm) divided by hip circumference (cm). Waist and hip circumference were measured using Seca 201 body tension tape and were repeated twice for accuracy. SBP (mmHg) and DBP (mmHg) were measured after two minutes rest by Omron M6 upper arm blood pressure/pulse monitor. The average of up to three seated measurement was used for calculation of SBP and DBP if more than 2 measurements were available. Glucose (mmol/L) was measured by the hexokinase method using GLUC3 (Glucose HK) Cat. No. 04404483 190 kit (Roche Diagnostics GmbH, Sandhofer Strasse 116, D-68305 Mannheim, Germany). No information on HbA1c is available in ALSPAC. TG (mmol/L) was measured by enzymatic colorimetric test using HDLC3 (HDL-Cholesterol plus 3rd generation) Cat. No. 04399803 190 kit (Roche Diagnostics GmbH, Sandhofer Strasse 116, D-68305 Mannheim, Germany). TC (mmol/L) and HDL-C (mmol/L) were measured by enzymatic colorimetric test using CHOL2 (Cholesterol gen.2) Cat. No. 03039773 190 kit (Roche Diagnostics GmbH, Sandhofer Strasse 116, D-68305 Mannheim, Germany). LDL-C (mmol/L) cholesterol was calculated using the Friedewald formula[35] where participants with triglycerides >4.5mmol/L were excluded. CRP (mg/L) was measured by particle enhanced immunoturbidimetric assay using CRPHS (Cardiac C-Reactive Protein (Latex) High Sensitive) Cat. No. 04628918 190 kit (Roche Diagnostics GmbH, Sandhofer Strasse 116, D-68305 Mannheim, Germany).

All the above clinical measurements of cardiometabolic risk factors were taken at a fasting state (90% of participants fasted for at least 8 hours prior to blood sampling). To exclude outliers, we removed values of cardiometabolic traits more or less than 4.56 standard deviations from the mean. CRP was strongly right skewed; therefore, the values were natural log transformed. In the sensitivity analyses, we log transformed BMI, WHR, glucose, TG, and HDL-C.

**Correction for medication use.** In HUNT and UKB, we added 15 mmHg to SBP and 10 mmHg to DBP values of participants who reported taking blood pressure lowering medication, which has been shown to reduce bias in the estimation of effects on blood pressure relative to including blood pressure lowering medication as a covariate[36]. In UKB, we divided LDL cholesterol values by 0.7 and total cholesterol and triglyceride values by 0.8 for participants who reported taking lipid lowering medication. In HUNT2, very few participants reported lipid lowering medication use at the time of lipid measurement, and data on lipid medication use was not available for HUNT3 at the time of analysis. In ALSPAC, very few participants reported lipid lowering or antihypertensive medication use at age 24 years.

**Covariates.** Administrative information on age, sex, genotyping batch, survey (for HUNT, describes which HUNT survey the participant attended), assessment center (for UKB, describes which assessment center participants attended), and fasting time (for UKB, describes time since last meal) were recorded by HUNT, UKB, and ALSPAC. In HUNT, age (in years) at participation was retrieved for the corresponding HUNT survey where specific cardiometabolic measurements were taken. Principal components (PCs) were included as covariates (20 in HUNT, 40 in UKB, and 20 in ALSPAC) (covariates are detailed in **Supplementary information S7**).

**Supplementary information S6. Sample size of offspring outcomes in unadjusted GWAS.**

| Offspring outcome (units) | Maternal sample | | | | Paternal sample | | | | Offspring sample | | | |
|---|---|---|---|---|---|---|---|---|---|---|---|---|
| | HUNT | UKB | ALSPAC | Meta-analysis | HUNT | UKB | ALSPAC | Meta-analysis | HUNT | UKB | ALSPAC | Meta-analysis |
| Birthweight (kg)[a] | 49,845 | NA[a] | NA[a] | 319,847 | 44,629 | 1,238 | 1,545 | 107,546[b] | 12,330 | NA[a] | NA[a] | 436,013 |
| BMI (kg/m$^2$) | 25,998 | 3,742 | 2,587 | 32,327 | 19,757 | 1,692 | 1,043 | 22,492 | 68,316 | 438,258 | 2,767 | 509,341 |
| WHR (cm/cm) | 25,953 | 3,756 | 2,583 | 32,292 | 19,709 | 1,698 | 1,041 | 22,448 | 68,294 | 439,481 | 2,763 | 510,538 |
| SBP (mmHg) | 26,013 | 3,758 | 2,601 | 32,372 | 19,761 | 1,700 | 1,049 | 22,510 | 68,583 | 439,871 | 2,785 | 511,239 |
| DBP (mmHg) | 26,011 | 3,758 | 2,602 | 32,371 | 19,760 | 1,700 | 1,048 | 22,508 | 68,555 | 439,914 | 2,784 | 511,253 |
| Glucose (mmol/L) | 25,841 | 3,233 | 2,126 | 31,200 | 19,614 | 1,481 | 892 | 21,987 | 68,043 | 380,633 | 2,320 | 450,996 |
| HbA1c (mmol/mol)[c] | 16,813 | 3,587 | NA[c] | 20,400 | 12,855 | 1,624 | NA[c] | 14,479 | 35,168 | 416,643 | NA[c] | 451,811 |
| TC (mmol/L) | 25,960 | 3,583 | 2,135 | 31,678 | 19,715 | 1,625 | 894 | 22,234 | 68,501 | 419,577 | 2,326 | 490,404 |
| HDL-C (mmol/L) | 25,958 | 3,257 | 2,135 | 31,350 | 19,714 | 1,482 | 894 | 22,090 | 68,502 | 384,017 | 2,326 | 454,845 |
| LDL-C (mmol/L) | 25,753 | 3,576 | 2,134 | 31,463 | 19,572 | 1,625 | 892 | 22,089 | 67,635 | 418,612 | 2,324 | 488,571 |
| TG (mmol/L) | 26,028 | 3,568 | 2,121 | 31,717 | 19,773 | 1,619 | 887 | 22,279 | 68,627 | 417,559 | 2,312 | 488,498 |
| CRP (mg/L)[d] | 22,375 | 3,584 | 1,981 | 27,940 | 17,110 | 1,622 | 833 | 19,565 | 51,581 | 418,969 | 2,161 | 472,711 |

[a]-publicly available maternal and offspring birthweight GWAS data from EGG (early growth genetics) already included UKB and ALSPAC participants, therefore analyses were not rerun for these samples, [b]- publicly available paternal and offspring birthweight GWAS data from deCODE (deCODE genetics) was meta-analyzed together with HUNT, UKB, and ALSPAC participants, [c]-not available in ALSPAC, [d]-natural log transformed for the main analyses.

**Supplementary information S7. Covariates of GWAS models of offspring outcomes in unadjusted GWAS.**

| Cohort | Sample | Covariates for GWAS of offspring cardiometabolic risk factors | Covariates for GWAS of offspring birthweight |
|---|---|---|---|
| **HUNT** | Maternal genotype | Offspring age, offspring age$^2$, offspring sex, offspring age × offspring sex, offspring age$^2$ × offspring sex, offspring HUNT study wave, 20 maternal genetic principal components, maternal genotyping batch | Offspring gestational age, offspring HUNT study wave, 20 maternal genetic principal components, maternal genotyping batch |
| | Paternal genotype | Offspring age, offspring age$^2$, offspring sex, offspring age × offspring sex, offspring age$^2$ × offspring sex, offspring HUNT study wave, 20 paternal genetic principal components, paternal genotyping batch | Offspring gestational age, offspring HUNT study wave, 20 paternal genetic principal components, paternal genotyping batch |
| | Offspring genotype | Offspring age, offspring age$^2$, offspring sex, offspring age × offspring sex, offspring age$^2$ × offspring sex, offspring HUNT study wave, 20 offspring genetic principal components, offspring genotyping batch | Offspring gestational age, offspring HUNT study wave, 20 offspring genetic principal components, offspring genotyping batch |
| **UKB** | Maternal genotype | Offspring age, offspring age$^2$, offspring sex, offspring age × offspring sex, offspring age$^2$ × offspring sex, offspring fasting time[a], offspring assessment center, 20 maternal genetic principal components, maternal genotyping array | Already included in publicly available **deCODE and EGG** meta-analysis, therefore not run |
| | Paternal genotype | Offspring age, offspring age$^2$, offspring sex, offspring age × offspring sex, offspring age$^2$ × offspring sex, offspring fasting time[a], offspring assessment center, 20 paternal genetic principal components, paternal genotyping array | 20 paternal genetic principal components, paternal genotyping array, paternal assessment center |
| | Offspring genotype | Offspring age, offspring age$^2$, offspring sex, offspring age × offspring sex, offspring age$^2$ × offspring sex, offspring fasting time[a], offspring assessment center, 20 offspring genetic principal components, offspring genotyping array | Already included in publicly available **deCODE and EGG** meta-analysis, therefore not run |
| **ALSPAC** | Maternal genotype | Offspring age, offspring age$^2$, offspring sex, offspring age × offspring sex, offspring age$^2$ × offspring sex, 20 maternal genetic principal components | Already included in publicly available **deCODE and EGG** meta-analysis, therefore not run |
| | Paternal genotype | Offspring age, offspring age$^2$, offspring sex, offspring age × offspring sex, offspring age$^2$ × offspring sex, 20 paternal genetic principal components | Offspring gestational age, 20 paternal genetic principal components |

| | | | |
|---|---|---|---|
| | Offspring genotype | Offspring age, offspring age$^2$, offspring sex, offspring age × offspring sex, offspring age$^2$ × offspring sex, 20 offspring genetic principal components | Already included in publicly available **deCODE and EGG** meta-analysis, therefore not run |
| **deCODE** | Paternal genotype | - | Publicly available deCODE GWAS |
| | Offspring genotype | - | Publicly available deCODE GWAS |

[a]-Fasting time before the blood sample was drawn was included as a covariate in UKB analyses of glucose, total cholesterol, HDL cholesterol, LDL cholesterol and triglycerides. For all cardiometabolic risk factors as outcomes, offspring age at outcome measurement was included as a covariate.

**Supplementary information S8. Quality control indices for unadjusted GWAS.**

| Offspring outcome (units) | Maternal sample | | | Paternal sample | | | Offspring sample | | |
|---|---|---|---|---|---|---|---|---|---|
| | **Lambda GC** | **LDSC intercept** | **LDSC attenuation ratio** | **Lambda GC** | **LDSC intercept** | **LDSC attenuation ratio** | **Lambda GC** | **LDSC intercept** | **LDSC attenuation ratio** |
| Birthweight (kg) | 1.41 | 1.20 | 0.21 | 1.11 | 1.11 | 0.52 | 1.44 | 1.28 | 0.23 |
| BMI (kg/m$^2$) | 1.03 | 1.01 | 0.24 | 1.02 | 1.03 | 1.02 | 2.05 | 1.11 | 0.16 |
| WHR (cm/cm) | 1.03 | 1.02 | 0.37 | 1.02 | 1.02 | 0.89 | 1.76 | 1.10 | 0.06 |
| SBP (mmHg) | 1.02 | 1.01 | 0.27 | 1.01 | 1.00 | 0.03 | 1.77 | 1.16 | 0.09 |
| DBP (mmHg) | 1.04 | 1.02 | 0.40 | 1.03 | 1.02 | 0.53 | 1.78 | 1.16 | 0.09 |
| Glucose (mmol/L) | 1.01 | 1.00 | 0.16 | 1.00 | 1.01 | – | 1.25 | 1.06 | 0.15 |
| HbA1c (mmol/mol) | 1.02 | 1.03 | 1.14 | 1.01 | 1.01 | 0.71 | 1.53 | 1.19 | 0.15 |
| TC (mmol/L) | 1.03 | 1.02 | 0.75 | 1.02 | 1.01 | 0.39 | 1.42 | 1.18 | 0.17 |
| HDL-C (mmol/L) | 1.01 | 1.02 | 0.56 | 1.00 | 1.02 | 1.22 | 1.62 | 1.22 | 0.15 |
| LDL-C (mmol/L) | 1.03 | 1.01 | 0.44 | 1.01 | 1.01 | 0.54 | 1.42 | 1.18 | 0.18 |
| TG (mmol/L) | 1.02 | 1.01 | 0.17 | 1.01 | 1.01 | 1.05 | 1.56 | 1.19 | 0.13 |
| CRP (mg/L) | 1.02 | 1.01 | 0.53 | 1.02 | 1.02 | 0.69 | 1.58 | 1.13 | 0.11 |

BMI (body mass index), GC (genomic control), WHR (waist-hip ratio), SBP (systolic blood pressure), DBP (diastolic blood pressure), HbA1c (glycated hemoglobin), TC (total cholesterol), HDL-C (high density lipoprotein cholesterol), LDL-C (low density lipoprotein cholesterol), LDSC (linkage disequilibrium score regression), TG (triglycerides), CRP (C-reactive protein). CRP: natural log transformed.

**Supplementary information S9. Adjusted GWAS.**

**Trios WLM:**
The unadjusted GWAS from individual cohorts were quality controlled using the GWASinspector R package[37] and the LD score regression software package[38] and then meta-analysis[39] was performed across cohorts in each sample (i.e. maternal, paternal, and offspring samples). Thereafter, a weighted linear model (WLM)[27,40,41] implemented in DONUTS R package[42] was applied to estimate adjusted maternal ($\hat{\beta}_{mat}$), paternal ($\hat{\beta}_{pat}$) and offspring ($\hat{\beta}_{off}$) GWAS summary statistics (i.e. the mutually adjusted coefficients for maternal, paternal and offspring genotype, fitted jointly in the same model) as linear combinations of the unadjusted offspring ($\hat{b}_{off}$), maternal ($\hat{b}_{mat}$) and paternal ($\hat{b}_{pat}$) SNP-outcome estimates:

$$\hat{\beta}_{off} = 2\hat{b}_{off} - \hat{b}_{mat} - \hat{b}_{pat}$$
$$\hat{\beta}_{mat} = \frac{3}{2}\hat{b}_{mat} - \hat{b}_{off} + \frac{1}{2}\hat{b}_{pat}$$
$$\hat{\beta}_{pat} = \frac{3}{2}\hat{b}_{pat} - \hat{b}_{off} + \frac{1}{2}\hat{b}_{mat}$$

and their standard errors:

$$SE(\hat{\beta}_{off}) = \sqrt{4var(\hat{b}_{off}) + var(\hat{b}_{mat}) + var(\hat{b}_{pat}) + 2 \times int_{mat,pat} \times SE(\hat{b}_{mat})SE(\hat{b}_{pat}) - 4 \times int_{off,mat} \times SE(\hat{b}_{off})SE(\hat{b}_{mat}) - 4 \times int_{off,pat} \times SE(\hat{b}_{off})SE(\hat{b}_{pat})}$$

$$SE(\hat{\beta}_{mat}) = \sqrt{\frac{9}{4}var(\hat{b}_{mat}) + var(\hat{b}_{off}) + \frac{1}{4}var(\hat{b}_{pat}) - int_{off,pat} \times SE(\hat{b}_{off})SE(\hat{b}_{pat}) - 3 \times int_{off,mat} \times SE(\hat{b}_{off})SE(\hat{b}_{mat}) + \frac{3}{2} \times int_{mat,pat} \times SE(\hat{b}_{mat})SE(\hat{b}_{pat})}$$

$$SE(\hat{\beta}_{pat}) = \sqrt{\frac{9}{4}var(\hat{b}_{pat}) + var(\hat{b}_{off}) + \frac{1}{4}var(\hat{b}_{mat}) - int_{off,mat} \times SE(\hat{b}_{off})SE(\hat{b}_{mat}) - 3 \times int_{off,pat} \times SE(\hat{b}_{off})SE(\hat{b}_{pat}) + \frac{3}{2} \times int_{mat,pat} \times SE(\hat{b}_{mat})SE(\hat{b}_{pat})}$$

Where, $int_{off,mat}$, $int_{off,pat}$, and $int_{mat,pat}$ are the intercepts calculated via bivariate LD score regression[43] for the unadjusted GWAS summary statistics of maternal and offspring, paternal and offspring, and maternal and paternal samples, respectively. These intercept terms are necessary to account for sample overlap between the unadjusted GWAS summary statistics of maternal, paternal, and offspring sample. We estimated these intercept terms using Hapmap 3 SNPs with MAF ≥1%, excluding SNPs in the MHC region and with association Chi square statistics >80.

*P* values are calculated from trios WLM estimates using a z test, with test statistic:

$$Z(\hat{\beta}_{off}) = \frac{\hat{\beta}_{off}}{SE(\hat{\beta}_{off})}$$

$$Z(\hat{\beta}_{mat}) = \frac{\hat{\beta}_{mat}}{SE(\hat{\beta}_{mat})}$$
$$Z(\hat{\beta}_{pat}) = \frac{\hat{\beta}_{pat}}{SE(\hat{\beta}_{pat})}$$

**Duos WLM:**
Additionally, if we can assume that the adjusted SNP-outcome estimates for fathers are zero ($\hat{\beta}_{pat}$=0) then we can employ a "duos WLM", to estimate adjusted maternal and offspring genetic effects; this model adjusts the maternal adjusted effects for the offspring's genotype only (i.e. not for the father's genotype as well). We will gain smaller standard errors [$SE(\hat{\beta}_{mat})$], and thereafter increased power for intergenerational MR analyses of maternal exposures on the offspring outcomes by fitting a duos WLM (details on MR estimates comparison between duos WLM and trios WLM in **Supplementary information S11**):

$\hat{\beta}_{off} = \frac{4}{3}\hat{b}_{off} - \frac{2}{3}\hat{b}_{mat}$ (for maternal/paternal and offspring duos WLM)
$\hat{\beta}_{mat} = \frac{4}{3}\hat{b}_{mat} - \frac{2}{3}\hat{b}_{off}$ (for maternal and offspring duos WLM)

and their standard errors:
$$SE(\hat{\beta}_{off}) = \sqrt{\frac{16}{9}var(\hat{b}_{off}) + \frac{4}{9}var(\hat{b}_{mat}) - \frac{16}{9} \times int_{off,mat} \times SE(\hat{b}_{mat})SE(\hat{b}_{off})}$$
$$SE(\hat{\beta}_{mat}) = \sqrt{\frac{16}{9}var(\hat{b}_{mat}) + \frac{4}{9}var(\hat{b}_{off}) - \frac{16}{9} \times int_{off,mat} \times SE(\hat{b}_{mat})SE(\hat{b}_{off})}$$

*P* values are calculated from duos WLM estimates using a z test, with test statistic:
$$Z(\hat{\beta}_{off}) = \frac{\hat{\beta}_{off}}{SE(\hat{\beta}_{off})}$$
$$Z(\hat{\beta}_{mat}) = \frac{\hat{\beta}_{mat}}{SE(\hat{\beta}_{mat})}$$

A duos WLM can also be estimated for paternal effects by substituting paternal unadjusted effects and LDSC intercepts into the equations above.

In these WLM analyses, we assumed no assortative mating (mating was random) given. Results from our sensitivity analyses in the Norwegian Mother, Father and Child Cohort Study (MoBa)[44,45] (Bond et. al., 2024) suggested that this gave the most accurate estimates for adjusted estimates on birthweight.

**Supplementary information S10. Re-scaling the MR estimates to standard deviation.**

The unadjusted and adjusted intergenerational MR estimates were re-scaled to the standard deviation (SD) scale, to facilitate comparison of estimates between outcomes.

We converted the betas and standard errors from MR analyses to the SD scale by dividing the betas and standard errors by the weighted mean SD (listed in the table below). The weighted mean SD of each phenotype (offspring outcome) is a weighted average of the corresponding phenotypic SD over the 9 sub-samples in HUNT, UKB, and ALSPAC cohorts (three maternal samples, three paternal samples, and three offspring samples).

The following equation was used to calculate the weighted mean SD [sqrt ($S_w^2$)] for each phenotype.

$$s_w^2 = \frac{(n_1-1)\, s_1^2 + (n_2-1)\, s_2^2 + ¼ + (n_k-1)\, s_k^2}{n_1 + n_2 + ¼ + n_k - k}$$

Where, we used the following parameters:

$n$ = sample size of sub-sample, where $n_1$ represents sample size of sample 1 (for example, maternal sample size in HUNT).

$s^2$ = variance of a phenotype in the sub-sample, where $s_1^2$ represents variance of a phenotype in sample 1 (for example, Birthweight variance in maternal sample in HUNT).

$k$ = total number of sub-samples, which is 9.

| **Offspring outcomes** | **Weighted mean standard deviation (SD)** | **Offspring outcomes** | **Weighted mean standard deviation (SD)** |
|---|---|---|---|
| BMI (kg/m$^2$) | 4.650540 | TC (mmol/L) | 1.102393 |
| log BMI (kg/m$^2$) | 0.165590 | HDL-C (mmol/L) | 0.377207 |
| WHR (cm/cm) | 0.090530 | log HDL-C (mmol/L) | 0.260264 |
| log WHR (cm/cm) | 0.103256 | LDL-C (mmol/L) | 0.889462 |
| SBP (mmHg) | 20.791015 | TG (mmol/L) | 1.026870 |
| DBP (mmHg) | 11.476896 | log TG (mmol/L) | 0.530413 |
| Glucose (mmol/L) | 0.904931 | CRP (mg/L) | 3.456568 |
| log glucose (mmol/L) | 0.157869 | log CRP (mg/L) | 1.056069 |
| HbA1c (mmol/mol) | 5.111408 | | |

| | |
|---|---|
| log HbA1c (mmol/mol) | 0.136360 |

BMI (body mass index), WHR (waist-hip ratio), SBP (systolic blood pressure), DBP (diastolic blood pressure), HbA1c (glycated hemoglobin), TC (total cholesterol), HDL-C (high density lipoprotein cholesterol), LDL-C (low density lipoprotein cholesterol), TG (triglycerides), CRP (C-reactive protein), log (natural log transformed).

### Supplementary information S11. Comparison of adjusted intergenerational MR estimates from adjusted GWAS in duos WLM and in trios WLM.

| Offspring outcome | Sub−sample | Model | Fasting Glucose beta (95% CI) | p value | Type−2 Diabetes beta (95% CI) | p value | Gestational Diabetes beta (95% CI) | p value |
|---|---|---|---|---|---|---|---|---|
| Birthweight (kg) | Maternal | Duos | 0.418 (0.304, 0.533) | 9.1e−13 | 0.081 (0.061, 0.101) | 9.7e−16 | 0.149 (0.095, 0.203) | 6.2e−08 |
| | | Trios | 0.349 (0.228, 0.469) | 1.3e−08 | 0.063 (0.041, 0.086) | 3.9e−08 | 0.133 (0.084, 0.183) | 1.4e−07 |
| | Offspring | Duos | −0.368 (−0.514, −0.223) | 6.9e−07 | −0.072 (−0.097, −0.047) | 1.5e−08 | −0.109 (−0.247, 0.029) | 0.121 |
| | | Trios | −0.231 (−0.391, −0.071) | 0.005 | −0.04 (−0.069, −0.011) | 0.008 | −0.08 (−0.178, 0.019) | 0.112 |
| BMI (kg/m$^2$) | Maternal | Duos | 0.088 (−0.09, 0.267) | 0.331 | −0.007 (−0.043, 0.03) | 0.726 | −0.016 (−0.105, 0.072) | 0.716 |
| | | Trios | 0.128 (−0.07, 0.326) | 0.206 | −0.008 (−0.051, 0.035) | 0.727 | −0.023 (−0.131, 0.084) | 0.668 |
| | Offspring | Duos | −0.131 (−0.257, −0.005) | 0.041 | 0.059 (0.023, 0.095) | 0.001 | −0.032 (−0.093, 0.028) | 0.297 |
| | | Trios | −0.21 (−0.416, −0.003) | 0.046 | 0.061 (0.011, 0.111) | 0.017 | −0.018 (−0.139, 0.103) | 0.77 |
| WHR (cm/cm) | Maternal | Duos | −0.022 (−0.147, 0.104) | 0.736 | −0.004 (−0.032, 0.024) | 0.776 | −0.07 (−0.131, −0.01) | 0.023 |
| | | Trios | −0.054 (−0.209, 0.101) | 0.495 | −0.009 (−0.044, 0.025) | 0.599 | −0.113 (−0.187, −0.039) | 0.003 |
| | Offspring | Duos | 0.018 (−0.068, 0.104) | 0.674 | 0.076 (0.055, 0.098) | 5.2e−12 | 0.035 (−0.01, 0.079) | 0.131 |
| | | Trios | 0.081 (−0.078, 0.239) | 0.318 | 0.084 (0.047, 0.121) | 7.4e−06 | 0.122 (0.044, 0.2) | 0.002 |
| SBP (mmHg) | Maternal | Duos | −0.016 (−0.13, 0.098) | 0.78 | −0.009 (−0.035, 0.016) | 0.485 | −0.017 (−0.072, 0.037) | 0.535 |
| | | Trios | 0.015 (−0.126, 0.156) | 0.838 | −0.01 (−0.041, 0.021) | 0.539 | −0.022 (−0.088, 0.044) | 0.515 |
| | Offspring | Duos | 0.055 (−0.056, 0.166) | 0.334 | 0.061 (0.04, 0.082) | 8.4e−09 | 0.02 (−0.021, 0.062) | 0.342 |
| | | Trios | −0.007 (−0.192, 0.178) | 0.941 | 0.062 (0.028, 0.097) | 3.7e−04 | 0.03 (−0.036, 0.096) | 0.372 |
| DBP (mmHg) | Maternal | Duos | 0.054 (−0.082, 0.19) | 0.434 | −0.005 (−0.035, 0.025) | 0.749 | 0.01 (−0.056, 0.075) | 0.769 |
| | | Trios | 0.152 (−0.022, 0.326) | 0.086 | −0.003 (−0.039, 0.034) | 0.892 | 0.025 (−0.055, 0.105) | 0.546 |
| | Offspring | Duos | −0.126 (−0.257, 0.005) | 0.06 | 0.042 (0.019, 0.066) | 3.4e−04 | −0.028 (−0.064, 0.007) | 0.119 |
| | | Trios | −0.324 (−0.533, −0.114) | 0.002 | 0.037 (−0.001, 0.076) | 0.056 | −0.057 (−0.156, 0.041) | 0.253 |
| Glucose (mmol/L) | Maternal | Duos | −0.23 (−0.404, −0.056) | 0.01 | −0.018 (−0.051, 0.016) | 0.296 | 0.008 (−0.071, 0.088) | 0.837 |
| | | Trios | −0.216 (−0.435, 0.003) | 0.054 | −0.028 (−0.069, 0.013) | 0.174 | 0.034 (−0.063, 0.132) | 0.489 |
| | Offspring | Duos | 1.121 (0.986, 1.255) | 3.2e−60 | 0.199 (0.172, 0.226) | 5.6e−48 | 0.249 (0.094, 0.404) | 0.002 |
| | | Trios | 1.087 (0.842, 1.332) | 3.6e−18 | 0.221 (0.174, 0.267) | 7.2e−21 | 0.195 (0.002, 0.388) | 0.047 |
| HbA1C (mmol/mol) | Maternal | Duos | 0 (−0.181, 0.181) | 0.999 | 0.002 (−0.04, 0.044) | 0.918 | 0.041 (−0.058, 0.141) | 0.413 |
| | | Trios | −0.023 (−0.242, 0.196) | 0.839 | −0.014 (−0.064, 0.037) | 0.597 | 0.05 (−0.07, 0.171) | 0.41 |
| | Offspring | Duos | 1.097 (0.946, 1.248) | 6.0e−46 | 0.313 (0.282, 0.344) | 2.2e−87 | 0.239 (0.077, 0.401) | 0.004 |
| | | Trios | 1.143 (0.895, 1.391) | 1.5e−19 | 0.344 (0.291, 0.397) | 1.7e−37 | 0.22 (0.023, 0.418) | 0.029 |
| TC (mmol/L) | Maternal | Duos | −0.023 (−0.182, 0.136) | 0.78 | 0.023 (−0.011, 0.057) | 0.192 | −0.023 (−0.104, 0.059) | 0.588 |
| | | Trios | −0.012 (−0.208, 0.183) | 0.902 | 0.029 (−0.012, 0.07) | 0.168 | −0.018 (−0.118, 0.082) | 0.724 |
| | Offspring | Duos | 0.155 (−0.035, 0.346) | 0.11 | −0.004 (−0.031, 0.023) | 0.768 | 0.036 (−0.008, 0.079) | 0.107 |
| | | Trios | 0.136 (−0.101, 0.373) | 0.26 | −0.016 (−0.061, 0.028) | 0.464 | 0.027 (−0.071, 0.124) | 0.593 |
| HDL−C (mmol/L) | Maternal | Duos | −0.1 (−0.245, 0.045) | 0.176 | 0 (−0.032, 0.032) | 0.994 | −0.002 (−0.071, 0.067) | 0.95 |
| | | Trios | −0.118 (−0.294, 0.058) | 0.189 | −0.003 (−0.042, 0.036) | 0.874 | −0.022 (−0.106, 0.061) | 0.6 |
| | Offspring | Duos | 0.021 (−0.141, 0.183) | 0.801 | −0.085 (−0.114, −0.056) | 9.0e−09 | 0.011 (−0.051, 0.072) | 0.731 |
| | | Trios | 0.058 (−0.172, 0.289) | 0.62 | −0.079 (−0.123, −0.034) | 5.4e−04 | 0.051 (−0.039, 0.141) | 0.263 |
| LDL−C (mmol/L) | Maternal | Duos | −0.048 (−0.205, 0.109) | 0.551 | −0.007 (−0.04, 0.026) | 0.673 | −0.025 (−0.112, 0.062) | 0.57 |
| | | Trios | −0.04 (−0.233, 0.153) | 0.686 | −0.012 (−0.052, 0.028) | 0.555 | −0.008 (−0.117, 0.101) | 0.89 |
| | Offspring | Duos | 0.155 (−0.002, 0.311) | 0.053 | 0.008 (−0.036, 0.052) | 0.713 | 0.033 (−0.01, 0.075) | 0.13 |
| | | Trios | 0.14 (−0.071, 0.351) | 0.193 | 0.019 (−0.037, 0.074) | 0.507 | −0.002 (−0.098, 0.094) | 0.961 |
| TG (mmol/L) | Maternal | Duos | 0.024 (−0.114, 0.161) | 0.736 | 0.029 (−0.001, 0.06) | 0.062 | −0.069 (−0.152, 0.013) | 0.1 |
| | | Trios | 0.058 (−0.123, 0.24) | 0.529 | 0.038 (0.001, 0.076) | 0.047 | −0.081 (−0.182, 0.019) | 0.113 |
| | Offspring | Duos | 0.083 (−0.073, 0.239) | 0.295 | 0.096 (0.065, 0.127) | 7.8e−10 | 0.064 (0.019, 0.108) | 0.005 |
| | | Trios | 0.015 (−0.218, 0.247) | 0.901 | 0.078 (0.034, 0.121) | 4.6e−04 | 0.088 (−0.012, 0.188) | 0.084 |
| CRP (mg/L) | Maternal | Duos | 0.227 (0.058, 0.396) | 0.008 | 0.03 (−0.007, 0.067) | 0.109 | 0.05 (−0.031, 0.132) | 0.228 |
| | | Trios | 0.208 (0.001, 0.414) | 0.048 | 0.026 (−0.017, 0.07) | 0.238 | 0.054 (−0.045, 0.154) | 0.284 |
| | Offspring | Duos | −0.278 (−0.485, −0.071) | 0.009 | 0.005 (−0.03, 0.04) | 0.776 | −0.136 (−0.3, 0.028) | 0.104 |
| | | Trios | −0.239 (−0.513, 0.035) | 0.088 | 0.013 (−0.039, 0.065) | 0.634 | −0.145 (−0.341, 0.052) | 0.149 |

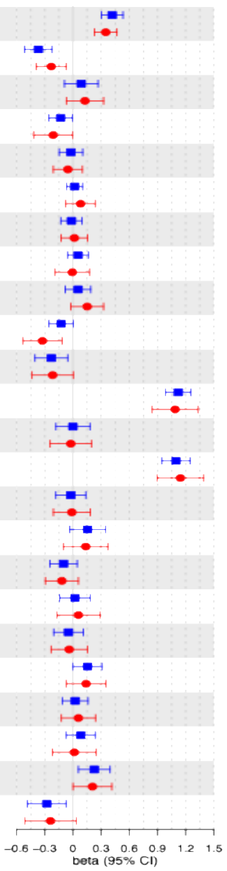

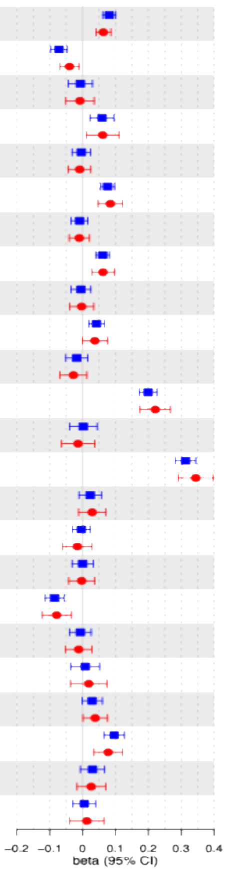

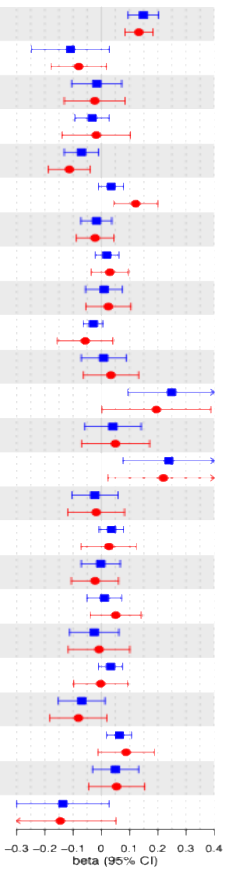

BMI (body mass index), WHR (waist-hip ratio), SBP (systolic blood pressure), DBP (diastolic blood pressure), HbA1c (glycated hemoglobin), TC (total cholesterol), HDL-C (high density lipoprotein cholesterol), LDL-C (low density lipoprotein cholesterol), TG (triglycerides), CRP (C-reactive protein). Birthweight: sex-stratified z-score. CRP: natural log transformed. Beta for binary exposures (type 2 diabetes and gestational diabetes): in log odds scale.

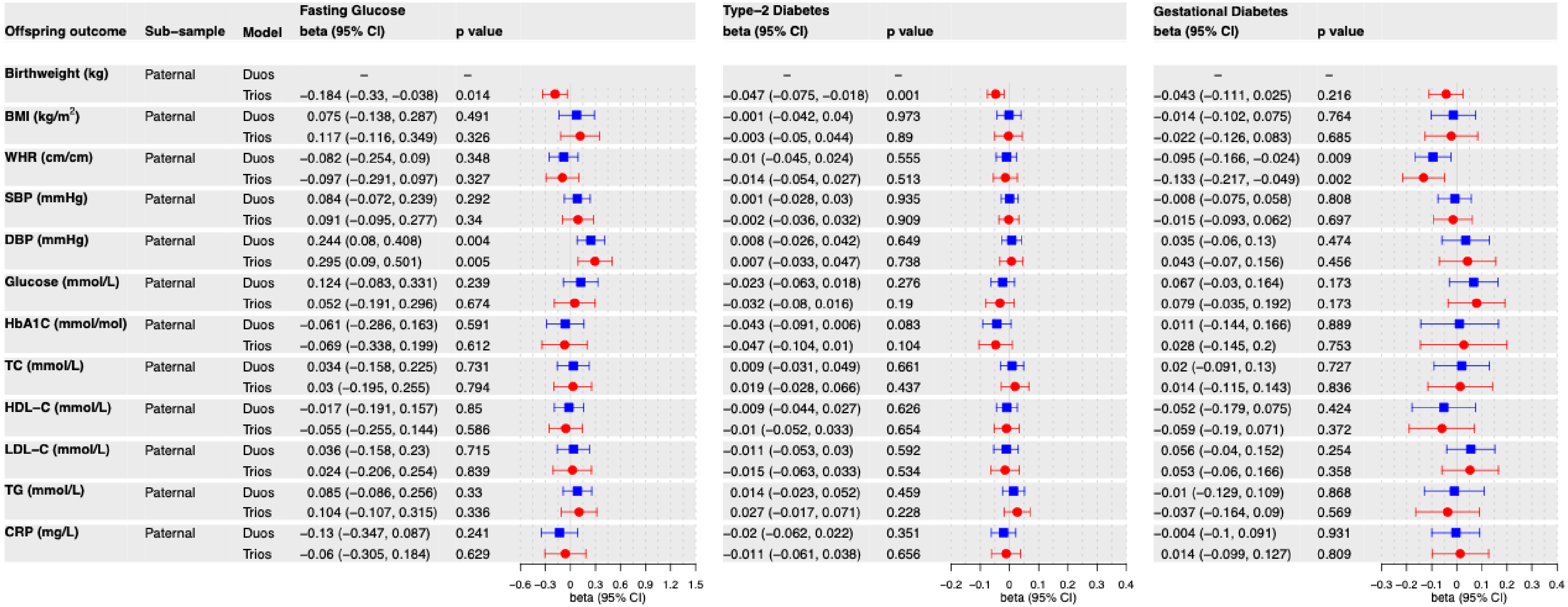

| Offspring outcome | Sub-sample | Model | Fasting Glucose beta (95% CI) | p value | Type-2 Diabetes beta (95% CI) | p value | Gestational Diabetes beta (95% CI) | p value |
|---|---|---|---|---|---|---|---|---|
| **Birthweight (kg)** | Paternal | Duos | – | – | – | – | – | – |
| | | Trios | −0.184 (−0.33, −0.038) | 0.014 | −0.047 (−0.075, −0.018) | 0.001 | −0.043 (−0.111, 0.025) | 0.216 |
| **BMI (kg/m²)** | Paternal | Duos | 0.075 (−0.138, 0.287) | 0.491 | −0.001 (−0.042, 0.04) | 0.973 | −0.014 (−0.102, 0.075) | 0.764 |
| | | Trios | 0.117 (−0.116, 0.349) | 0.326 | −0.003 (−0.05, 0.044) | 0.89 | −0.022 (−0.126, 0.083) | 0.685 |
| **WHR (cm/cm)** | Paternal | Duos | −0.082 (−0.254, 0.09) | 0.348 | −0.01 (−0.045, 0.024) | 0.555 | −0.095 (−0.166, −0.024) | 0.009 |
| | | Trios | −0.097 (−0.291, 0.097) | 0.327 | −0.014 (−0.054, 0.027) | 0.513 | −0.133 (−0.217, −0.049) | 0.002 |
| **SBP (mmHg)** | Paternal | Duos | 0.084 (−0.072, 0.239) | 0.292 | 0.001 (−0.028, 0.03) | 0.935 | −0.008 (−0.075, 0.058) | 0.808 |
| | | Trios | 0.091 (−0.095, 0.277) | 0.34 | −0.002 (−0.036, 0.032) | 0.909 | −0.015 (−0.093, 0.062) | 0.697 |
| **DBP (mmHg)** | Paternal | Duos | 0.244 (0.08, 0.408) | 0.004 | 0.008 (−0.026, 0.042) | 0.649 | 0.035 (−0.06, 0.13) | 0.474 |
| | | Trios | 0.295 (0.09, 0.501) | 0.005 | 0.007 (−0.033, 0.047) | 0.738 | 0.043 (−0.07, 0.156) | 0.456 |
| **Glucose (mmol/L)** | Paternal | Duos | 0.124 (−0.083, 0.331) | 0.239 | −0.023 (−0.063, 0.018) | 0.276 | 0.067 (−0.03, 0.164) | 0.173 |
| | | Trios | 0.052 (−0.191, 0.296) | 0.674 | −0.032 (−0.08, 0.016) | 0.19 | 0.079 (−0.035, 0.192) | 0.173 |
| **HbA1C (mmol/mol)** | Paternal | Duos | −0.061 (−0.286, 0.163) | 0.591 | −0.043 (−0.091, 0.006) | 0.083 | 0.011 (−0.144, 0.166) | 0.889 |
| | | Trios | −0.069 (−0.338, 0.199) | 0.612 | −0.047 (−0.104, 0.01) | 0.104 | 0.028 (−0.145, 0.2) | 0.753 |
| **TC (mmol/L)** | Paternal | Duos | 0.034 (−0.158, 0.225) | 0.731 | 0.009 (−0.031, 0.049) | 0.661 | 0.02 (−0.091, 0.13) | 0.727 |
| | | Trios | 0.03 (−0.195, 0.255) | 0.794 | 0.019 (−0.028, 0.066) | 0.437 | 0.014 (−0.115, 0.143) | 0.836 |
| **HDL-C (mmol/L)** | Paternal | Duos | −0.017 (−0.191, 0.157) | 0.85 | −0.009 (−0.044, 0.027) | 0.626 | −0.052 (−0.179, 0.075) | 0.424 |
| | | Trios | −0.055 (−0.255, 0.144) | 0.586 | −0.01 (−0.052, 0.033) | 0.654 | −0.059 (−0.19, 0.071) | 0.372 |
| **LDL-C (mmol/L)** | Paternal | Duos | 0.036 (−0.158, 0.23) | 0.715 | −0.011 (−0.053, 0.03) | 0.592 | 0.056 (−0.04, 0.152) | 0.254 |
| | | Trios | 0.024 (−0.206, 0.254) | 0.839 | −0.015 (−0.063, 0.033) | 0.534 | 0.053 (−0.06, 0.166) | 0.358 |
| **TG (mmol/L)** | Paternal | Duos | 0.085 (−0.086, 0.256) | 0.33 | 0.014 (−0.023, 0.052) | 0.459 | −0.01 (−0.129, 0.109) | 0.868 |
| | | Trios | 0.104 (−0.107, 0.315) | 0.336 | 0.027 (−0.017, 0.071) | 0.228 | −0.037 (−0.164, 0.09) | 0.569 |
| **CRP (mg/L)** | Paternal | Duos | −0.13 (−0.347, 0.087) | 0.241 | −0.02 (−0.062, 0.022) | 0.351 | −0.004 (−0.1, 0.091) | 0.931 |
| | | Trios | −0.06 (−0.305, 0.184) | 0.629 | −0.011 (−0.061, 0.038) | 0.656 | 0.014 (−0.099, 0.127) | 0.809 |

BMI (body mass index), WHR (waist-hip ratio), SBP (systolic blood pressure), DBP (diastolic blood pressure), HbA1c (glycated hemoglobin), TC (total cholesterol), HDL-C (high density lipoprotein cholesterol), LDL-C (low density lipoprotein cholesterol), TG (triglycerides), CRP (C-reactive protein). Birthweight: sex-stratified z-score. CRP: natural log transformed. Beta for binary exposures (type 2 diabetes and gestational diabetes): in log odds scale.

**Supplementary information S12. F-statistics for genetic variants of glycemic traits used in the intergenerational MR analysis in trios WLM.**

| Offspring outcome (units) | Maternal glycemia | | | Paternal glycemia | | | Offspring glycemia | | |
|---|---|---|---|---|---|---|---|---|---|
| | **Fasting glucose (SNPs)** | **Type 2 diabetes (SNPs)** | **Gestational diabetes (SNPs)** | **Fasting glucose (SNPs)** | **Type 2 diabetes (SNPs)** | **Gestational diabetes (SNPs)** | **Fasting glucose (SNPs)** | **Type 2 diabetes (SNPs)** | **Gestational diabetes (SNPs)** |
| Birthweight (kg) | 124.6 (56) | 103.1 (202) | 48.9 (7) | 124.6 (56) | 103.1 (202) | 48.9 (7) | 124.6 (56) | 103.1 (202) | 48.9 (7) |
| BMI (kg/m$^2$) | 126.0 (58) | 103.6 (200) | 50.9 (8) | 126.0 (58) | 103.6 (200) | 50.9 (8) | 126.0 (58) | 103.6 (200) | 50.9 (8) |
| WHR (cm/cm) | 121.4 (60) | 90.2 (207) | 50.9 (8) | 121.4 (60) | 90.2 (207) | 50.9 (8) | 121.4 (60) | 90.2 (207) | 50.9 (8) |
| SBP (mmHg) | 123.5 (60) | 104.6 (209) | 50.9 (8) | 123.5 (60) | 104.6 (209) | 50.9 (8) | 123.5 (60) | 104.6 (209) | 50.9 (8) |
| DBP (mmHg) | 123.7 (60) | 104.7 (209) | 50.8 (8) | 123.7 (60) | 104.7 (209) | 50.8 (8) | 123.7 (60) | 104.7 (209) | 50.8 (8) |
| Glucose (mmol/L) | 90.9 (56) | 85.7 (203) | 44.7 (7) | 90.9 (56) | 85.7 (203) | 44.7 (7) | 90.9 (56) | 85.7 (203) | 44.7 (7) |
| HbA1c (mmol/mol) | 121.4 (60) | 90.1 (207) | 44.7 (7) | 121.4 (60) | 90.1 (207) | 44.7 (7) | 121.4 (60) | 90.1 (207) | 44.7 (7) |
| TC (mmol/L) | 121.2 (59) | 103.5 (206) | 48.9 (7) | 121.2 (59) | 103.5 (206) | 48.9 (7) | 121.2 (59) | 103.5 (206) | 48.9 (7) |
| HDL-C (mmol/L) | 122.9 (60) | 104.7 (209) | 50.9 (8) | 122.9 (60) | 104.7 (209) | 50.9 (8) | 122.9 (60) | 104.7 (209) | 50.9 (8) |
| LDL-C (mmol/L) | 122.6 (58) | 103.3 (206) | 48.9 (7) | 122.6 (58) | 103.3 (206) | 48.9 (7) | 122.6 (58) | 103.3 (206) | 48.9 (7) |
| TG (mmol/L) | 118.8 (59) | 89.4 (207) | 41.3 (6) | 118.8 (59) | 89.4 (207) | 41.3 (6) | 118.8 (59) | 89.4 (207) | 41.3 (6) |
| CRP (mg/L) | 122.5 (61) | 104.3 (209) | 50.9 (8) | 122.5 (61) | 104.3 (209) | 50.9 (8) | 122.5 (61) | 104.3 (209) | 50.9 (8) |

BMI (body mass index), WHR (waist-hip ratio), SBP (systolic blood pressure), DBP (diastolic blood pressure), HbA1c (glycated hemoglobin), TC (total cholesterol), HDL-C (high density lipoprotein cholesterol), LDL-C (low density lipoprotein cholesterol), TG (triglycerides), CRP (C-reactive protein).

CRP: natural log transformed

Note: Mean F statistics across all genetic variants were calculated as F = mean(SNP-exposure effect size^2/standard error of SNP-exposure effect size^2). F-statistics were similar in unadjusted and adjusted MR estimates.

**Supplementary information S13. Sensitivity analysis for adjusted intergenerational MR estimates.**

**A. Maternal – adjusted intergenerational MR estimates in trios WLM**

| Offspring outcome | Sub-sample | Method | Fasting Glucose beta (95% CI) | p value | Type-2 Diabetes beta (95% CI) | p value | Gestational Diabetes beta (95% CI) | p value |
|---|---|---|---|---|---|---|---|---|
| BMI (kg/m$^2$) | Maternal | IVW | 0.128 (−0.07, 0.326) | 0.206 | −0.008 (−0.051, 0.035) | 0.727 | −0.023 (−0.131, 0.084) | 0.668 |
| | | MR Egger | 0.245 (−0.154, 0.644) | 0.235 | −0.067 (−0.153, 0.019) | 0.128 | −0.204 (−0.71, 0.302) | 0.459 |
| | | Weighted median | 0.164 (−0.144, 0.471) | 0.297 | −0.023 (−0.09, 0.044) | 0.502 | −0.036 (−0.169, 0.096) | 0.59 |
| | | Weighted mode | 0.159 (−0.159, 0.476) | 0.331 | −0.054 (−0.137, 0.029) | 0.203 | −0.062 (−0.323, 0.199) | 0.655 |
| WHR (cm/cm) | Maternal | IVW | −0.054 (−0.209, 0.101) | 0.495 | −0.009 (−0.044, 0.025) | 0.599 | −0.113 (−0.187, −0.039) | 0.003 |
| | | MR Egger | 0.012 (−0.299, 0.322) | 0.941 | −0.024 (−0.106, 0.058) | 0.571 | −0.227 (−0.565, 0.111) | 0.236 |
| | | Weighted median | 0.006 (−0.242, 0.253) | 0.964 | −0.022 (−0.076, 0.032) | 0.429 | −0.116 (−0.216, −0.017) | 0.022 |
| | | Weighted mode | −0.064 (−0.348, 0.221) | 0.662 | −0.016 (−0.11, 0.079) | 0.745 | −0.129 (−0.273, 0.015) | 0.123 |
| SBP (mmHg) | Maternal | IVW | 0.015 (−0.126, 0.156) | 0.838 | −0.01 (−0.041, 0.021) | 0.539 | −0.022 (−0.088, 0.044) | 0.515 |
| | | MR Egger | −0.15 (−0.433, 0.134) | 0.305 | −0.042 (−0.105, 0.021) | 0.193 | 0.018 (−0.284, 0.321) | 0.909 |
| | | Weighted median | −0.076 (−0.296, 0.144) | 0.5 | −0.006 (−0.066, 0.054) | 0.843 | −0.032 (−0.114, 0.05) | 0.448 |
| | | Weighted mode | −0.103 (−0.323, 0.118) | 0.366 | −0.007 (−0.066, 0.051) | 0.805 | −0.034 (−0.151, 0.083) | 0.588 |
| DBP (mmHg) | Maternal | IVW | 0.152 (−0.022, 0.326) | 0.086 | −0.003 (−0.039, 0.034) | 0.892 | 0.025 (−0.055, 0.105) | 0.546 |
| | | MR Egger | 0.195 (−0.16, 0.55) | 0.287 | −0.054 (−0.127, 0.018) | 0.145 | −0.024 (−0.388, 0.34) | 0.902 |
| | | Weighted median | 0.081 (−0.168, 0.33) | 0.522 | −0.004 (−0.073, 0.064) | 0.905 | 0.031 (−0.071, 0.134) | 0.549 |
| | | Weighted mode | 0.034 (−0.241, 0.309) | 0.81 | −0.022 (−0.092, 0.047) | 0.529 | 0.048 (−0.084, 0.179) | 0.499 |
| Glucose (mmol/L) | Maternal | IVW | −0.216 (−0.435, 0.003) | 0.054 | −0.028 (−0.069, 0.013) | 0.174 | 0.034 (−0.063, 0.132) | 0.489 |
| | | MR Egger | 0.194 (−0.312, 0.701) | 0.456 | −0.027 (−0.128, 0.074) | 0.6 | 0.095 (−0.416, 0.606) | 0.731 |
| | | Weighted median | −0.126 (−0.436, 0.184) | 0.424 | −0.017 (−0.081, 0.046) | 0.594 | 0.023 (−0.103, 0.15) | 0.717 |
| | | Weighted mode | −0.011 (−0.427, 0.405) | 0.958 | 0.002 (−0.115, 0.119) | 0.97 | 0.029 (−0.136, 0.194) | 0.743 |
| HbA1C (mmol/mol) | Maternal | IVW | −0.023 (−0.242, 0.196) | 0.839 | −0.014 (−0.064, 0.037) | 0.597 | 0.05 (−0.07, 0.171) | 0.41 |
| | | MR Egger | 0.06 (−0.38, 0.5) | 0.79 | 0.064 (−0.057, 0.185) | 0.303 | −0.045 (−0.672, 0.582) | 0.894 |
| | | Weighted median | −0.102 (−0.457, 0.252) | 0.572 | 0.005 (−0.076, 0.086) | 0.906 | 0.076 (−0.082, 0.233) | 0.345 |
| | | Weighted mode | −0.049 (−0.487, 0.39) | 0.829 | 0.119 (−0.044, 0.283) | 0.153 | 0.087 (−0.112, 0.286) | 0.425 |
| TC (mmol/L) | Maternal | IVW | −0.012 (−0.208, 0.183) | 0.902 | 0.029 (−0.012, 0.07) | 0.168 | −0.018 (−0.118, 0.082) | 0.724 |
| | | MR Egger | −0.011 (−0.402, 0.381) | 0.958 | −0.063 (−0.146, 0.019) | 0.133 | −0.088 (−0.509, 0.333) | 0.698 |
| | | Weighted median | 0.082 (−0.225, 0.389) | 0.601 | −0.003 (−0.069, 0.063) | 0.926 | −0.033 (−0.165, 0.099) | 0.623 |
| | | Weighted mode | −0.033 (−0.384, 0.318) | 0.855 | −0.037 (−0.119, 0.045) | 0.374 | −0.097 (−0.265, 0.072) | 0.305 |
| HDL−C (mmol/L) | Maternal | IVW | −0.118 (−0.294, 0.058) | 0.189 | −0.003 (−0.042, 0.036) | 0.874 | −0.022 (−0.106, 0.061) | 0.6 |
| | | MR Egger | −0.318 (−0.673, 0.037) | 0.085 | 0.023 (−0.056, 0.102) | 0.571 | 0.215 (−0.167, 0.596) | 0.313 |
| | | Weighted median | −0.139 (−0.411, 0.134) | 0.318 | 0.028 (−0.042, 0.098) | 0.43 | −0.011 (−0.117, 0.096) | 0.846 |
| | | Weighted mode | −0.185 (−0.442, 0.072) | 0.163 | −0.001 (−0.076, 0.075) | 0.985 | 0.031 (−0.12, 0.182) | 0.7 |
| LDL−C (mmol/L) | Maternal | IVW | −0.04 (−0.233, 0.153) | 0.686 | −0.012 (−0.052, 0.028) | 0.555 | −0.008 (−0.117, 0.101) | 0.89 |
| | | MR Egger | 0.053 (−0.333, 0.439) | 0.789 | −0.081 (−0.162, 0) | 0.052 | −0.115 (−0.61, 0.38) | 0.667 |
| | | Weighted median | 0.047 (−0.245, 0.339) | 0.753 | −0.054 (−0.118, 0.01) | 0.1 | 0.008 (−0.124, 0.14) | 0.902 |
| | | Weighted mode | −0.02 (−0.351, 0.31) | 0.905 | −0.084 (−0.166, −0.003) | 0.045 | 0.018 (−0.213, 0.25) | 0.882 |
| TG (mmol/L) | Maternal | IVW | 0.058 (−0.123, 0.24) | 0.529 | 0.038 (0.001, 0.076) | 0.047 | −0.081 (−0.182, 0.019) | 0.113 |
| | | MR Egger | 0.224 (−0.137, 0.585) | 0.229 | −0.056 (−0.146, 0.033) | 0.219 | −0.134 (−0.61, 0.342) | 0.611 |
| | | Weighted median | 0.094 (−0.167, 0.354) | 0.48 | 0.012 (−0.046, 0.07) | 0.691 | −0.117 (−0.241, 0.007) | 0.064 |
| | | Weighted mode | 0.138 (−0.137, 0.413) | 0.329 | 0.002 (−0.081, 0.085) | 0.962 | −0.117 (−0.273, 0.038) | 0.199 |
| CRP (mg/L) | Maternal | IVW | 0.208 (0.001, 0.414) | 0.048 | 0.026 (−0.017, 0.07) | 0.238 | 0.054 (−0.045, 0.154) | 0.284 |
| | | MR Egger | 0.106 (−0.312, 0.523) | 0.622 | −0.009 (−0.098, 0.08) | 0.848 | 0.021 (−0.431, 0.473) | 0.931 |
| | | Weighted median | 0.164 (−0.179, 0.508) | 0.348 | 0.068 (−0.012, 0.147) | 0.096 | 0.089 (−0.047, 0.225) | 0.197 |
| | | Weighted mode | 0.095 (−0.264, 0.455) | 0.606 | 0.058 (−0.029, 0.146) | 0.194 | 0.119 (−0.066, 0.304) | 0.249 |
| Birthweight (kg) | Maternal | IVW | 0.349 (0.228, 0.469) | 1.3e−08 | 0.063 (0.041, 0.086) | 3.9e−08 | 0.133 (0.084, 0.183) | 1.4e−07 |
| | | MR Egger | 0.511 (0.289, 0.734) | 3.5e−05 | 0.109 (0.068, 0.15) | 5.2e−07 | 0.27 (0.094, 0.446) | 0.03 |
| | | Weighted median | 0.461 (0.337, 0.585) | 3.0e−13 | 0.089 (0.059, 0.12) | 8.5e−09 | 0.138 (0.088, 0.187) | 6.3e−08 |
| | | Weighted mode | 0.462 (0.338, 0.586) | 1.2e−09 | 0.093 (0.067, 0.118) | 1.7e−11 | 0.136 (0.082, 0.191) | 0.003 |

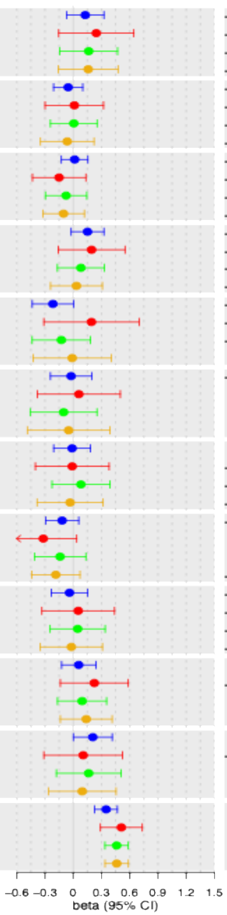

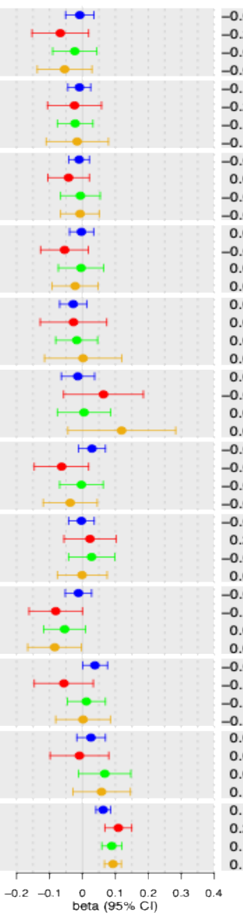

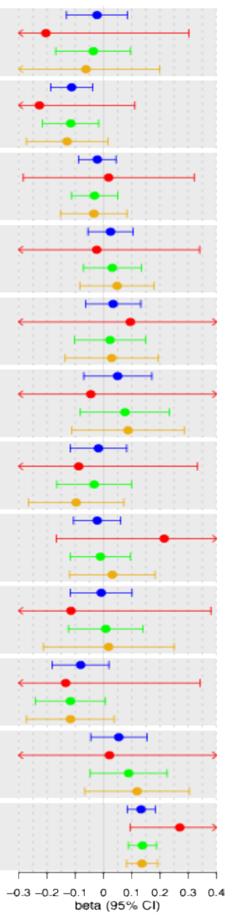

BMI (body mass index), WHR (waist-hip ratio), SBP (systolic blood pressure), DBP (diastolic blood pressure), HbA1c (glycated hemoglobin), TC (total cholesterol), HDL-C (high density lipoprotein cholesterol), LDL-C (low density lipoprotein cholesterol), TG (triglycerides), CRP (C-reactive protein). Birthweight: sex-stratified z-score. CRP: natural log transformed. Beta for binary exposures (type 2 diabetes and gestational diabetes): in log odds scale.

### B. Offspring – adjusted intergenerational MR estimates in trios WLM

| Offspring outcome | Sub-sample | Method | Fasting Glucose beta (95% CI) | p value | Type-2 Diabetes beta (95% CI) | p value | Gestational Diabetes beta (95% CI) | p value |
|---|---|---|---|---|---|---|---|---|
| Birthweight (kg) | Offspring | IVW | −0.231 (−0.391, −0.071) | 0.005 | −0.04 (−0.069, −0.011) | 0.008 | −0.08 (−0.178, 0.019) | 0.112 |
| | | MR Egger | −0.257 (−0.563, 0.048) | 0.105 | −0.085 (−0.139, −0.03) | 0.003 | 0.045 (−0.359, 0.448) | 0.836 |
| | | Weighted median | −0.349 (−0.509, −0.189) | 1.9e−05 | −0.021 (−0.063, 0.021) | 0.327 | −0.048 (−0.113, 0.018) | 0.153 |
| | | Weighted mode | −0.32 (−0.502, −0.138) | 0.001 | −0.04 (−0.08, 0) | 0.052 | −0.037 (−0.103, 0.029) | 0.316 |
| BMI (kg/m²) | Offspring | IVW | −0.21 (−0.416, −0.003) | 0.046 | 0.061 (0.011, 0.111) | 0.017 | −0.018 (−0.139, 0.103) | 0.77 |
| | | MR Egger | −0.333 (−0.749, 0.084) | 0.123 | −0.029 (−0.129, 0.071) | 0.572 | −0.101 (−0.689, 0.488) | 0.749 |
| | | Weighted median | −0.162 (−0.452, 0.128) | 0.273 | 0.015 (−0.072, 0.103) | 0.729 | 0.003 (−0.131, 0.137) | 0.963 |
| | | Weighted mode | −0.161 (−0.477, 0.154) | 0.32 | −0.005 (−0.088, 0.078) | 0.907 | 0.03 (−0.161, 0.221) | 0.766 |
| WHR (cm/cm) | Offspring | IVW | 0.081 (−0.078, 0.239) | 0.318 | 0.084 (0.047, 0.121) | 7.4e−06 | 0.122 (0.044, 0.2) | 0.002 |
| | | MR Egger | −0.031 (−0.348, 0.286) | 0.849 | 0.023 (−0.064, 0.111) | 0.603 | 0.111 (−0.272, 0.494) | 0.589 |
| | | Weighted median | 0.062 (−0.182, 0.307) | 0.618 | 0.095 (0.041, 0.149) | 5.6e−04 | 0.104 (0.008, 0.2) | 0.034 |
| | | Weighted mode | 0.091 (−0.218, 0.4) | 0.565 | 0.089 (0.003, 0.176) | 0.045 | 0.11 (−0.009, 0.23) | 0.114 |
| SBP (mmHg) | Offspring | IVW | −0.007 (−0.192, 0.178) | 0.941 | 0.062 (0.028, 0.097) | 3.7e−04 | 0.03 (−0.036, 0.096) | 0.372 |
| | | MR Egger | 0.108 (−0.269, 0.485) | 0.578 | 0.053 (−0.017, 0.122) | 0.14 | 0.105 (−0.203, 0.414) | 0.528 |
| | | Weighted median | 0.152 (−0.091, 0.394) | 0.22 | 0.051 (−0.007, 0.109) | 0.084 | 0.054 (−0.03, 0.139) | 0.209 |
| | | Weighted mode | 0.247 (0.001, 0.494) | 0.054 | 0.06 (−0.001, 0.122) | 0.056 | 0.059 (−0.051, 0.17) | 0.329 |
| DBP (mmHg) | Offspring | IVW | −0.324 (−0.533, −0.114) | 0.002 | 0.037 (−0.001, 0.076) | 0.056 | −0.057 (−0.156, 0.041) | 0.253 |
| | | MR Egger | −0.498 (−0.923, −0.073) | 0.025 | 0.017 (−0.061, 0.095) | 0.665 | −0.031 (−0.514, 0.452) | 0.903 |
| | | Weighted median | −0.23 (−0.492, 0.031) | 0.085 | 0.003 (−0.06, 0.065) | 0.934 | −0.081 (−0.186, 0.024) | 0.131 |
| | | Weighted mode | −0.257 (−0.534, 0.019) | 0.073 | 0.008 (−0.068, 0.085) | 0.833 | −0.076 (−0.213, 0.061) | 0.314 |
| Glucose (mmol/L) | Offspring | IVW | 1.087 (0.842, 1.332) | 3.6e−18 | 0.221 (0.174, 0.267) | 7.2e−21 | 0.195 (0.002, 0.388) | 0.047 |
| | | MR Egger | 0.996 (0.414, 1.579) | 0.001 | 0.255 (0.141, 0.368) | 1.9e−05 | 0.045 (−1.055, 1.144) | 0.94 |
| | | Weighted median | 1.13 (0.779, 1.481) | 2.7e−10 | 0.21 (0.142, 0.278) | 1.5e−09 | 0.144 (0.011, 0.277) | 0.034 |
| | | Weighted mode | 1.205 (0.787, 1.623) | 5.8e−07 | 0.171 (0.044, 0.297) | 0.009 | 0.102 (−0.082, 0.287) | 0.319 |
| HbA1C (mmol/mol) | Offspring | IVW | 1.143 (0.895, 1.391) | 1.5e−19 | 0.344 (0.291, 0.397) | 1.7e−37 | 0.22 (0.023, 0.418) | 0.029 |
| | | MR Egger | 1.29 (0.79, 1.79) | 4.6e−06 | 0.31 (0.183, 0.436) | 3.0e−06 | 0.416 (−0.7, 1.531) | 0.498 |
| | | Weighted median | 1.118 (0.767, 1.47) | 4.5e−10 | 0.323 (0.249, 0.398) | 1.6e−17 | 0.111 (−0.045, 0.266) | 0.159 |
| | | Weighted mode | 1.159 (0.714, 1.604) | 3.7e−06 | 0.265 (0.077, 0.452) | 0.006 | 0.091 (−0.088, 0.271) | 0.358 |
| TC (mmol/L) | Offspring | IVW | 0.136 (−0.101, 0.373) | 0.26 | −0.016 (−0.061, 0.028) | 0.464 | 0.027 (−0.071, 0.124) | 0.593 |
| | | MR Egger | −0.027 (−0.504, 0.451) | 0.913 | 0.041 (−0.048, 0.129) | 0.371 | 0.038 (−0.374, 0.449) | 0.865 |
| | | Weighted median | 0.034 (−0.264, 0.332) | 0.825 | 0.036 (−0.031, 0.103) | 0.296 | 0.071 (−0.048, 0.191) | 0.242 |
| | | Weighted mode | −0.069 (−0.421, 0.284) | 0.703 | 0.035 (−0.047, 0.117) | 0.398 | 0.086 (−0.083, 0.255) | 0.358 |
| HDL-C (mmol/L) | Offspring | IVW | 0.058 (−0.172, 0.289) | 0.62 | −0.079 (−0.123, −0.034) | 5.4e−04 | 0.051 (−0.039, 0.141) | 0.263 |
| | | MR Egger | 0.367 (−0.094, 0.829) | 0.124 | 0.022 (−0.068, 0.111) | 0.636 | −0.023 (−0.46, 0.414) | 0.92 |
| | | Weighted median | 0.028 (−0.253, 0.308) | 0.846 | −0.017 (−0.084, 0.05) | 0.623 | 0.004 (−0.107, 0.115) | 0.945 |
| | | Weighted mode | 0.025 (−0.237, 0.287) | 0.85 | −0.014 (−0.086, 0.058) | 0.706 | −0.018 (−0.167, 0.131) | 0.819 |
| LDL-C (mmol/L) | Offspring | IVW | 0.14 (−0.071, 0.351) | 0.193 | 0.019 (−0.037, 0.074) | 0.507 | −0.002 (−0.098, 0.094) | 0.961 |
| | | MR Egger | −0.098 (−0.519, 0.323) | 0.65 | 0.014 (−0.098, 0.126) | 0.806 | 0.031 (−0.373, 0.436) | 0.885 |
| | | Weighted median | −0.084 (−0.381, 0.212) | 0.577 | 0.058 (−0.015, 0.132) | 0.121 | −0.027 (−0.155, 0.1) | 0.673 |
| | | Weighted mode | 0.014 (−0.33, 0.357) | 0.939 | 0.051 (−0.028, 0.13) | 0.207 | −0.051 (−0.251, 0.149) | 0.637 |
| TG (mmol/L) | Offspring | IVW | 0.015 (−0.218, 0.247) | 0.901 | 0.078 (0.034, 0.121) | 4.6e−04 | 0.088 (−0.012, 0.188) | 0.084 |
| | | MR Egger | −0.241 (−0.701, 0.219) | 0.309 | 0.089 (−0.016, 0.193) | 0.098 | 0.039 (−0.44, 0.518) | 0.881 |
| | | Weighted median | −0.137 (−0.41, 0.137) | 0.327 | 0.084 (0.023, 0.145) | 0.007 | 0.09 (−0.038, 0.218) | 0.169 |
| | | Weighted mode | −0.042 (−0.33, 0.246) | 0.775 | 0.073 (−0.031, 0.177) | 0.173 | 0.096 (−0.079, 0.271) | 0.331 |
| CRP (mg/L) | Offspring | IVW | −0.239 (−0.513, 0.035) | 0.088 | 0.013 (−0.039, 0.065) | 0.634 | −0.145 (−0.341, 0.052) | 0.149 |
| | | MR Egger | −0.033 (−0.59, 0.524) | 0.908 | −0.052 (−0.157, 0.053) | 0.331 | −0.082 (−1.046, 0.882) | 0.873 |
| | | Weighted median | 0.016 (−0.335, 0.367) | 0.929 | −0.034 (−0.105, 0.037) | 0.346 | −0.108 (−0.251, 0.036) | 0.14 |
| | | Weighted mode | 0.041 (−0.27, 0.352) | 0.798 | −0.045 (−0.137, 0.047) | 0.339 | −0.04 (−0.224, 0.143) | 0.679 |

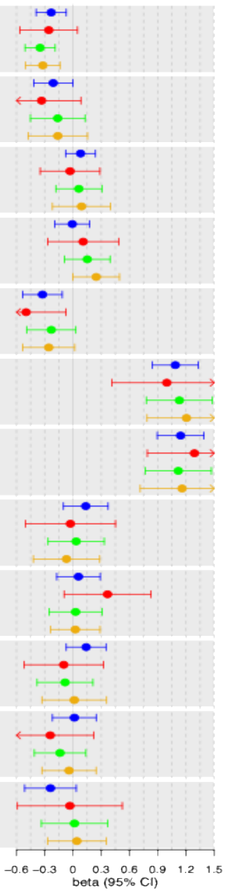

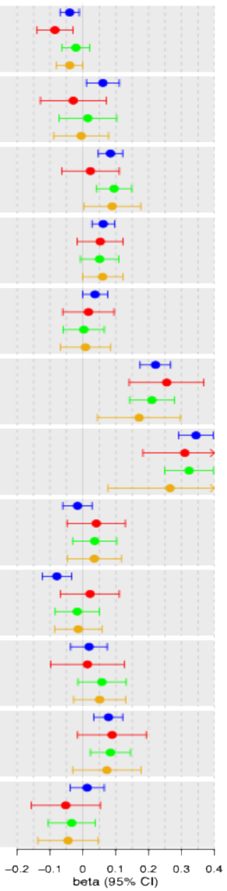

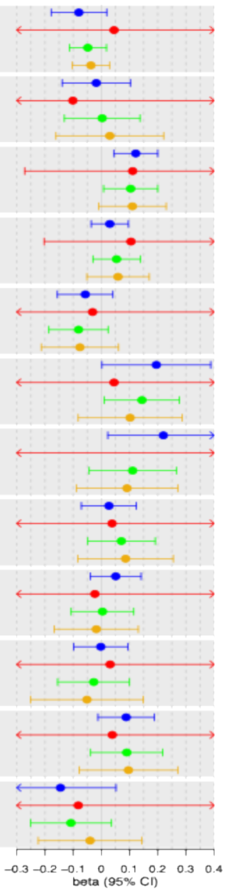

BMI (body mass index), WHR (waist-hip ratio), SBP (systolic blood pressure), DBP (diastolic blood pressure), HbA1c (glycated hemoglobin), TC (total cholesterol), HDL-C (high density lipoprotein cholesterol), LDL-C (low density lipoprotein cholesterol), TG (triglycerides), CRP (C-reactive protein). Birthweight: sex-stratified z-score. CRP: natural log transformed. Beta for binary exposures (type 2 diabetes and gestational diabetes): in log odds scale.

### C. Paternal – adjusted intergenerational MR estimates in trios WLM

| Offspring outcome | Sub-sample | Method | Fasting Glucose beta (95% CI) | p value | Type-2 Diabetes beta (95% CI) | p value | Gestational Diabetes beta (95% CI) | p value |
|---|---|---|---|---|---|---|---|---|
| Birthweight (kg) | Paternal | IVW | –0.184 (–0.33, –0.038) | 0.014 | –0.047 (–0.075, –0.018) | 0.001 | –0.043 (–0.111, 0.025) | 0.216 |
| | | MR Egger | –0.147 (–0.433, 0.139) | 0.318 | 0.015 (–0.038, 0.068) | 0.58 | 0.127 (–0.128, 0.382) | 0.373 |
| | | Weighted median | –0.142 (–0.324, 0.04) | 0.126 | –0.02 (–0.061, 0.022) | 0.358 | –0.046 (–0.134, 0.041) | 0.3 |
| | | Weighted mode | –0.13 (–0.333, 0.073) | 0.215 | 0.013 (–0.035, 0.061) | 0.595 | 0.059 (–0.112, 0.23) | 0.522 |
| BMI (kg/m$^2$) | Paternal | IVW | 0.117 (–0.116, 0.349) | 0.326 | –0.003 (–0.05, 0.044) | 0.89 | –0.022 (–0.126, 0.083) | 0.685 |
| | | MR Egger | 0.26 (–0.209, 0.73) | 0.282 | –0.028 (–0.123, 0.067) | 0.569 | –0.013 (–0.492, 0.466) | 0.96 |
| | | Weighted median | 0.147 (–0.206, 0.5) | 0.414 | –0.029 (–0.122, 0.063) | 0.532 | –0.056 (–0.192, 0.08) | 0.421 |
| | | Weighted mode | 0.177 (–0.213, 0.568) | 0.378 | –0.003 (–0.089, 0.084) | 0.947 | –0.07 (–0.296, 0.155) | 0.56 |
| WHR (cm/cm) | Paternal | IVW | –0.097 (–0.291, 0.097) | 0.327 | –0.014 (–0.054, 0.027) | 0.513 | –0.133 (–0.217, –0.049) | 0.002 |
| | | MR Egger | –0.048 (–0.439, 0.344) | 0.812 | –0.047 (–0.143, 0.05) | 0.347 | –0.078 (–0.458, 0.303) | 0.702 |
| | | Weighted median | –0.097 (–0.407, 0.214) | 0.541 | –0.018 (–0.081, 0.044) | 0.568 | –0.137 (–0.245, –0.03) | 0.012 |
| | | Weighted mode | –0.079 (–0.399, 0.241) | 0.629 | –0.026 (–0.144, 0.092) | 0.669 | –0.136 (–0.267, –0.004) | 0.083 |
| SBP (mmHg) | Paternal | IVW | 0.091 (–0.095, 0.277) | 0.34 | –0.002 (–0.036, 0.032) | 0.909 | –0.015 (–0.093, 0.062) | 0.697 |
| | | MR Egger | 0.031 (–0.348, 0.411) | 0.873 | –0.045 (–0.114, 0.024) | 0.2 | –0.143 (–0.497, 0.21) | 0.457 |
| | | Weighted median | –0.003 (–0.253, 0.246) | 0.98 | –0.019 (–0.086, 0.048) | 0.58 | –0.037 (–0.144, 0.07) | 0.502 |
| | | Weighted mode | –0.08 (–0.368, 0.209) | 0.591 | –0.023 (–0.092, 0.047) | 0.527 | –0.045 (–0.187, 0.096) | 0.549 |
| DBP (mmHg) | Paternal | IVW | 0.295 (0.09, 0.501) | 0.005 | 0.007 (–0.033, 0.047) | 0.738 | 0.043 (–0.07, 0.156) | 0.456 |
| | | MR Egger | 0.529 (0.114, 0.944) | 0.015 | –0.053 (–0.134, 0.029) | 0.208 | –0.094 (–0.637, 0.449) | 0.745 |
| | | Weighted median | 0.305 (–0.01, 0.62) | 0.057 | –0.007 (–0.093, 0.079) | 0.878 | 0.028 (–0.099, 0.154) | 0.668 |
| | | Weighted mode | 0.287 (–0.019, 0.592) | 0.071 | 0 (–0.093, 0.092) | 0.995 | 0.029 (–0.124, 0.181) | 0.725 |
| Glucose (mmol/L) | Paternal | IVW | 0.052 (–0.191, 0.296) | 0.674 | –0.032 (–0.08, 0.016) | 0.19 | 0.079 (–0.035, 0.192) | 0.173 |
| | | MR Egger | 0.088 (–0.491, 0.666) | 0.767 | 0.006 (–0.111, 0.124) | 0.916 | 0.179 (–0.414, 0.773) | 0.58 |
| | | Weighted median | 0.059 (–0.346, 0.464) | 0.776 | –0.034 (–0.11, 0.042) | 0.382 | 0.026 (–0.123, 0.175) | 0.73 |
| | | Weighted mode | 0.003 (–0.5, 0.505) | 0.992 | –0.061 (–0.215, 0.092) | 0.435 | –0.036 (–0.301, 0.229) | 0.8 |
| HbA1C (mmol/mol) | Paternal | IVW | –0.069 (–0.338, 0.199) | 0.612 | –0.047 (–0.104, 0.01) | 0.104 | 0.028 (–0.145, 0.2) | 0.753 |
| | | MR Egger | –0.364 (–0.902, 0.174) | 0.19 | 0 (–0.136, 0.137) | 0.994 | –0.742 (–1.446, –0.037) | 0.094 |
| | | Weighted median | –0.067 (–0.447, 0.313) | 0.73 | –0.032 (–0.114, 0.051) | 0.452 | –0.04 (–0.216, 0.136) | 0.654 |
| | | Weighted mode | –0.233 (–0.7, 0.233) | 0.331 | –0.029 (–0.189, 0.131) | 0.723 | –0.077 (–0.28, 0.127) | 0.488 |
| TC (mmol/L) | Paternal | IVW | 0.03 (–0.195, 0.255) | 0.794 | 0.019 (–0.028, 0.066) | 0.437 | 0.014 (–0.115, 0.143) | 0.836 |
| | | MR Egger | 0.183 (–0.271, 0.636) | 0.433 | –0.046 (–0.141, 0.048) | 0.339 | –0.044 (–0.636, 0.548) | 0.89 |
| | | Weighted median | 0.158 (–0.185, 0.502) | 0.365 | –0.042 (–0.133, 0.05) | 0.373 | –0.071 (–0.228, 0.086) | 0.373 |
| | | Weighted mode | 0.234 (–0.227, 0.695) | 0.324 | –0.036 (–0.131, 0.058) | 0.452 | –0.097 (–0.276, 0.083) | 0.332 |
| HDL–C (mmol/L) | Paternal | IVW | –0.055 (–0.255, 0.144) | 0.586 | –0.01 (–0.052, 0.033) | 0.654 | –0.059 (–0.19, 0.071) | 0.372 |
| | | MR Egger | –0.045 (–0.448, 0.359) | 0.828 | –0.067 (–0.153, 0.018) | 0.126 | –0.162 (–0.8, 0.475) | 0.635 |
| | | Weighted median | 0.086 (–0.232, 0.403) | 0.596 | –0.06 (–0.139, 0.019) | 0.135 | –0.111 (–0.245, 0.023) | 0.103 |
| | | Weighted mode | 0.153 (–0.22, 0.526) | 0.425 | –0.056 (–0.141, 0.028) | 0.194 | –0.125 (–0.282, 0.032) | 0.162 |
| LDL–C (mmol/L) | Paternal | IVW | 0.024 (–0.206, 0.254) | 0.839 | –0.015 (–0.063, 0.033) | 0.534 | 0.053 (–0.06, 0.166) | 0.358 |
| | | MR Egger | 0.139 (–0.326, 0.603) | 0.561 | –0.031 (–0.128, 0.066) | 0.535 | 0 (–0.496, 0.496) | 1 |
| | | Weighted median | 0.155 (–0.212, 0.521) | 0.408 | –0.033 (–0.122, 0.056) | 0.464 | –0.003 (–0.153, 0.147) | 0.968 |
| | | Weighted mode | 0.052 (–0.353, 0.456) | 0.804 | –0.029 (–0.117, 0.058) | 0.51 | –0.035 (–0.222, 0.151) | 0.724 |
| TG (mmol/L) | Paternal | IVW | 0.104 (–0.107, 0.315) | 0.336 | 0.027 (–0.017, 0.071) | 0.228 | –0.037 (–0.164, 0.09) | 0.569 |
| | | MR Egger | 0.271 (–0.15, 0.692) | 0.212 | –0.044 (–0.149, 0.06) | 0.406 | –0.038 (–0.708, 0.633) | 0.918 |
| | | Weighted median | 0.216 (–0.115, 0.546) | 0.2 | 0.009 (–0.059, 0.076) | 0.803 | –0.105 (–0.26, 0.05) | 0.184 |
| | | Weighted mode | 0.195 (–0.148, 0.539) | 0.27 | 0.059 (–0.062, 0.18) | 0.342 | –0.144 (–0.385, 0.098) | 0.297 |
| CRP (mg/L) | Paternal | IVW | –0.06 (–0.305, 0.184) | 0.629 | –0.011 (–0.061, 0.038) | 0.656 | 0.014 (–0.099, 0.127) | 0.809 |
| | | MR Egger | –0.279 (–0.777, 0.218) | 0.275 | 0 (–0.1, 0.1) | 0.996 | 0.091 (–0.422, 0.604) | 0.741 |
| | | Weighted median | –0.075 (–0.444, 0.294) | 0.69 | 0.021 (–0.072, 0.115) | 0.654 | 0.046 (–0.103, 0.195) | 0.547 |
| | | Weighted mode | –0.069 (–0.433, 0.295) | 0.713 | 0.012 (–0.086, 0.11) | 0.809 | 0.068 (–0.134, 0.27) | 0.528 |

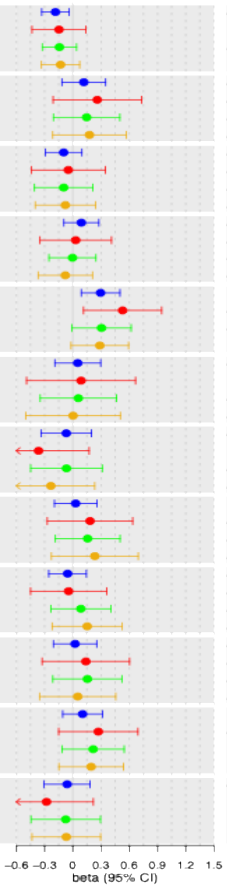

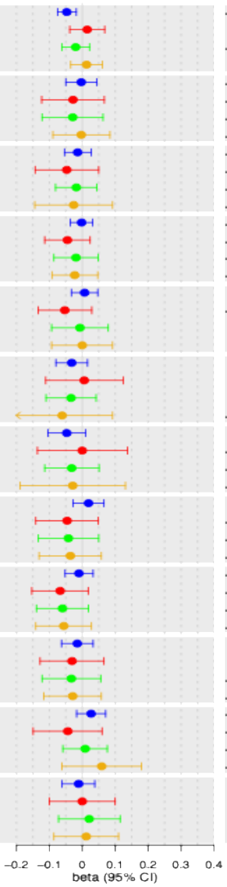

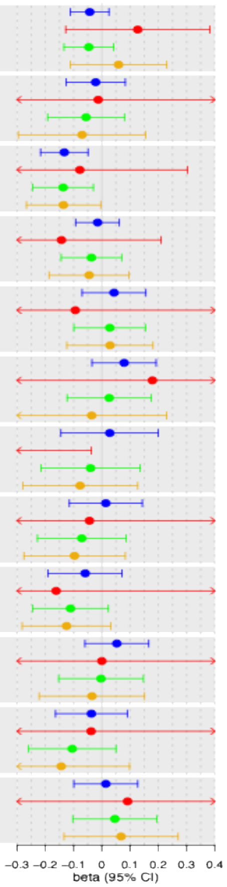

BMI (body mass index), WHR (waist-hip ratio), SBP (systolic blood pressure), DBP (diastolic blood pressure), HbA1c (glycated hemoglobin), TC (total cholesterol), HDL-C (high density lipoprotein cholesterol), LDL-C (low density lipoprotein cholesterol), TG (triglycerides), CRP (C-reactive protein). Birthweight: sex-stratified z-score. CRP: natural log transformed. Beta for binary exposures (type 2 diabetes and gestational diabetes): in log odds scale.

**Supplementary information S14. Test of bias due to horizontal pleiotropy of adjusted intergenerational MR analysis in trios WLM.**

### A. MR Egger intercept

| **Offspring outcome (units)** | **Maternal glycemia** | | | **Paternal glycemia** | | | **Offspring glycemia** | | |
|---|---|---|---|---|---|---|---|---|---|
| | **Fasting glucose** | **Type 2 diabetes** | **Gestational diabetes** | **Fasting glucose** | **Type 2 diabetes** | **Gestational diabetes** | **Fasting glucose** | **Type 2 diabetes** | **Gestational diabetes** |
| | **Intercept (*P*)** | **Intercept (*P*)** | **Intercept (*P*)** | **Intercept (*P*)** | **Intercept (*P*)** | **Intercept (*P*)** | **Intercept (*P*)** | **Intercept (*P*)** | **Intercept (*P*)** |
| Birthweight (kg) | -0.0043 (0.096) | -0.0030 (0.010) | -0.0184 (0.176) | -0.0010 (0.769) | -0.0039 (0.009) | -0.0228 (0.234) | 0.0007 (0.843) | 0.0029 (0.057) | -0.0168 (0.559) |
| BMI (kg/m$^2$) | -0.0136 (0.510) | 0.0168 (0.120) | 0.1085 (0.500) | -0.0166 (0.492) | 0.0068 (0.563) | -0.0053 (0.971) | 0.0142 (0.507) | 0.0253 (0.044) | 0.0497 (0.787) |
| WHR (cm/cm) | -0.0001 (0.634) | $7.2x10^{-05}$ (0.702) | 0.0013 (0.522) | -0.0001 (0.775) | 0.0001 (0.462) | -0.0006 (0.781) | 0.0002 (0.429) | 0.0003 (0.135) | 0.0001 (0.957) |
| SBP (mmHg) | 0.0839 (0.195) | 0.0409 (0.250) | -0.1087 (0.796) | 0.0303 (0.724) | 0.0549 (0.159) | 0.3439 (0.494) | -0.0584 (0.4953) | 0.0124 (0.751) | -0.2024 (0.641) |
| DBP (mmHg) | -0.0121 (0.788) | 0.0364 (0.110) | 0.0718 (0.798) | -0.0663 (0.209) | 0.0419 (0.101) | 0.2036 (0.630) | 0.0495 (0.358) | 0.0142 (0.560) | -0.0385 (0.917) |
| Glucose (mmol/L) | -0.0077 (0.085) | $-6.5x10^{-05}$ (0.977) | -0.0065 (0.822) | -0.0006 (0.894) | -0.0018 (0.485) | -0.0108 (0.749) | 0.0017 (0.738) | -0.0016 (0.521) | 0.0163 (0.795) |
| HbA1c (mmol/mol) | -0.0103 (0.672) | -0.0217 (0.169) | 0.0585 (0.773) | 0.0369 (0.221) | -0.0133 (0.451) | 0.4724 (0.081) | -0.0184 (0.509) | 0.0096 (0.557) | -0.1198 (0.740) |
| TC (mmol/L) | $-4.1x10^{-05}$ (0.992) | 0.0056 (0.012) | 0.0091 (0.750) | -0.0037 (0.449) | 0.0039 (0.123) | 0.0075 (0.851) | 0.0040 (0.444) | -0.0034 (0.147) | -0.0014 (0.959) |
| HDL-C (mmol/L) | 0.0018 (0.208) | -0.0006 (0.459) | -0.0115 (0.258) | $-9.9x10^{-05}$ (0.952) | 0.0013 (0.131) | 0.0050 (0.756) | -0.0028 (0.136) | -0.0023 (0.012) | 0.0036 (0.743) |
| LDL-C (mmol/L) | -0.0020 (0.589) | 0.0037 (0.057) | 0.0125 (0.679) | -0.0025 (0.579) | 0.0008 (0.718) | 0.0061 (0.837) | 0.0052 (0.206) | 0.0002 (0.924) | -0.0039 (0.873) |
| TG (mmol/L) | -0.0041 (0.301) | 0.0053 (0.023) | 0.0064 (0.835) | -0.0041 (0.371) | 0.0040 (0.143) | $8.4x10^{-05}$ (0.998) | 0.0064 (0.213) | -0.0006 (0.823) | -0.0014 (0.959) |
| CRP (mg/L) | 0.0025 | 0.0021 | 0.0043 | 0.0053 | -0.0006 | -0.0099 | -0.0050 | 0.0039 | -0.0080 |

| | (0.584) | (0.374) | (0.886) | (0.325) | (0.804) | (0.773) | (0.408) | (0.165) | (0.900) |
|---|---|---|---|---|---|---|---|---|---|

BMI (body mass index), WHR (waist-hip ratio), SBP (systolic blood pressure), DBP (diastolic blood pressure), HbA1c (glycated hemoglobin), TC (total cholesterol), HDL-C (high density lipoprotein cholesterol), LDL-C (low density lipoprotein cholesterol), TG (triglycerides), CRP (C-reactive protein), T (True directionality), F (False directionality).
Birthweight: sex-stratified z-score. CRP: natural log transformed. Note: *P* represents p value for MR Egger intercept.

### B. Between-SNPs MR heterogeneity test (Cochran´s Q statistics)

| **Offspring outcome (units)** | **Maternal glycemia** | | | **Paternal glycemia** | | | **Offspring glycemia** | | |
|---|---|---|---|---|---|---|---|---|---|
| | **Fasting glucose** | **Type 2 diabetes** | **Gestational diabetes** | **Fasting glucose** | **Type 2 diabetes** | **Gestational diabetes** | **Fasting glucose** | **Type 2 diabetes** | **Gestational diabetes** |
| | **Q statistics (*P*)** | **Q statistics (*P*)** | **Q statistics (*P*)** | **Q statistics (*P*)** | **Q statistics (*P*)** | **Q statistics (*P*)** | **Q statistics (*P*)** | **Q statistics (*P*)** | **Q statistics (*P*)** |
| Birthweight (kg) | 153.5 ($2.9x10^{-11}$) | 409.2 ($2.4x10^{-16}$) | 10.5 (0.101) | 89.5 (0.002) | 265.1 (0.001) | 8.1 (0.228) | 142.3 ($1.1x10^{-09}$) | 360.3 ($3.7x10^{-11}$) | 21.3 (0.001) |
| BMI (kg/m$^2$) | 60.0 (0.366) | 214.3 (0.216) | 9.5 (0.217) | 64.0 (0.244) | 201.2 (0.441) | 6.1 (0.526) | 69.3 (0.126) | 311.3 ($6.0x10^{-07}$) | 12.8 ($7.5x10^{-02}$) |
| WHR (cm/cm) | 52.1 (0.725) | 199.4 (0.616) | 4.5 (0.709) | 69.1 (0.172) | 203.269 (0.540) | 5.0 (0.657) | 63.9 (0.306) | 239.6 (0.054) | 8.327 (0.304) |
| SBP (mmHg) | 61.2 (0.394) | 237.5 (0.077) | 2.1 (0.949) | 77.2 (0.055) | 208.1 (0.484) | 4.1 (0.766) | 106.4 ($1.5x10^{-04}$) | 294.0 ($7.9x10^{-05}$) | 6.5 (0.473) |
| DBP (mmHg) | 64.5 (0.290) | 220.6 (0.261) | 4.1 (0.762) | 67.3 (0.213) | 205.7 (0.530) | 10.4 (0.163) | 96.2 (0.001) | 261.3 ($7.1x10^{-03}$) | 10.9 (0.140) |
| Glucose (mmol/L) | 61.3 (0.257) | 184.5 (0.805) | 4.2 (0.641) | 55.4 (0.457) | 189.8 (0.720) | 5.1 (0.529) | 77.9 ($2.2x10^{-02}$) | 261.0 (0.003) | 23.9 ($5.3x10^{-04}$) |
| HbA1c (mmol/mol) | 58.0 (0.509) | 220.5 (0.231) | 2.8 (0.824) | 70.7 (0.140) | 221.3 (0.220) | 9.8 (0.132) | 81.5 ($2.7x10^{-02}$) | 257.1 ($8.8x10^{-03}$) | 17.4 ($7.6x10^{-03}$) |
| TC (mmol/L) | 48.4 (0.810) | 208.8 (0.412) | 5.0 (0.535) | 47.6 (0.832) | 198.8 (0.606) | 7.5 (0.269) | 88.5 ($6.0x10^{-03}$) | 249.5 ($1.8x10^{-02}$) | 4.0 ($6.6x10^{-01}$) |
| HDL-C (mmol/L) | 52.5 (0.711) | 233.0 (0.112) | 3.6 (0.814) | 56.1 (0.580) | 214.1 (0.371) | 13.0 (0.070) | 108.6 ($8.7x10^{-05}$) | 329.1 ($1.7x10^{-07}$) | 8.5 ($2.8x10^{-01}$) |
| LDL-C (mmol/L) | 56.0 (0.512) | 203.9 (0.507) | 7.4 (0.284) | 60.5 (0.348) | 221.3 (0.206) | 5.4 (0.483) | 70.9 (0.101) | 405.3 ($2.7x10^{-15}$) | 5.0 (0.543) |
| TG (mmol/L) | 68.3 (0.165) | 217.1 (0.284) | 0.770 (0.978) | 65.2 (0.240) | 180.2 (0.902) | 5.6 (0.340) | 112.9 ($2.1x10^{-05}$) | 291.4 ($8.2x10^{-05}$) | 4.1 (0.527) |
| CRP (mg/L) | 54.2 (0.684) | 210.0 (0.447) | 3.1 (0.873) | 65.6 (0.286) | 200.8 (0.625) | 3.8 (0.797) | 112.4 ($4.7x10^{-05}$) | 315.6 ($2.1x10^{-06}$) | 29.0 ($1.4x10^{-04}$) |

BMI (body mass index), WHR (waist-hip ratio), SBP (systolic blood pressure), DBP (diastolic blood pressure), HbA1c (glycated hemoglobin), TC (total cholesterol), HDL-C (high density lipoprotein cholesterol), LDL-C (low density lipoprotein cholesterol), TG (triglycerides), CRP (C-reactive protein), T (True directionality), F (False directionality).
Birthweight: sex-stratified z-score. CRP: natural log transformed. Note: *P* represents p value for heterogeneity test.

### C. MR-PRESSO global test

| Offspring outcome (units) | Maternal glycemia | | | Paternal glycemia | | | Offspring glycemia | | |
|---|---|---|---|---|---|---|---|---|---|
| | Fasting glucose | Type 2 diabetes | Gestational diabetes | Fasting glucose | Type 2 diabetes | Gestational diabetes | Fasting glucose | Type 2 diabetes | Gestational diabetes |
| | ***P*** | ***P*** | ***P*** | ***P*** | ***P*** | ***P*** | ***P*** | ***P*** | ***P*** |
| Birthweight (kg) | <0.001 | <0.001 | 0.226 | 0.001 | <0.001 | 0.204 | <0.001 | <0.001 | 0.009 |
| BMI (kg/m$^2$) | 0.396 | 0.219 | 0.2 | 0.243 | 0.446 | 0.498 | 0.142 | <0.001 | 0.083 |
| WHR (cm/cm) | 0.722 | 0.617 | 0.723 | 0.181 | 0.555 | 0.717 | 0.286 | 0.057 | 0.389 |
| SBP (mmHg) | 0.38 | 0.09 | 0.955 | 0.06 | 0.487 | 0.787 | 0.001 | <0.001 | 0.491 |
| DBP (mmHg) | 0.28 | 0.274 | 0.784 | 0.205 | 0.522 | 0.228 | 0.001 | 0.007 | 0.194 |
| Glucose (mmol/L) | 0.239 | 0.809 | 0.681 | 0.432 | 0.72 | 0.478 | 0.02 | 0.006 | 0.002 |
| HbA1c (mmol/mol) | 0.465 | 0.216 | 0.85 | 0.122 | 0.241 | 0.172 | 0.026 | 0.011 | 0.023 |
| TC (mmol/L) | 0.808 | 0.396 | 0.5 | 0.82 | 0.618 | 0.301 | 0.006 | 0.022 | 0.678 |
| HDL-C (mmol/L) | 0.74 | 0.107 | 0.8 | 0.572 | 0.371 | 0.085 | <0.001 | <0.001 | 0.305 |
| LDL-C (mmol/L) | 0.499 | 0.508 | 0.245 | 0.357 | 0.189 | 0.492 | 0.098 | <0.001 | 0.511 |
| TG (mmol/L) | 0.171 | 0.279 | 0.978 | 0.258 | 0.9 | 0.379 | <0.001 | <0.001 | 0.579 |
| CRP (mg/L) | 0.694 | 0.47 | 0.862 | 0.299 | 0.65 | 0.771 | <0.001 | <0.001 | <0.001 |

BMI (body mass index), WHR (waist-hip ratio), SBP (systolic blood pressure), DBP (diastolic blood pressure), HbA1c (glycated hemoglobin), TC (total cholesterol), HDL-C (high density lipoprotein cholesterol), LDL-C (low density lipoprotein cholesterol), TG (triglycerides), CRP (C-reactive protein), T (True directionality), F (False directionality).
Birthweight: sex-stratified z-score. CRP: natural log transformed. Note: *P* represents p value for MR-PRESSO global test.

**Supplementary information S15. Comparing GWAS summary statistics between unadjusted and BMI-adjusted glucose.**

For our Mendelian randomization (MR) analyses we used fasting glucose-associated genetic variants identified by a GWAS that was adjusted for body mass index (BMI)[1]. In order to investigate whether such adjustment for BMI could have induced collider bias for our MR estimates, we explored how glucose GWAS effect sizes changed for on adjustment for BMI in UKB. We conducted GWAS of natural log-transformed random glucose (mmol/L) in 370,554 European ancestry UKB participants, adjusted and unadjusted for BMI. To account for sample relatedness and population stratification, we fit a linear mixed model (LMM) using the fastGWA method[46] implemented in the GCTA[9] software package version 1.93.3beta2. fastGWA uses a genetic relationship matrix (GRM), which we calculated using GCTA, from directly genotyped SNPs that had undergone the centrally applied UKB QC protocol[20], as well as additional filters in PLINK version 1.9[33] including --geno 0.1, --hwe 0.000001 and --maf 0.01. We applied the default GRM sparsity threshold of 0.05. We conducted GWAS analyses using the residuals from regression of log glucose on offspring sex, age at outcome measurement (including linear, quadratic and sex interaction effects), time since last meal, genotyping batch, study centre, the top 20 genetic principal components, and BMI (for BMI adjusted models only) as the phenotype.

We examined GWAS effect sizes with and without adjustment for BMI, for 124 independent variants with unadjusted $P < 5\times10^{-8}$. Regression of GWAS effect estimates with BMI adjustment on those without BMI adjustment demonstrated strong agreement between the two (intercept = $5.15\times10^{-5}$, slope = 1.007, $R^2$=0.998, $P$ value for the null hypothesis that the slope was equal to 1 = 0.166). Similar results were obtained when using 726 independent variants with unadjusted $P$ <0.001 (intercept = $2.11\times10^{-5}$, slope = 0.983, $R^2$=0.996, $P$ value for the null hypothesis that the slope was equal to 1 = $8.75\times10^{-10}$). The figures below illustrate these regression models. We concluded that random glucose GWAS effect estimates change little on adjustment for BMI. On the assumption that the same would also be true for fasting glucose GWAS summary statistics, it follows that our MR analyses would not be meaningfully affected by any collider bias due to adjustment for BMI in the original Chen et. al., 2021[1] fasting glucose GWAS.

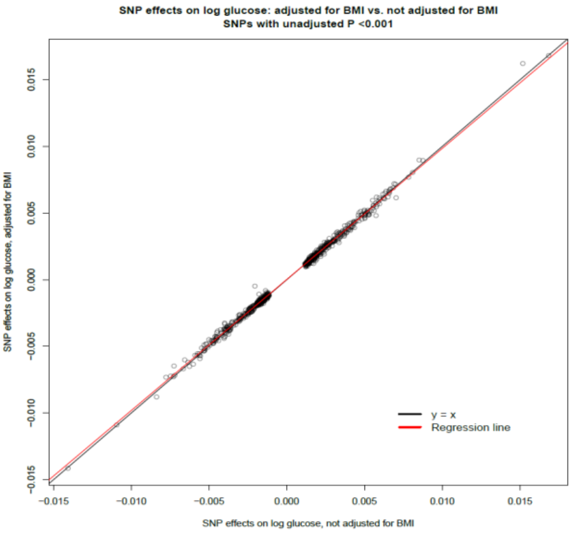

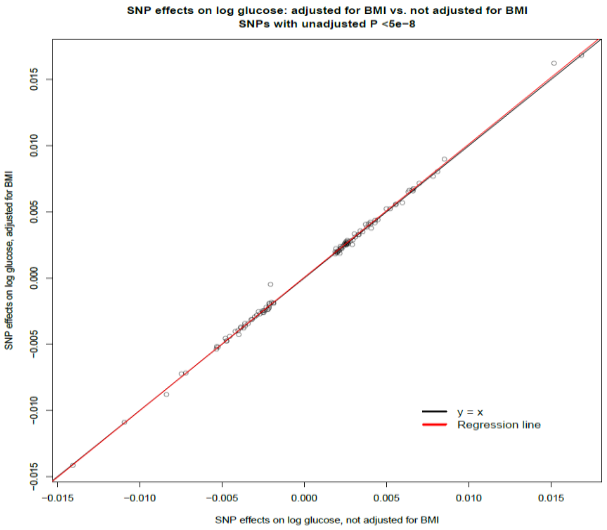

### References

1. Chen J, Spracklen CN, Marenne G, et al. The trans-ancestral genomic architecture of glycemic traits. *Nature Genetics*. 2021/06/01 2021;53(6):840-860. doi:10.1038/s41588-021-00852-9
2. Vujkovic M, Keaton JM, Lynch JA, et al. Discovery of 318 new risk loci for type 2 diabetes and related vascular outcomes among 1.4 million participants in a multi-ancestry meta-analysis. *Nature Genetics*. 2020/07/01 2020;52(7):680-691. doi:10.1038/s41588-020-0637-y
3. Elliott A, Walters RK, Pirinen M, et al. Distinct and shared genetic architectures of gestational diabetes mellitus and type 2 diabetes. *Nature Genetics*. 2024/03/01 2024;56(3):377-382. doi:10.1038/s41588-023-01607-4
4. Wright J, Small N, Raynor P, et al. Cohort Profile: The Born in Bradford multi-ethnic family cohort study. *International journal of epidemiology*. 2013;42(4):978-991. doi:10.1093/ije/dys112
5. Altshuler DM, Gibbs RA, Peltonen L, et al. Integrating common and rare genetic variation in diverse human populations. *Nature*. Sep 2 2010;467(7311):52-8. doi:10.1038/nature09298
6. McCarthy S, Das S, Kretzschmar W, et al. A reference panel of 64,976 haplotypes for genotype imputation. *Nature Genetics*. 2016/10/01 2016;48(10):1279-1283. doi:10.1038/ng.3643
7. Xue A, Wu Y, Zhu Z, et al. Genome-wide association analyses identify 143 risk variants and putative regulatory mechanisms for type 2 diabetes. *Nature Communications*. 2018/07/27 2018;9(1):2941. doi:10.1038/s41467-018-04951-w
8. Chang CC, Chow CC, Tellier LC, Vattikuti S, Purcell SM, Lee JJ. Second-generation PLINK: rising to the challenge of larger and richer datasets. *Gigascience*. 2015;4:7. doi:10.1186/s13742-015-0047-8
9. Yang J, Lee SH, Goddard ME, Visscher PM. GCTA: a tool for genome-wide complex trait analysis. *American journal of human genetics*. 2011;88(1):76-82. doi:10.1016/j.ajhg.2010.11.011
10. Åsvold BO, Langhammer A, Rehn TA, et al. Cohort Profile Update: The HUNT Study, Norway. *International journal of epidemiology*. May 17 2022;doi:10.1093/ije/dyac095
11. Brumpton BM, Graham S, Surakka I, et al. The HUNT study: A population-based cohort for genetic research. *Cell Genomics*. 2022/10/12/ 2022;2(10):100193. doi:https://doi.org/10.1016/j.xgen.2022.100193
12. Krokstad S, Langhammer A, Hveem K, et al. Cohort Profile: The HUNT Study, Norway. *International journal of epidemiology*. 2012. 9 August 2012. doi:10.1093/ije/dys095 Accessed Aug 9. http://www.ncbi.nlm.nih.gov/pubmed/22879362

    http://ije.oxfordjournals.org/content/early/2012/08/09/ije.dys095.full.pdf
13. Næss M, Kvaløy K, Sørgjerd EP, et al. Data Resource Profile: The HUNT Biobank. *International journal of epidemiology*. 2024;53(3):dyae073. doi:10.1093/ije/dyae073
14. Das S, Forer L, Schönherr S, et al. Next-generation genotype imputation service and methods. *Nature Genetics*. 2016/10/01 2016;48(10):1284-1287. doi:10.1038/ng.3656
15. Wang C, Zhan X, Bragg-Gresham J, et al. Ancestry estimation and control of population stratification for sequence-based association studies. *Nat Genet*. Apr 2014;46(4):409-15. doi:10.1038/ng.2924
16. Li JZ, Absher DM, Tang H, et al. Worldwide human relationships inferred from genome-wide patterns of variation. *Science*. Feb 22 2008;319(5866):1100-4. doi:10.1126/science.1153717
17. Norwegian Institute of Public Health. Medical Birth Registry of Norway Available at: www.fhi.no/en/hn/health-registries/medical-birth-registry-of-norway/medical-birth-registry-of-norway.
18. Moth FN, Sebastian TR, Horn J, Rich-Edwards J, Romundstad PR, Åsvold BO. Validity of a selection of pregnancy complications in the Medical Birth Registry of Norway. *Acta Obstet Gynecol Scand*. May 2016;95(5):519-27. doi:10.1111/aogs.12868
19. UK Biobank. 2006-2010, https://www.ukbiobank.ac.uk/, Accessed 01 February 2019.
20. Bycroft C, Freeman C, Petkova D, et al. The UK Biobank resource with deep phenotyping and genomic data. *Nature*. Oct 2018;562(7726):203-209. doi:10.1038/s41586-018-0579-z

21. Huang J, Howie B, McCarthy S, et al. Improved imputation of low-frequency and rare variants using the UK10K haplotype reference panel. *Nature Communications*. 2015/09/14 2015;6(1):8111. doi:10.1038/ncomms9111
22. Howie B, Fuchsberger C, Stephens M, Marchini J, Abecasis GR. Fast and accurate genotype imputation in genome-wide association studies through pre-phasing. *Nature Genetics*. 2012/08/01 2012;44(8):955-959. doi:10.1038/ng.2354
23. Auton A, Abecasis GR, Altshuler DM, et al. A global reference for human genetic variation. *Nature*. 2015/10/01 2015;526(7571):68-74. doi:10.1038/nature15393
24. Abraham G, Qiu Y, Inouye M. FlashPCA2: principal component analysis of Biobank-scale genotype datasets. *Bioinformatics*. 2017;33(17):2776-2778. doi:10.1093/bioinformatics/btx299
25. Fraser A, Macdonald-Wallis C, Tilling K, et al. Cohort Profile: the Avon Longitudinal Study of Parents and Children: ALSPAC mothers cohort. *International journal of epidemiology*. 2013;42(1):97-110. doi:10.1093/ije/dys066
26. Taylor AE, Jones HJ, Sallis H, et al. Exploring the association of genetic factors with participation in the Avon Longitudinal Study of Parents and Children. *International journal of epidemiology*. 2018;47(4):1207-1216. doi:10.1093/ije/dyy060
27. Warrington NM, Beaumont RN, Horikoshi M, et al. Maternal and fetal genetic effects on birth weight and their relevance to cardio-metabolic risk factors. *Nature Genetics*. 2019/05/01 2019;51(5):804-814. doi:10.1038/s41588-019-0403-1
28. Juliusdottir T, Steinthorsdottir V, Stefansdottir L, et al. Distinction between the effects of parental and fetal genomes on fetal growth. *Nat Genet*. Aug 2021;53(8):1135-1142. doi:10.1038/s41588-021-00896-x
29. Loh P-R, Tucker G, Bulik-Sullivan BK, et al. Efficient Bayesian mixed-model analysis increases association power in large cohorts. *Nature Genetics*. 2015/03/01 2015;47(3):284-290. doi:10.1038/ng.3190
30. Moen GH, Brumpton B, Willer C, et al. Mendelian randomization study of maternal influences on birthweight and future cardiometabolic risk in the HUNT cohort. *Nat Commun*. Oct 26 2020;11(1):5404. doi:10.1038/s41467-020-19257-z
31. Wang G, Bhatta L, Moen GH, et al. Investigating a Potential Causal Relationship Between Maternal Blood Pressure During Pregnancy and Future Offspring Cardiometabolic Health. *Hypertension*. Jan 2022;79(1):170-177. doi:10.1161/hypertensionaha.121.17701
32. Manichaikul A, Mychaleckyj JC, Rich SS, Daly K, Sale M, Chen WM. Robust relationship inference in genome-wide association studies. *Bioinformatics*. Nov 15 2010;26(22):2867-73. doi:10.1093/bioinformatics/btq559
33. Purcell S, Neale B, Todd-Brown K, et al. PLINK: a tool set for whole-genome association and population-based linkage analyses. *American journal of human genetics*. Sep 2007;81(3):559-75. doi:10.1086/519795
34. Holmen J, Holmen TL, Tverdal A, Holmen OL, Sund ER, Midthjell K. Blood pressure changes during 22-year of follow-up in large general population - the HUNT Study, Norway. *BMC Cardiovascular Disorders*. 2016;16(1)doi:10.1186/s12872-016-0257-8
35. Friedewald WT, Levy RI, Fredrickson DS. Estimation of the concentration of low-density lipoprotein cholesterol in plasma, without use of the preparative ultracentrifuge. *Clin Chem*. Jun 1972;18(6):499-502.
36. Tobin MD, Sheehan NA, Scurrah KJ, Burton PR. Adjusting for treatment effects in studies of quantitative traits: antihypertensive therapy and systolic blood pressure. *Statistics in medicine*. Oct 15 2005;24(19):2911-35. doi:10.1002/sim.2165
37. Ani A, van der Most PJ, Snieder H, Vaez A, Nolte IM. GWASinspector: comprehensive quality control of genome-wide association study results. *Bioinformatics*. Apr 9 2021;37(1):129-130. doi:10.1093/bioinformatics/btaa1084

38. Bulik-Sullivan BK, Loh PR, Finucane HK, et al. LD Score regression distinguishes confounding from polygenicity in genome-wide association studies. *Nat Genet*. Mar 2015;47(3):291-5. doi:10.1038/ng.3211
39. Willer CJ, Li Y, Abecasis GR. METAL: fast and efficient meta-analysis of genomewide association scans. *Bioinformatics*. 2010;26(17):2190-2191. doi:10.1093/bioinformatics/btq340
40. Beaumont RN, Flatley C, Vaudel M, et al. Genome-wide association study of placental weight identifies distinct and shared genetic influences between placental and fetal growth. *Nat Genet*. Nov 2023;55(11):1807-1819. doi:10.1038/s41588-023-01520-w
41. Warrington NM, Hwang LD, Nivard MG, Evans DM. Estimating direct and indirect genetic effects on offspring phenotypes using genome-wide summary results data. *Nat Commun*. Sep 14 2021;12(1):5420. doi:10.1038/s41467-021-25723-z
42. Wu Y, Zhong X, Lin Y, et al. Estimating genetic nurture with summary statistics of multigenerational genome-wide association studies. *Proc Natl Acad Sci U S A*. Jun 22 2021;118(25)doi:10.1073/pnas.2023184118
43. Bulik-Sullivan B, Finucane HK, Anttila V, et al. An atlas of genetic correlations across human diseases and traits. *Nature Genetics*. 2015/11/01 2015;47(11):1236-1241. doi:10.1038/ng.3406
44. Magnus P, Birke C, Vejrup K, et al. Cohort Profile Update: The Norwegian Mother and Child Cohort Study (MoBa). *International journal of epidemiology*. Apr 2016;45(2):382-8. doi:10.1093/ije/dyw029
45. Elizabeth CC, Oleksandr F, Alexey AS, et al. The Norwegian Mother, Father, and Child cohort study (MoBa) genotyping data resource: MoBaPsychGen pipeline v.1. *bioRxiv*. 2022:2022.06.23.496289. doi:10.1101/2022.06.23.496289
46. Jiang L, Zheng Z, Qi T, et al. A resource-efficient tool for mixed model association analysis of large-scale data. *Nature Genetics*. 2019/12/01 2019;51(12):1749-1755. doi:10.1038/s41588-019-0530-8
